# Influenza outbreak, disease transmission rate and mortality risk in the United States (2021 to 2050 Projection)

**DOI:** 10.1101/2021.04.16.21255632

**Authors:** Peter M. Etaware

**Author notes:** Corresponding Author Contact: 12, John Etaware Street, Tedi, P. O. Box 282, Navy Town, Lagos State, Nigeria.

## Abstract

Influenza and COVID-19 pandemics are among the deadliest respiratory diseases recorded in the history of humanity. Influenza infections are difficult to diagnose and nearly impossible to predict because the disease symptoms changes as the pathogen(s) evolve. The U.S. was the case study for this research (Latitude: 37.0902^°^N and Longitude: 95.7129°W). The principle employed in the design of the influenza forecast model was “The Integrated Independent System of Disease Prediction (IISDP)”, where the functionality of one forecast model depends on the predictive capacity of one or more forecast models. It was predicted that 49,734,427 individuals will be infected in 2021 out of the current estimation of 322,900.000 U.S. citizens (only about 1.5% of the total population). The situation was estimated by Etaware-Pred-2021 to worsen in 2050 (300,803,433 infected individuals out of the estimated 400,000,000 U.S. citizens), with a geometric increase in the ratio of infected individuals from 1.5% (in 2021) to 75.2% (in 2050), if necessary health precautions fail. unless there are potent influenza vaccines in circulation, total compliance to influenza vaccination and a high level of personal hygiene among U.S. citizens, influenza infection will become a global threat to humanity. These situations can be averted if the U.S. citizens act accordingly.

## 1.0 Introduction

The new Influenza outbreak and the deadly COVID-19 pandemic are among the worst ever recorded life-threatening diseases in the history of humanity. These respiratory ailments have similar symptoms and devastating effects both in endemic and pandemic proportions [1]. Sadly, human populations (regardless of age, gender, race, colour or religion) have very little or no pre-existing herd immunity against these diseases [1], the only options available are rapid vaccine development and mass vaccination to increase global herd immunity. The Influenza disease is a biological “time bomb waiting to explode” based on the fact that it’s outbreak is nearly impossible to predict since the disease symptoms changes as the pathogen(s) evolve [1]. The influenza outbreak is a seasonal flu that is responsible for the death of close to 500,000 individuals annually and over 1 billion deaths have been recorded globally throughout the history of the disease [2]. The first wave of pandemic in the 21^st^ century occurred in 2009 and was caused by influenza A (subtype H1N1) virus. It was estimated to have killed between 100,000 and 400,000 people globally in the first year alone [1].

The criteria for medical diagnosis of influenza ailment was first provided by Hippocrates in 400BCE [3, 4]. The symptoms of the disease include fever, cough, sore throat, runny or stuffy nose, muscle or body aches, headaches, fatigue, vomiting, diarrhoea etc. [5], which is synonymous to the symptoms of other severe respiratory diseases like SARS, MERS and COVID-19 etc. [6], making it very difficult for precise medical diagnosis. According to WHO [7], there are four (4) types of seasonal influenza viruses i.e. Influenza A (H5N1, H9N2, subtypes (H2N8, H3N8, H2N2, H1N1, H3N2 and the mutagen “H1N2” etc.), B, C, and D viruses [7]. The first successful medical diagnosis of influenza disease took place in 10^th^ century Europe during the pandemic outbreak of influenza in 1173 [6] that killed both the elderly and the handicapped [8]. The term influenza was first used to describe the disease in Italy in 1357 [9, 10].

Thereafter, a new wave of influenza pandemic struck Asia in the 14^th^ century, precisely 1510 [8], where it spread northwards to North Africa and Europe, and westwards, through travels aboard European vessels, across the Atlantic Ocean to America, Oceania/Australia, Antarctica and the rest of the world, causing another wave of disease pandemic which lasted for 2 years (1557-1558) [8]. The influenza disease morbidity was relatively high as it spread faster, fortunately, the mortality (Death rate) was low and only restricted to immunocompromised or immunosuppressed individuals, children and the aged [8]. This was the first pandemic recorded in history to be associated with miscarriages and foetal mortality [11]. Indeed, it was undoubtedly the historical advent of the spread of Influenza pandemic worldwide.

Currently, in the 21^st^ century, the United States is one of the most affected countries worldwide, with severe and recurrent influenza attacks. Well over 30.9 million cases were reported in 2017 and another 14.5 million influenza-related cases recorded in hospitals within the United States alone. Also, about 143 deaths (Influenza and Pneumonia) were recorded in every 1,000,000 individuals encountered in 2017 [12]. Initially, Heart disease was the leading cause of death in the United States, accounting for 23% of all deaths, alongside cancer, accidents, chronic lower respiratory diseases and cerebrovascular diseases etc., but influenza and pneumonia are fast becoming a nightmare in recent years [12]. Influenza infections are associated with thousands of deaths every year in the United States, mostly among adults aged ≥65 years [13].

The Centre for Disease Control of the United States have made it a point of duty to estimate annual influenza outbreaks and mortality in the U.S., in order to improve researches conducted globally on influenza and also, enhance the efficacy of management policies and strategies developed thereafter [13]. Therefore, in consortium with this policy, the current study was thematically synchronized with the aims and objectives of the CDC of U.S.A. to evaluate past influenza outbreaks and associated deaths in the United States in order to develop a functional system for decision making, through quality prediction of influenza outbreak across the U.S., effective estimation of morbidity risk and a quantitative description of the expected mortality rate in the United States. This will further increase the level of awareness of the disease among the U.S. populace and further facilitate a beef up in the level of preparedness of both government and health officials in combating the disease when the need arises.

## 1.1 Hypotheses

### Hypothesis 1

The number of U.S. citizens vaccinated (annually), herd immunity or acquired immunity, and the susceptibility level of individuals to influenza infection have no significant effects on the rising incidence of seasonal influenza outbreak and spread in the U.S.

**H**_**o1**_: β_1_ = β_2_ = ……………………..β_n_ = 0

Or

There are underlying medical relationships between the variables listed and influenza outbreak in the U.S.

**H**_**a1**_: β_1_ ≠ 0 or β_2_ ≠ 0 ……………or β_n_ ≠ 0

### Hypothesis 2

The increasing death toll from influenza infection is not affected by the rising cases of seasonal influenza outbreak and spread in the U.S., and the rapidly increasing U.S. population (either by natality or immigration).

**H**_**o2**_: β_1_ = β_2_ = ………………………β_3_ = 0

Or

There are significant medical interactions between the outlined predictors and influenza mortality in the U.S.

**H**_**a2**_: β_1_ ≠ 0 or β_2_ ≠ 0 ……………or β_3_ ≠ 0

## 1.2 Research Assumptions

1. The United States is vulnerable to massive seasonal influenza outbreak i.e. mild or sporadic or endemic or pandemic proportions.
2. All U.S. citizens (regardless of their race, gender, status, colour or religion) are susceptible to influenza infections (irrespective of the identity of the causal viral agent). Therefore, the research domain was hinged on the assumption that the U.S. population, in its entirety, was at risk of influenza infection.
3. The magnitude of seasonal outbreak of influenza infection is not restricted to a single pathotype or subtype of the influenza virus.
4. The duration for vaccine development and administration has no significant effects on the outcome and data collected annually on influenza outbreak or mortality in the U.S.
5. The causal agent of the disease is indeed infectious and the mode of transmission of the disease is by human-human or nosocomial (asymptomatic transmission). The research was indeed relaxed or silent on zoonosis, partly due to the fact that influenza D virus was only restricted to animals, as at the time of filing in this research.

## 2.0 Methodology

### 2.1 The study region

The study area was located on Latitude 37.0902°N and Longitude 95.7129°W in the great continent of North America (Fig 1). The United States is a conglomerate of fifty (50) States and a federal district, Washington D.C., (Fig 2) with annual temperature range between -3°C (Alaska) and 21.5°C (Florida), average precipitation (Rainfall) of 767mm and a relative humidity range between 38.3% (Nevada) and 77.1% (Alaska).

**Fig 1.**
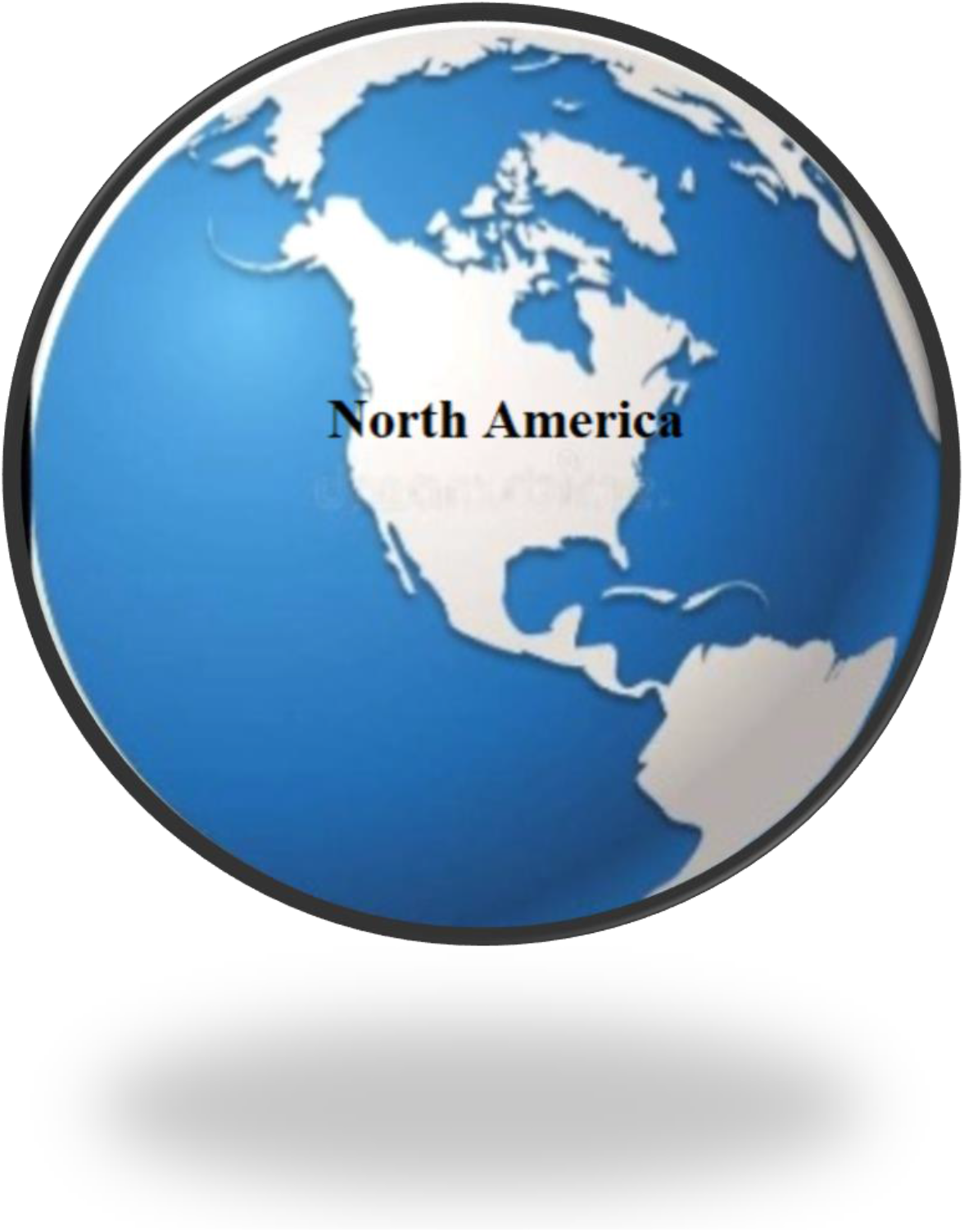
The geographical location of North America in the world map.

**Fig 2.**
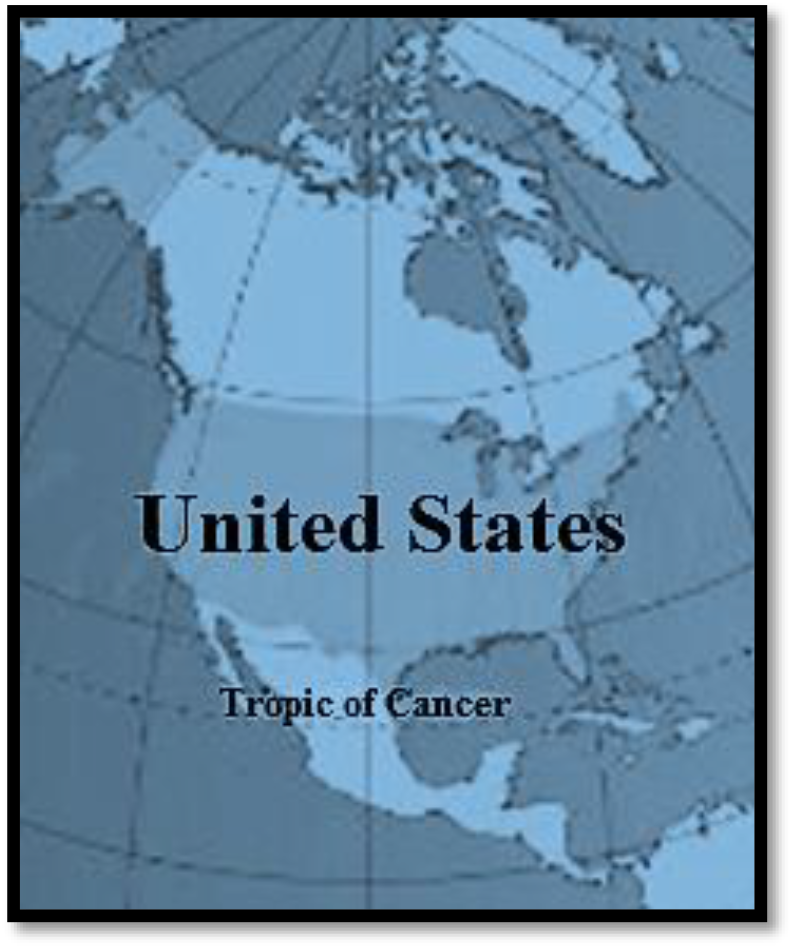
The location of the United States of America in the North American continent.

### 2.2 The U.S. Population Data

Census data and estimated values of the United States’ population from 1950 to 2060 were obtained from published articles of Statista Research Department [14], Ewert [15], Macrotrends [16] and the Federal Interagency Forum on Child and Family Statistics [17].

### 2.3 The U.S. population growth rate

The U.S. population growth rate was estimated by the formula:

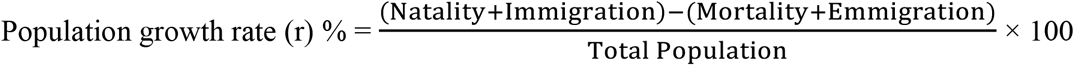

Therefore,

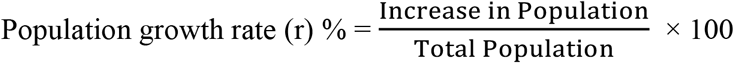

### 2.4 Influenza outbreak and morbidity data

The morbidity data for influenza outbreak in the U.S. was obtained from the published articles of Elflein [18], CDC [19] and Wikipedia [20]. The data collected was from 2010-2020 (10years).

### 2.5 The U.S. Stat on annual vaccination

The stats for annual influenza vaccination in the United States was obtained from Elflein [21].

### 2.6 Influenza mortality data

The mortality data from 1950-2017 (67years) spanning through six (6) decades and 67 seasons of documented cases of death from influenza outbreak in the U.S. was obtained from Statista and the U.S. Centre for Disease Control [12, 13]. All recorded data for deaths with underlying pneumonia and influenza causes, and/or respiratory and circulatory causes were actual counts based on the information contained in the death certificate ICD codes referenced by the U.S. CDC Department [13].

### 2.7 The Infection rate

The Infection rate (IR) for influenza outbreak in the U.S. was the number of new cases identified in relation to the existing population of people at risk of contracting the flu within a specified time frame. It was calculated using the formula:

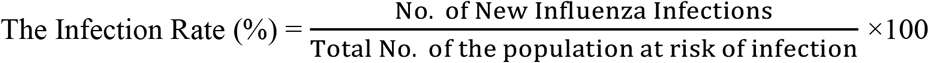

### 2.8 Acquired Immunity and Susceptibility Index

Acquired immunity (A.I.) is regarded as the disease resistance conferred on any population by virtue of the number of individuals vaccinated against that disease, while the susceptibility index (S.I.) is the likelihood of an individual, within the population, to contract the disease. They are determined thus:

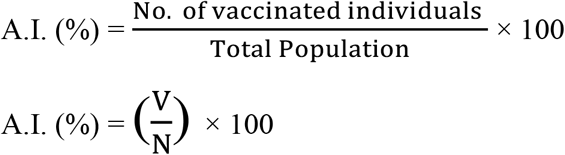

while,

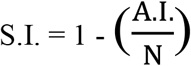

### 2.9 Mean, standard deviation and standard error of the mean

The population mean (μ), standard deviation (σ) and standard error of the mean (σ_ẍ_) were calculated by the formulae given below:

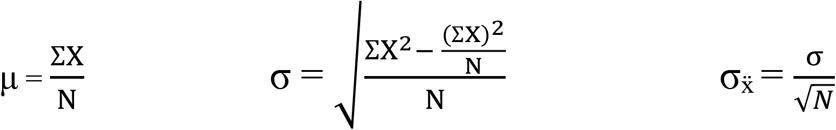

### 2.10 Mortality Risk

The mortality risk or case fatality rate is the measure of the likelihood or chances of death of an infected individual. This was determined by the formula:

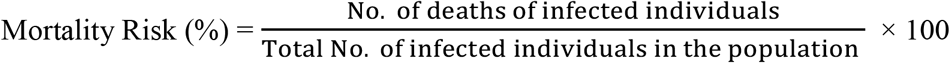

### 2.11 Mortality Rate

The mortality rate is a measure of the time limit through infection to death in a given population. It was calculated by the formula:

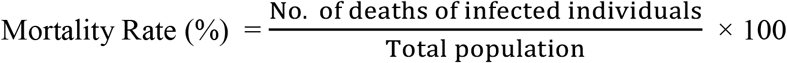

### 2.12 Data Analysis

The use of data mining and cleansing techniques facilitated the detection of misrepresented values, outliers and ambiguous datasets. COSTAT 6.451, Minitab 16.0 and SPSS 20.0 software were used for data analysis. The predictors were tested against the desired response variables using Pearson’s Product Moment of Correlation (PPMC), Cluster Analysis of predictors and Scatterplots. The models were structured using multiple linear regression equation and programmed on Microsoft Excel (2016). The quality of each models was determined by the values of the coefficient and Adjusted coefficient of correlation (R^2^ and Adj. R^2^), Mean Square Error of Prediction (MSEP.), Leave-Group-Out of Prediction (LGO Press), and Leave-Group-Out Prediction of the coefficient of correlation (LGOPreR^2^). The models developed were validated by bootstrapping and a backlog comparison with primary data. The test statistics used in discerning the relatedness of the estimated and actual results was the correlated T-test (P≤0.05). Graphs and figures were generated from Microsoft Office (2016).

## 3.0 Results

The principle employed in the design of the influenza forecast model was “The Integrated Independent System of Disease Prediction (IISDP)”, where the functionality of one forecast model depends on the predictive capacity of one or more forecast models. Therefore, the prediction of influenza outbreak in the United States was a function of the number of individuals vaccinated annually (Acquired Immunity) and the ratio of disease resistant to susceptible individuals in the population (Herd Immunity), vis a vis the U.S. population dynamics (migration, natality and mortality). Estimated data for vaccination was the most convenient and reliable source of predictor for the disease, as vaccination stimulates innate immunity which is the fundamental mechanism for collective defence (Herd Immunity).

The model for vaccination was carefully structured using the multiple regression equation described below (Data available in Supplementary File “S1 and S2”):

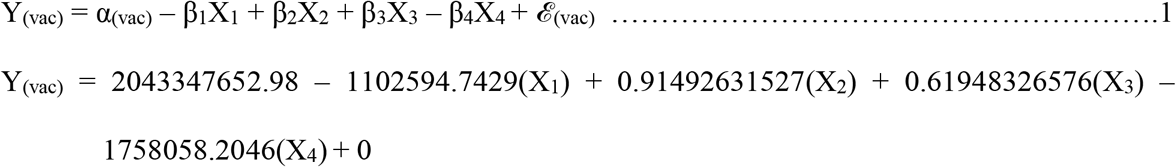

Where,

Y_(vac)_ = Vaccinated U.S. citizens (using age distribution)

X_1_ = The period of vaccination (Year)

X_2_ = Population size for each age distribution

X_3_ = Total Population

X_4_ = The percentage of the population of each age group within the population

*ℰ*_(vac)_ = The correction factor for the regression equation

α_(vac)_ = The Intercept on Y_(vac)_

The model statistics was stated below:

▪ R^2^ = 0.96
▪ MSEP = 5.77 × 10^12^
▪ Adj. R^2^ = 0.95
▪ Press = 1.98 × 10^14^
▪ Pre R^2^ = 0.95

Model Validation

▪ Method: Bootstrap
▪ Validate “N” Times: 10
▪ LGO Press = 8.75 × 10^13^
▪ LGO Pre R^2^ = 0.95

The forecast model for estimation of the minimum expected number of U.S. populace to be vaccinated (annually) against Influenza virus (regardless of the pathotype or subtype) was programmed on Microsoft Excel (2016) as shown in Fig 3.

**Fig 3.**
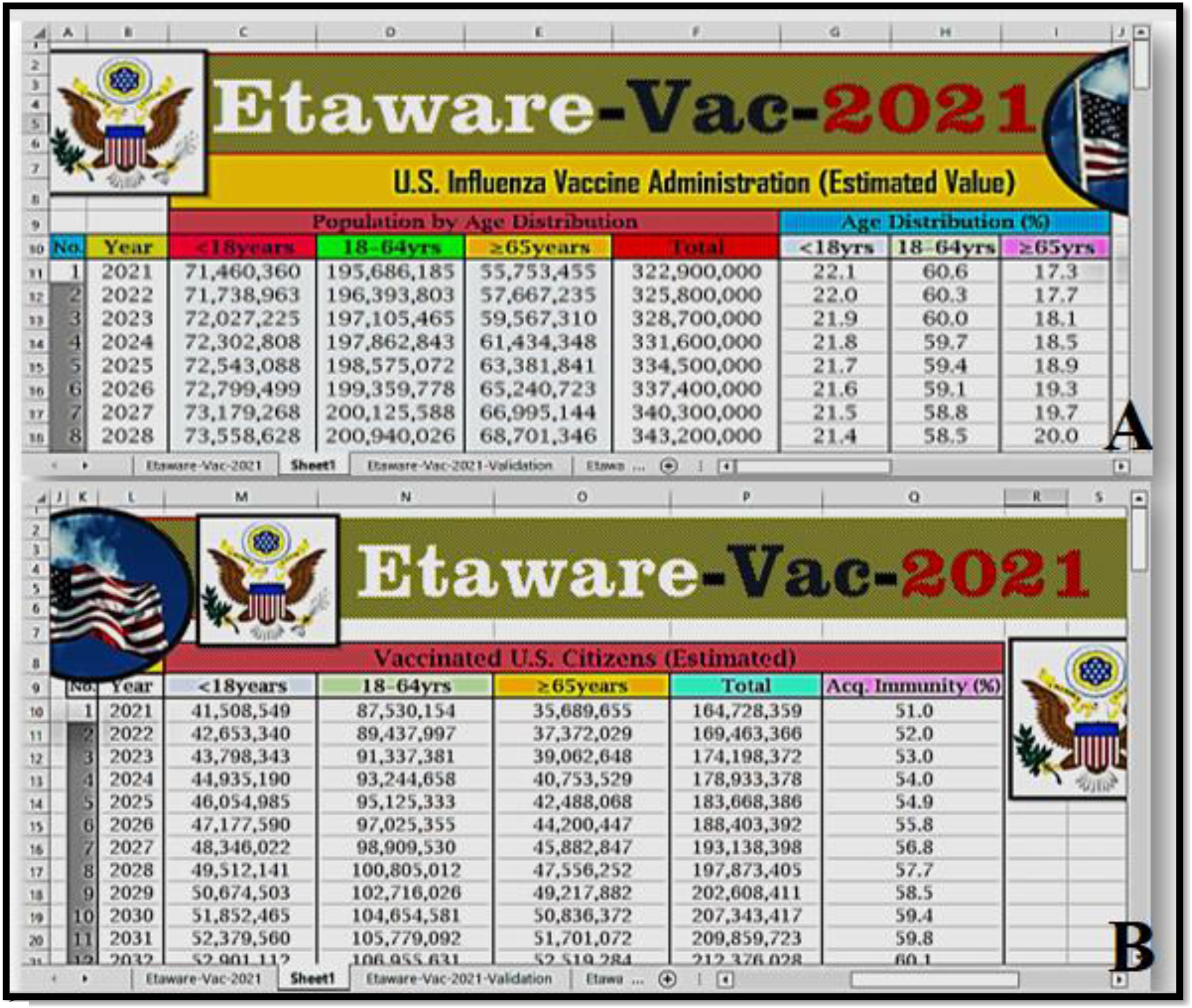
Etaware-Vac-2021 forecast model (A) Input (B) Output.

The structured model was further validated using real-life data recorded for U.S. senior citizens. There was no significant difference in the estimated number of U.S. senior citizens vaccinated over the years (2015 – 2020) and that of the actual values recorded by the U.S. CDC (T-test = -0.53, P = 0.62). Also, there was no significant difference in the estimated immunity level and those calculated for the actual population of senior citizens in the United States from 2015 to 2020 (T-test = -0.62, P = 0.56) as shown in Table 1. The correlation between the predicted and actual results was 0.84, while the coefficient of correlation was 0.71 (Table 1).

**Table 1.** Contrast between the observed and estimated values for U.S. senior citizens.

| Year | Acquired Immunity(Age≥65yrs) |  | Vaccinated (Age≥65yrs) |  | Population (Age≥65yrs) |
| --- | --- | --- | --- | --- | --- |
|  | Observed | Estimated | Observed | Estimated |  |
| 2015 | 66.7 | 70.8 | 31,174,914 | 33,082,326 | 46,739,001 |
| 2016 | 63.4 | 69.1 | 30,484,988 | 33,233,416 | 48,083,578 |
| 2017 | 65.3 | 67.5 | 32,297,015 | 33,400,333 | 49,459,441 |
| 2018 | 59.6 | 66.0 | 30,363,003 | 33,611,442 | 50,944,636 |
| 2019 | 68.1 | 64.5 | 35,766,204 | 33,860,397 | 52,520,123 |
| 2020 | 69.8 | 63.1 | 37,637,421 | 34,051,124 | 53,921,806 |
| Mean | 65.5 | 66.8 | 32,953,924 | 33,539,840 |  |
| SD | 3.63 | 2.88 | 3,041,348 | 372,014 |  |
| SE | 1.48 | 1.17 | 1,241,625 | 151,874 |  |
| T-test |  | -0.62 |  | -0.53 |  |
| P value |  | 0.56 |  | 0.62 |  |
| R |  | -0.33 |  | 0.84 |  |
| R<sup>2</sup> |  | 0.11 |  | 0.71 |  |

Mean values were declared statistically significant at $P \leq 0.05$ by T-test pairwise comparison (Supplementary File “S6”). SD – Standard Deviation, SE – Standard Error of the Mean.

A review of the analysis conducted on the results obtained from Etaware-Vac-2021 showed that there was a steady decline in the number of individuals vaccinated annually from 2011 to 2020, across all age groups in the United States (in relation to the actual population size of that age group) i.e. 43,730,468 (<18years), 86,122,878 (18-64years), and 32,453,641(≥65years) individuals were vaccinated as at 2011, while 40,350,848 (<18years), 85,591,381 (18-64years), and 34,051,124 (≥65years) turned out for vaccination in 2020 (Table 2), amidst the rising cases of influenza outbreak and the challenges associated with the geometric increase in population over the years. Also, the immunity level of the U.S. populace to influenza infection was on the decrease over the years i.e. From 52.3% in 2011 to 50.0% in 2020 (Table 2).

**Table 2.** Estimated values for influenza vaccination in the U.S. based on age distribution.

| Vaccinated U.S. Citizens (Estimated Values) |  |  |  |  |  |  |  |  |
| --- | --- | --- | --- | --- | --- | --- | --- | --- |
| Year | <18years |  | 18-64yrs |  | ≥65years |  | Total |  |
|  | Vaccinated | A.I. (%) | Vaccinated | A.I. (%) | Vaccinated | A.I. (%) | Vaccinated | A.I. (%) |
| 2011 | 43,730,468 | 59.4 | 86,122,878 | 44.1 | 32,453,641 | 78.9 | 162,306,987 | 52.3 |
| 2012 | 43,287,637 | 59.2 | 86,083,087 | 44.1 | 32,679,193 | 76.4 | 162,049,917 | 52.1 |
| 2013 | 42,881,621 | 59.0 | 86,096,785 | 44.0 | 32,814,440 | 74.4 | 161,792,845 | 51.8 |
| 2014 | 42,505,821 | 58.7 | 86,075,731 | 44.0 | 32,954,223 | 72.5 | 161,535,776 | 51.5 |
| 2015 | 42,147,890 | 58.4 | 86,048,489 | 44.0 | 33,082,326 | 70.8 | 161,278,705 | 51.3 |
| 2016 | 41,783,586 | 58.1 | 86,004,633 | 44.0 | 33,233,416 | 69.1 | 161,021,635 | 51.0 |
| 2017 | 41,406,013 | 57.7 | 85,958,218 | 44.0 | 33,400,333 | 67.5 | 160,764,564 | 50.8 |
| 2018 | 40,983,450 | 57.5 | 85,912,602 | 43.9 | 33,611,442 | 66.0 | 160,507,494 | 50.5 |
| 2019 | 40,566,180 | 57.2 | 85,823,847 | 43.9 | 33,860,397 | 64.5 | 160,250,424 | 50.3 |
| 2020 | 40,350,848 | 56.7 | 85,591,381 | 43.9 | 34,051,124 | 63.1 | 159,993,353 | 50.0 |

**Note:** The A.I. was calculated as a percentage of each vaccinated proportion in relation to the total population and that of the population of each age distribution presented in Supplementary file “S1”. A.I. = Acquired Immunity

The acquired immunity conferred on the populace through mass vaccination was further used to structure a predictive model for determination or estimation of annual increase in influenza infection rate within the United States (Fig 4). The model dynamics for estimation of influenza infection rate was described thus (Data available in Supplementary File “S3 and S4”):

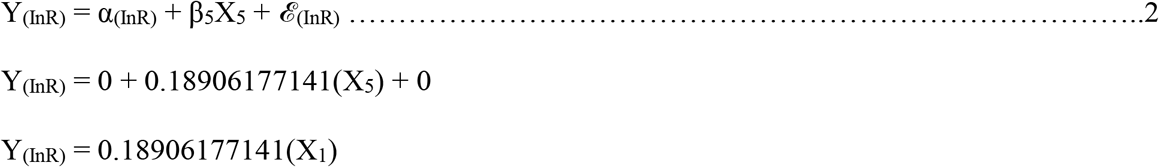

Where,

Y_(InR)_ = Influenza Infection Rate

X_5_ = Acquired Immunity (%)

α_(InR)_ = 0 (Suppressed)

*ℰ*_(InR)_ = 0 (Suppressed)

**Fig 4.**
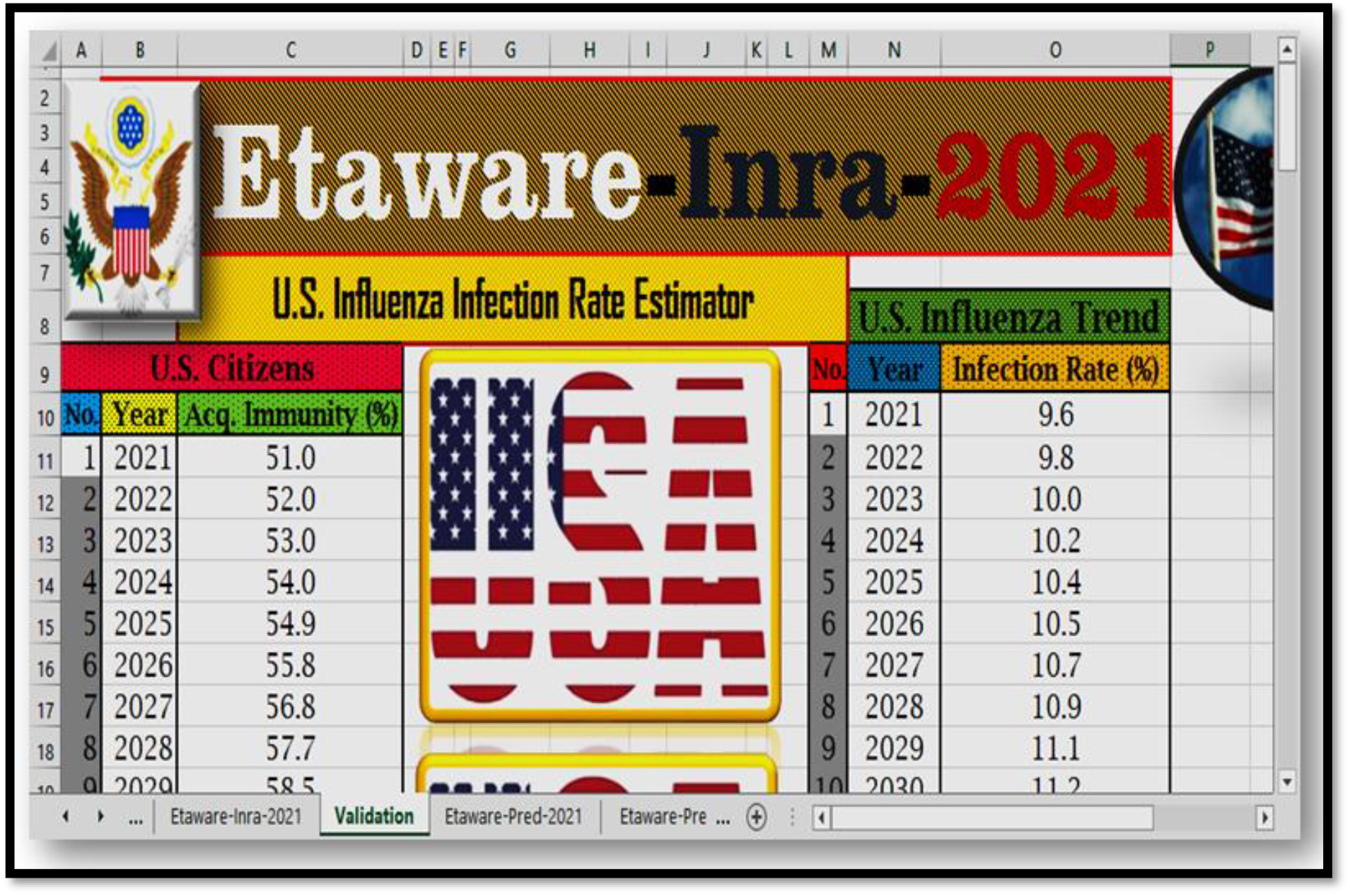
Etaware-Inra-2021 predictive model for influenza infection rate estimation in the U.S.

The model statistics was described as follow:

▪ R^2^ = 0.91
▪ MSEP = 11.52
▪ Adj. R^2^ = 0.90
▪ Press = 105.37
▪ Pre R^2^ = 0.95

Model validation

▪ Method: Bootstrap
▪ Validate “N” Times: 10
▪ LGO Pre R^2^ = 0.65

Further validation of the influenza infection rate model was carried out using past recorded data of influenza infection rate estimated in the United States. The test statistics conducted showed that there was no significant difference between the computer simulated values and the actual recorded values (T-test = -0.03, P = 0.98). Also, there was a strong relationship between the results obtained (R = -0.73 and R^2^ = 0.53) as shown in Table 3.

**Table 3.**
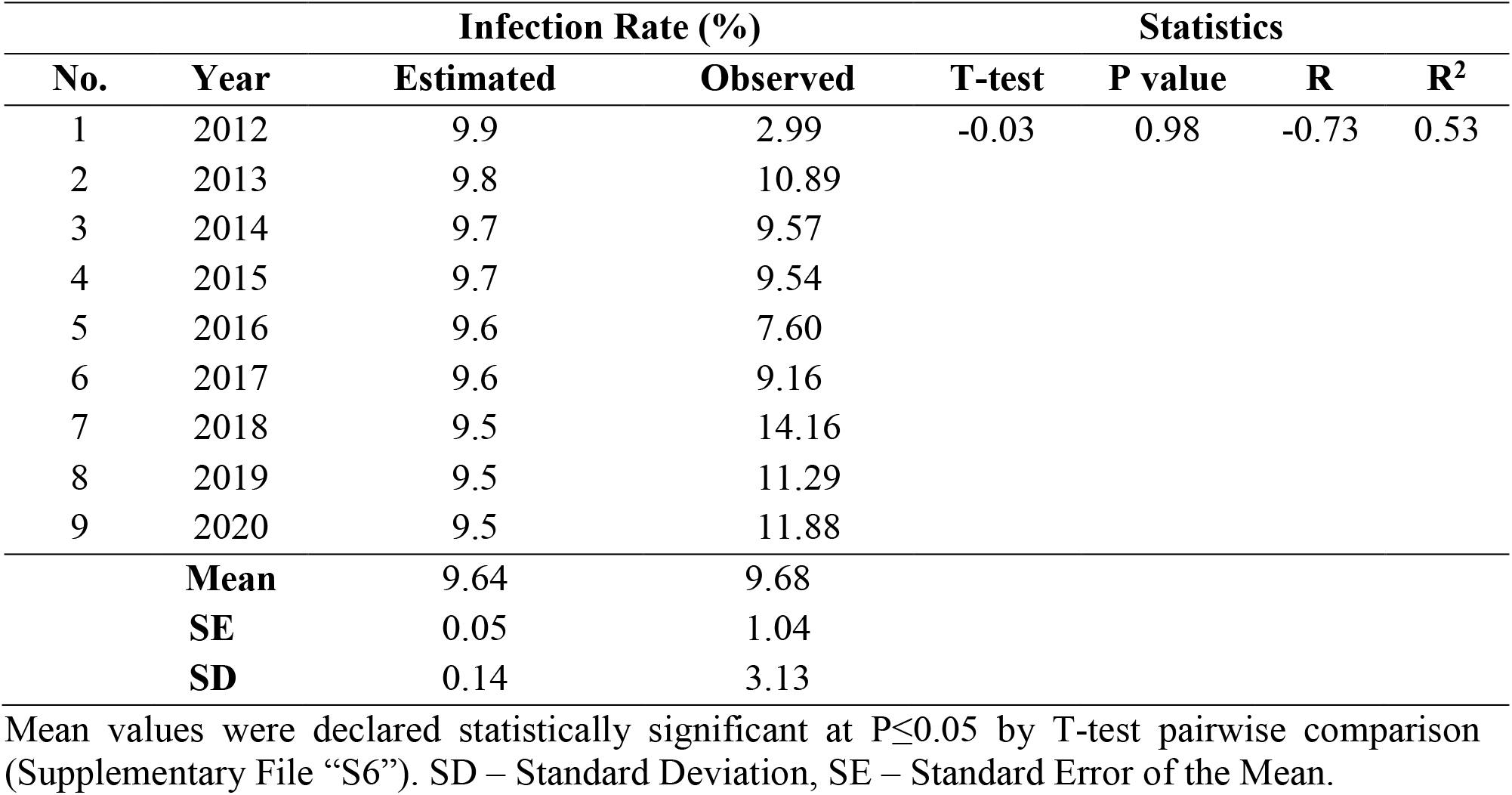
Correlation between the estimated and actual influenza infection rate in the U.S.

| No. | Year | Infection Rate (%) |  | Statistics |  |  |  |
| --- | --- | --- | --- | --- | --- | --- | --- |
|  |  | Estimated | Observed | T-test | P value | R | R<sup>2</sup> |
| 1 | 2012 | 9.9 | 2.99 | -0.03 | 0.98 | -0.73 | 0.53 |
| 2 | 2013 | 9.8 | 10.89 |  |  |  |  |
| 3 | 2014 | 9.7 | 9.57 |  |  |  |  |
| 4 | 2015 | 9.7 | 9.54 |  |  |  |  |
| 5 | 2016 | 9.6 | 7.60 |  |  |  |  |
| 6 | 2017 | 9.6 | 9.16 |  |  |  |  |
| 7 | 2018 | 9.5 | 14.16 |  |  |  |  |
| 8 | 2019 | 9.5 | 11.29 |  |  |  |  |
| 9 | 2020 | 9.5 | 11.88 |  |  |  |  |
|  | **Mean** | 9.64 | 9.68 |  |  |  |  |
|  | **SE** | 0.05 | 1.04 |  |  |  |  |
|  | **SD** | 0.14 | 3.13 |  |  |  |  |

Mean values were declared statistically significant at $P \leq 0.05$ by T-test pairwise comparison (Supplementary File “S6”). SD – Standard Deviation, SE – Standard Error of the Mean.

The predictors defined by the multiple regression models developed in the course of this research and those estimated by other researchers or those linked to the United States census board (actual and computer simulations) and the Centres for Disease Control were tested for their reliability and relatedness to influenza outbreak in the U.S. using PPMC, cluster analysis (Fig 5) or scatterplots fitted with regression lines (Fig 6) and connect (trend) lines (Fig 7). Majority of the predictors tested had >85% relatedness to the response variable, with a correlation factor of 1.00.

**Fig 5.**
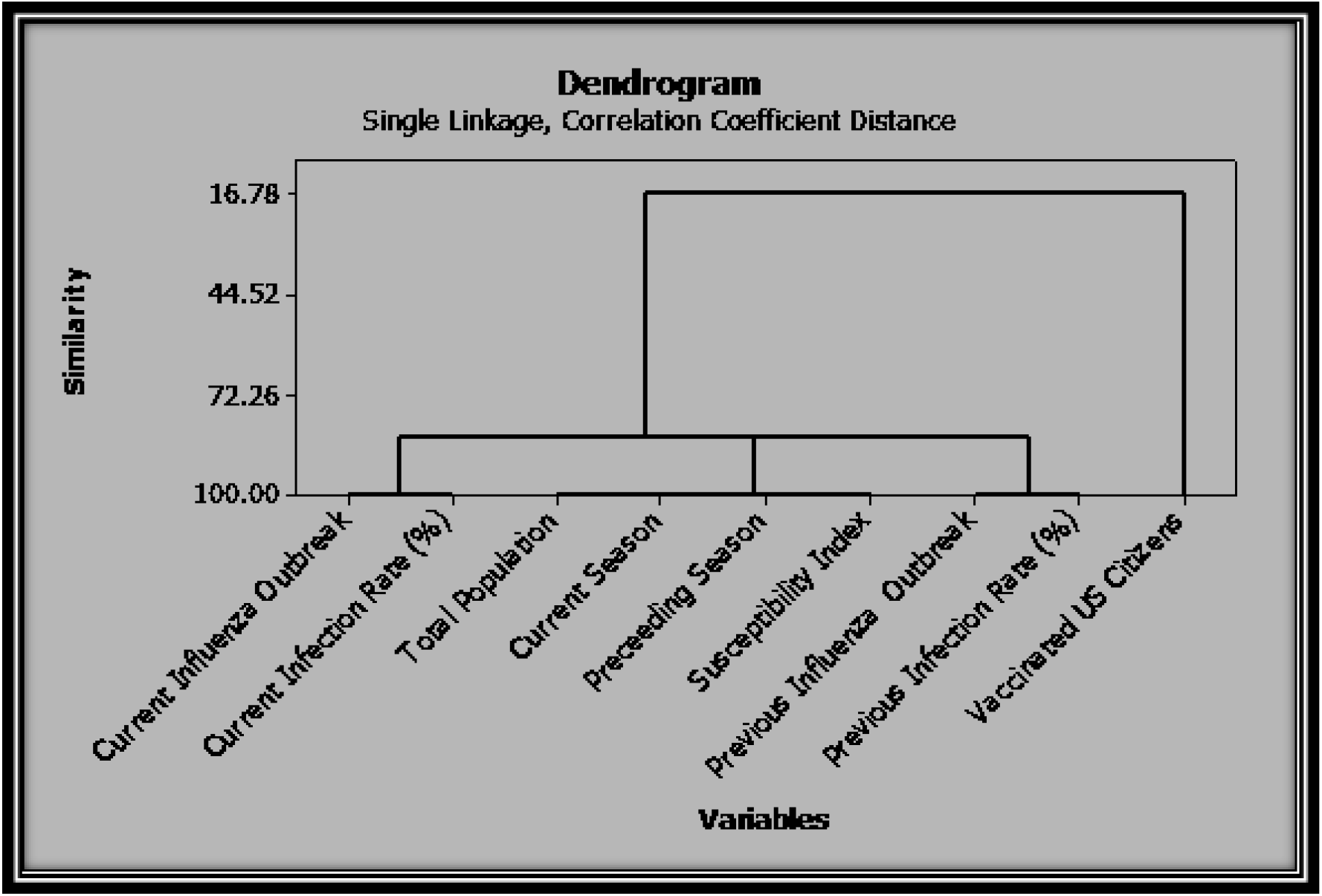
The cluster analysis of all predictors used for influenza outbreak model structuring.

**Fig 6.**
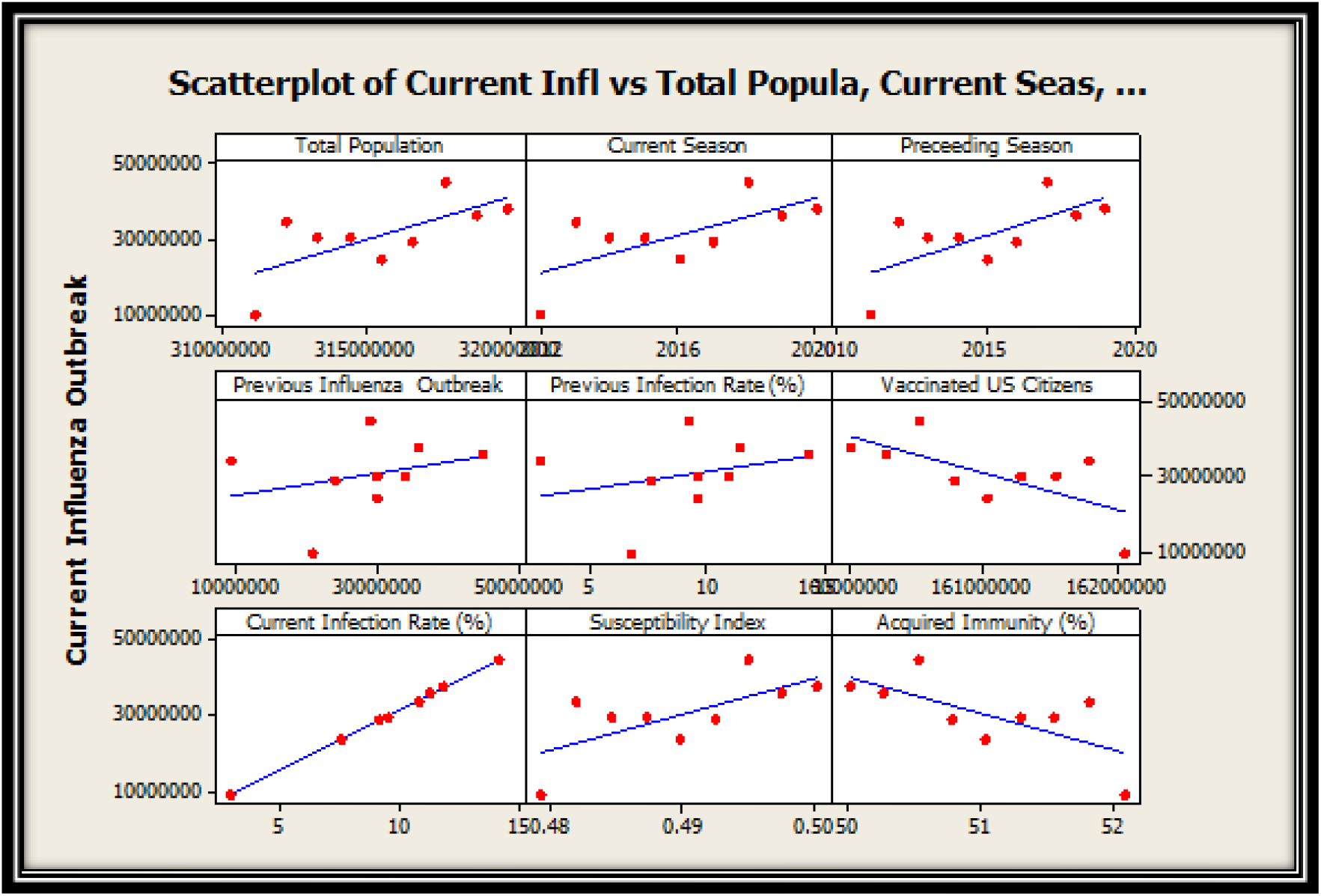
Correlation coefficient and reliability indices of the prospective influenza predictors.

**Fig 7.**
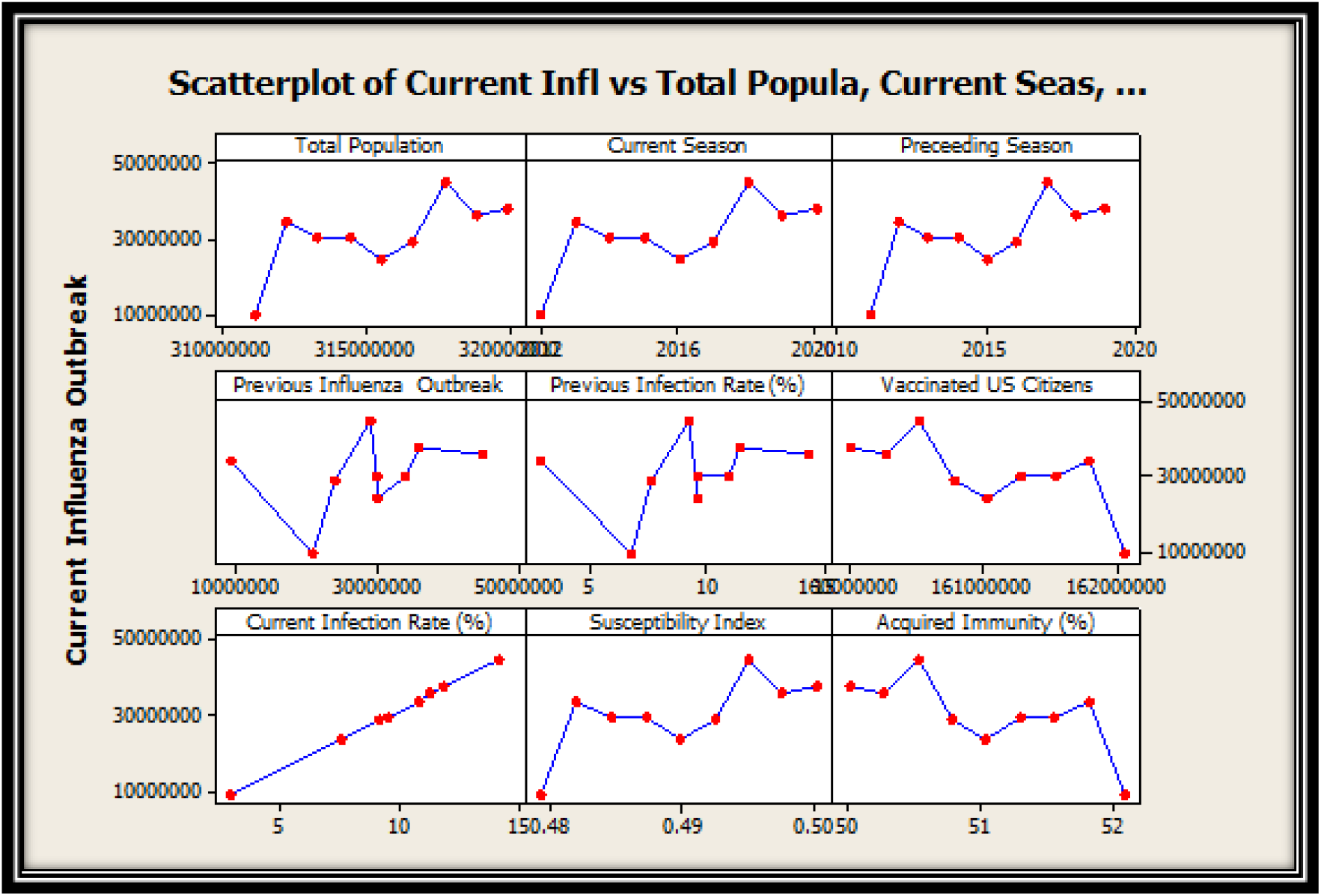
Pattern of relationship between each predictors and the response variable.

The structured models were incorporated into the influenza outbreak forecast system to enhance the model’s performance, predicative precision and result accuracy (Fig 8). The developed model was designed using the multiple regression equation stated thus (Data available in Supplementary File “S3 and S5”):

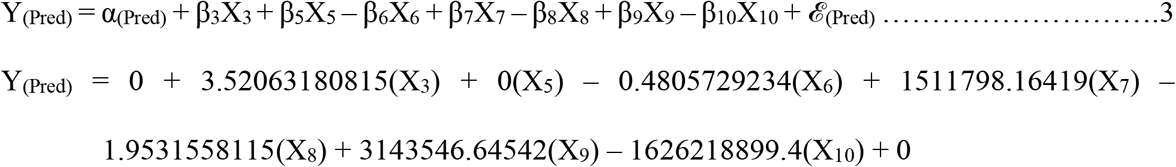

Where,

Y_(Pred)_ = Influenza outbreak

X_3_ = Total Population

X_5_ = Acquired immunity (%)

X_6_ = Previous influenza outbreak

X_7_ = Previous influenza infection rate (%)

X_8_ = Total vaccinated U.S. citizens

X_9_ = Current influenza infection rate (%)

X_10_ = Susceptibility index

α_(Pred)_ = 0 (Suppressed)

*ℰ*_(Pred)_ = 0 (Suppressed)

**Fig 8.**
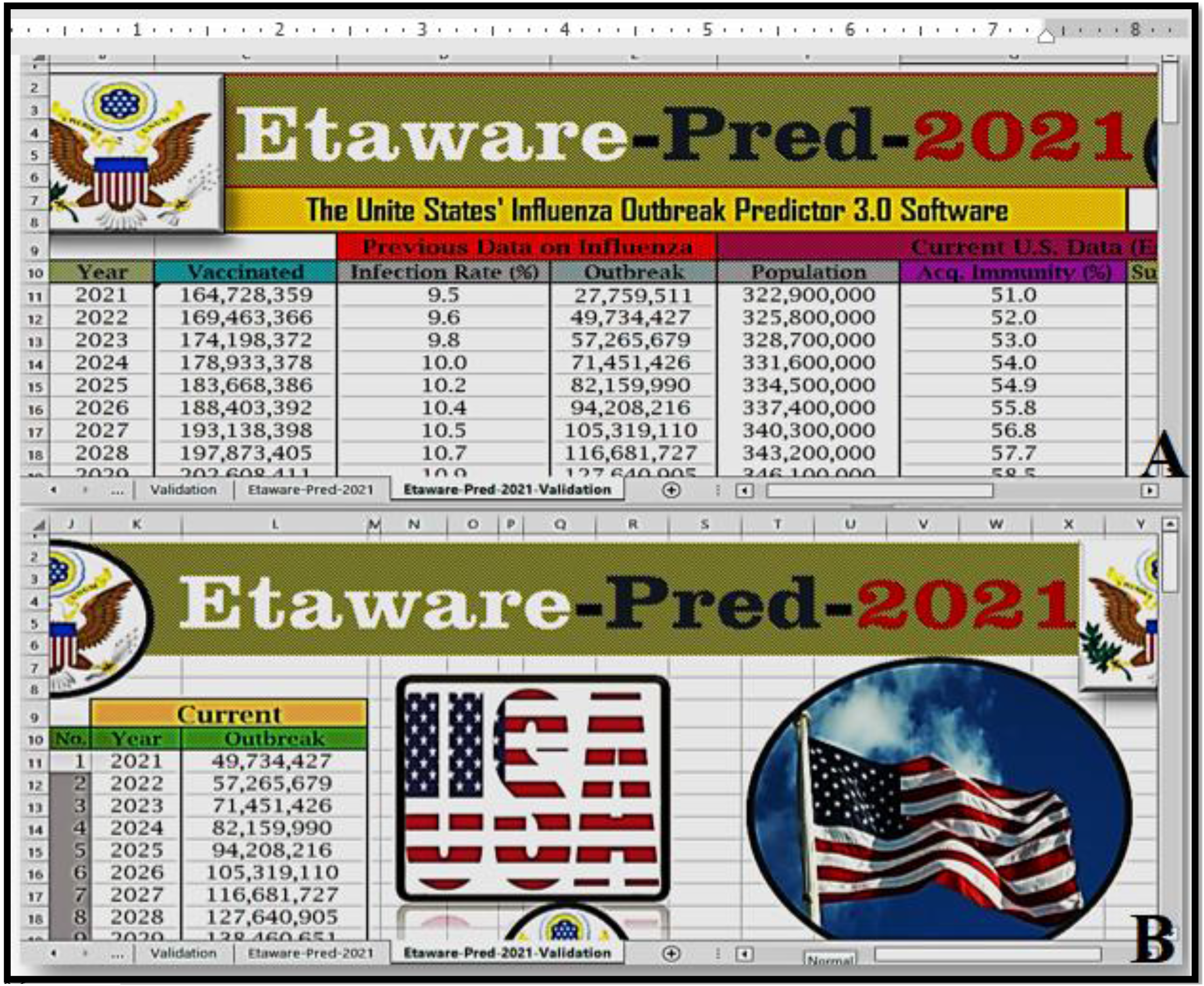
Etaware-Pred-2021 influenza outbreak forecast model (A) Input (B) Output.

The model statistics was described thus:

▪ R^2^ = 1.00
▪ MSEP = 90434009808.2
▪ Adj. R^2^ = 1.00
▪ Press = 747002587243
▪ Pre R^2^ = 1.00

Model Validation

▪ Method: Bootstrap
▪ Validate “N” Times: 10
▪ LGO Pre R^2^ = 1.00

Further validation was conducted using already established influenza outbreak data in the United States (2011 – 2020). The test statistics conducted (T-test = 0.55) showed that there was no significant difference in the estimated values for influenza outbreak in the United States and that of the actual recorded values obtained from the U.S. CDC (Table 4).

**Table 4.** The disparity between the estimated and actual influenza outbreak in the U.S.

| Influenza Outbreak |  |  | Statistics |  |  |  |
| --- | --- | --- | --- | --- | --- | --- |
| Year | Estimated | Observed | T-test | P value | R | R<sup>2</sup> |
| 2011 | 35,624,250 | 21,000,000 | 0.55 | 0.60 | -0.38 | 0.14 |
| 2012 | 35,528,201 | 9,300,000 |  |  |  |  |
| 2013 | 40,968,909 | 34,000,000 |  |  |  |  |
| 2014 | 28,918,526 | 30,000,000 |  |  |  |  |
| 2015 | 30,750,027 | 30,000,000 |  |  |  |  |
| 2016 | 30,658,093 | 24,000,000 |  |  |  |  |
| 2017 | 33,509,836 | 29,000,000 |  |  |  |  |
| 2018 | 31,073,518 | 45,000,000 |  |  |  |  |
| 2019 | 23,410,534 | 36,000,000 |  |  |  |  |
| 2020 | 27,759,511 | 38,000,000 |  |  |  |  |
| **Mean** | 31,397,462 | 30,588,889 |  |  |  |  |
| **SE** | 1,540,052 | 3,137,800 |  |  |  |  |
| **SD** | 4,967,134 | 10,021,033 |  |  |  |  |

Mean values were declared statistically significant at $P \leq 0.05$ by T-test pairwise comparison (Supplementary File “S6”). SD – Standard Deviation, SE – Standard Error of the Mean.

The minimum expected turn out for mass vaccination against influenza infection of the United States’ citizen was shown in Table 5. It is expected that the number of people that will be vaccinated in 2050 will almost double the amount recorded in 2021 i.e. 69,070,083 infants and youths below the age of 18 years, 140,847,200 persons between the ages of 18 and 64 years, and 72,712,632 senior citizens above the age of 65 years (2050) compared to 41,508,549 infants and youths below the age of 18 years, 87,530,154 young adults between the ages of 18 and 64 years old, and 35,689,655 senior citizens above the age of 65 years, estimated to undergo vaccination against influenza infection in 2021 (Table 5).

**Table 5.** Least projection of the expected population for influenza vaccination in the U.S.

| Minimum estimated mass vaccination in the U.S. by age groups |  |  |  |  |  |
| --- | --- | --- | --- | --- | --- |
| No. | Year | <18years | 18-64yrs | ≥65years | Total |
| 1 | 2021 | 41,508,549 | 87,530,154 | 35,689,655 | 164,728,359 |
| 2 | 2022 | 42,653,340 | 89,437,997 | 37,372,029 | 169,463,366 |
| 3 | 2023 | 43,798,343 | 91,337,381 | 39,062,648 | 174,198,372 |
| 4 | 2024 | 44,935,190 | 93,244,658 | 40,753,529 | 178,933,378 |
| 5 | 2025 | 46,054,985 | 95,125,333 | 42,488,068 | 183,668,386 |
| 6 | 2026 | 47,177,590 | 97,025,355 | 44,200,447 | 188,403,392 |
| 7 | 2027 | 48,346,022 | 98,909,530 | 45,882,847 | 193,138,398 |
| 8 | 2028 | 49,512,141 | 100,805,012 | 47,556,252 | 197,873,405 |
| 9 | 2029 | 50,674,503 | 102,716,026 | 49,217,882 | 202,608,411 |
| 10 | 2030 | 51,852,465 | 104,654,581 | 50,836,372 | 207,343,417 |
| 11 | 2031 | 52,379,560 | 105,779,092 | 51,701,072 | 209,859,723 |
| 12 | 2032 | 52,901,112 | 106,955,631 | 52,519,284 | 212,376,028 |
| 13 | 2033 | 53,419,886 | 108,156,381 | 53,316,068 | 214,892,334 |
| 14 | 2034 | 53,948,078 | 109,334,271 | 54,126,290 | 217,408,639 |
| 15 | 2035 | 54,442,576 | 110,480,091 | 55,002,279 | 219,924,946 |
| 16 | 2036 | 54,932,204 | 111,668,157 | 55,840,890 | 222,441,251 |
| 17 | 2037 | 55,417,818 | 112,966,069 | 56,573,669 | 224,957,557 |
| 18 | 2038 | 55,900,599 | 114,351,167 | 57,222,096 | 227,473,862 |
| 19 | 2039 | 56,381,947 | 115,776,790 | 57,831,431 | 229,990,168 |
| 20 | 2040 | 56,863,049 | 117,170,416 | 58,473,009 | 232,506,474 |
| 21 | 2041 | 58,072,049 | 119,656,172 | 59,790,596 | 237,518,818 |
| 22 | 2042 | 59,281,924 | 122,107,550 | 61,141,687 | 242,531,162 |
| 23 | 2043 | 60,493,547 | 124,551,126 | 62,498,833 | 247,543,506 |
| 24 | 2044 | 61,707,462 | 126,968,486 | 63,879,902 | 252,555,850 |
| 25 | 2045 | 62,924,210 | 129,280,127 | 65,363,858 | 257,568,194 |
| 26 | 2046 | 64,144,523 | 131,623,590 | 66,812,425 | 262,580,537 |
| 27 | 2047 | 65,369,120 | 133,944,033 | 68,279,729 | 267,592,882 |
| 28 | 2048 | 66,598,307 | 136,256,791 | 69,750,129 | 272,605,226 |
| 29 | 2049 | 67,832,064 | 138,587,786 | 71,197,722 | 277,617,571 |
| 30 | 2050 | 69,070,083 | 140,847,200 | 72,712,632 | 282,629,915 |

The data was obtained using **Etaware-Vac-2021** U.S. Influenza Prediction model (Supplementary file “S7”).
© P. M. Etaware

It is expected that in 2050, the U.S. population will almost or totally achieve herd immunity against influenza infection with an average estimated value of 70.7% of the total population immune against the disease. It is also expected that 85.9% of the total population of infants and youths below 18 years will be immune against influenza infection, 60.9% of the population of young adults (age between 18 and 64 years) will be immune against the infection, while 82.5% of senior citizens of the United States will have acquired immunity against influenza infection in 2050, provided that there are potent influenza vaccines available or in circulation for mass vaccination and the perception of the populace towards the reception of vaccines increase with the current awareness campaign against respiratory diseases (Table 6).

**Table 6.** Projected level of acquired immunity against influenza infection in the U.S.

| Projected Acquired Immunity (%) / Age group |  |  |  |  |  |
| --- | --- | --- | --- | --- | --- |
| No. | Year | <18years | 18-64yrs | ≥65years | Total |
| 1 | 2021 | 58.1 | 44.7 | 64.0 | 51.0 |
| 2 | 2022 | 59.5 | 45.5 | 64.8 | 52.0 |
| 3 | 2023 | 60.8 | 46.3 | 65.6 | 53.0 |
| 4 | 2024 | 62.1 | 47.1 | 66.3 | 54.0 |
| 5 | 2025 | 63.5 | 47.9 | 67.0 | 54.9 |
| 6 | 2026 | 64.8 | 48.7 | 67.7 | 55.8 |
| 7 | 2027 | 66.1 | 49.4 | 68.5 | 56.8 |
| 8 | 2028 | 67.3 | 50.2 | 69.2 | 57.7 |
| 9 | 2029 | 68.5 | 50.9 | 70.0 | 58.5 |
| 10 | 2030 | 69.7 | 51.6 | 70.7 | 59.4 |
| 11 | 2031 | 70.2 | 52.0 | 70.9 | 59.8 |
| 12 | 2032 | 70.7 | 52.3 | 71.0 | 60.1 |
| 13 | 2033 | 71.1 | 52.7 | 71.2 | 60.5 |
| 14 | 2034 | 71.6 | 53.0 | 71.4 | 60.8 |
| 15 | 2035 | 72.1 | 53.3 | 71.6 | 61.2 |
| 16 | 2036 | 72.6 | 53.7 | 71.7 | 61.5 |
| 17 | 2037 | 73.0 | 54.0 | 72.0 | 61.9 |
| 18 | 2038 | 73.5 | 54.3 | 72.3 | 62.2 |
| 19 | 2039 | 74.0 | 54.6 | 72.7 | 62.5 |
| 20 | 2040 | 74.4 | 54.9 | 73.0 | 62.8 |
| 21 | 2041 | 75.6 | 55.5 | 74.1 | 63.7 |
| 22 | 2042 | 76.9 | 56.1 | 75.2 | 64.5 |
| 23 | 2043 | 78.0 | 56.7 | 76.3 | 65.3 |
| 24 | 2044 | 79.2 | 57.3 | 77.3 | 66.1 |
| 25 | 2045 | 80.4 | 58.0 | 78.2 | 66.9 |
| 26 | 2046 | 81.5 | 58.5 | 79.1 | 67.7 |
| 27 | 2047 | 82.6 | 59.1 | 80.0 | 68.4 |
| 28 | 2048 | 83.7 | 59.7 | 80.8 | 69.2 |
| 29 | 2049 | 84.8 | 60.3 | 81.7 | 69.9 |
| 30 | 2050 | 85.9 | 60.9 | 82.5 | 70.7 |

**Note:** The A.I. was calculated as a percentage of each vaccinated proportion in relation to the total population and that of the population of each age distribution presented in Supplementary file “S7”.

Due to the anticipated increase in the immunity level of the U.S. population, it is expected that in 2050, the susceptibility of humans to influenza infection in the United States will drastically reduce by almost half of the value calculated in 2021 i.e. from 0.49 to 0.29, while that of infants and young adults below the age of 18 years will be reduced by two-third of the value recorded in 2021 (0.42 in 2021 to 0.14 in 2050) as shown in Table 7, if only strict compliance to influenza vaccination is upheld.

**Table 7.** Estimated susceptibility levels to influenza infection in the U.S.

| Estimated Susceptibility Levels/Age group |  |  |  |  |  |
| --- | --- | --- | --- | --- | --- |
| No. | Year | <18years | 18-64yrs | ≥65years | Total |
| 1 | 2021 | 0.42 | 0.55 | 0.36 | 0.49 |
| 2 | 2022 | 0.41 | 0.54 | 0.35 | 0.48 |
| 3 | 2023 | 0.39 | 0.54 | 0.34 | 0.47 |
| 4 | 2024 | 0.38 | 0.53 | 0.34 | 0.46 |
| 5 | 2025 | 0.37 | 0.52 | 0.33 | 0.45 |
| 6 | 2026 | 0.35 | 0.51 | 0.32 | 0.44 |
| 7 | 2027 | 0.34 | 0.51 | 0.32 | 0.43 |
| 8 | 2028 | 0.33 | 0.50 | 0.31 | 0.42 |
| 9 | 2029 | 0.31 | 0.49 | 0.30 | 0.41 |
| 10 | 2030 | 0.30 | 0.48 | 0.29 | 0.41 |
| 11 | 2031 | 0.30 | 0.48 | 0.29 | 0.40 |
| 12 | 2032 | 0.29 | 0.48 | 0.29 | 0.40 |
| 13 | 2033 | 0.29 | 0.47 | 0.29 | 0.40 |
| 14 | 2034 | 0.28 | 0.47 | 0.29 | 0.39 |
| 15 | 2035 | 0.28 | 0.47 | 0.28 | 0.39 |
| 16 | 2036 | 0.27 | 0.46 | 0.28 | 0.38 |
| 17 | 2037 | 0.27 | 0.46 | 0.28 | 0.38 |
| 18 | 2038 | 0.27 | 0.46 | 0.28 | 0.38 |
| 19 | 2039 | 0.26 | 0.45 | 0.27 | 0.37 |
| 20 | 2040 | 0.26 | 0.45 | 0.27 | 0.37 |
| 21 | 2041 | 0.24 | 0.44 | 0.26 | 0.36 |
| 22 | 2042 | 0.23 | 0.44 | 0.25 | 0.35 |
| 23 | 2043 | 0.22 | 0.43 | 0.24 | 0.35 |
| 24 | 2044 | 0.21 | 0.43 | 0.23 | 0.34 |
| 25 | 2045 | 0.20 | 0.42 | 0.22 | 0.33 |
| 26 | 2046 | 0.18 | 0.41 | 0.21 | 0.32 |
| 27 | 2047 | 0.17 | 0.41 | 0.20 | 0.32 |
| 28 | 2048 | 0.16 | 0.40 | 0.19 | 0.31 |
| 29 | 2049 | 0.15 | 0.40 | 0.18 | 0.30 |
| 30 | 2050 | 0.14 | 0.39 | 0.17 | 0.29 |

The Susceptibility index of U.S. citizens to Influenza infection was calculated from the A. I. (%)

It was predicted that 49,734,427 individuals will be infected in 2021 out of the current estimation of 322,900.000 U.S. citizens i.e. 1.5% of the total population of U.S. citizens in 2021 (Table 8). The situation was estimated by Etaware-Pred-2021 to aggravate as the ratio of infected individuals in the population is expected to increase geometrically from 1.5% (in 2021) to 75.2% (in 2050) i.e. 300,803,433 infected individuals out of the estimated 400,000,000 citizens (if necessary health actions are not taken on time). It is also expected that the influenza infection rate among U.S. citizens will increase drastically from 9.6% in 2021 (96 persons out of every 1,000 individuals encountered in the United States) to 13.4% in 2050 i.e. 134 infected individuals out of every 1,000 U.S. citizens (Table 8).

**Table 8.** Prediction of future influenza outbreak per estimated population of the U.S.

| No. | Year | Infection Rate (%) | Influenza Outbreak | Total Population (N) |
| --- | --- | --- | --- | --- |
| 1 | 2021 | 9.6 | 49,734,427 | 322,900,000 |
| 2 | 2022 | 9.8 | 57,265,679 | 325,800,000 |
| 3 | 2023 | 10.0 | 71,451,426 | 328,700,000 |
| 4 | 2024 | 10.2 | 82,159,990 | 331,600,000 |
| 5 | 2025 | 10.4 | 94,208,216 | 334,500,000 |
| 6 | 2026 | 10.5 | 105,319,110 | 337,400,000 |
| 7 | 2027 | 10.7 | 116,681,727 | 340,300,000 |
| 8 | 2028 | 10.9 | 127,640,905 | 343,200,000 |
| 9 | 2029 | 11.1 | 138,460,651 | 346,100,000 |
| 10 | 2030 | 11.2 | 149,139,073 | 349,000,000 |
| 11 | 2031 | 11.3 | 152,857,126 | 351,100,000 |
| 12 | 2032 | 11.4 | 159,647,922 | 353,200,000 |
| 13 | 2033 | 11.4 | 164,924,256 | 355,300,000 |
| 14 | 2034 | 11.5 | 170,830,119 | 357,400,000 |
| 15 | 2035 | 11.6 | 176,398,057 | 359,500,000 |
| 16 | 2036 | 11.6 | 182,032,446 | 361,600,000 |
| 17 | 2037 | 11.7 | 187,601,790 | 363,700,000 |
| 18 | 2038 | 11.8 | 193,108,677 | 365,800,000 |
| 19 | 2039 | 11.8 | 198,555,214 | 367,900,000 |
| 20 | 2040 | 11.9 | 203,970,019 | 370,000,000 |
| 21 | 2041 | 12.0 | 216,394,346 | 373,000,000 |
| 22 | 2042 | 12.2 | 225,344,585 | 376,000,000 |
| 23 | 2043 | 12.3 | 235,723,399 | 379,000,000 |
| 24 | 2044 | 12.5 | 245,208,287 | 382,000,000 |
| 25 | 2045 | 12.6 | 254,920,228 | 385,000,000 |
| 26 | 2046 | 12.8 | 264,325,206 | 388,000,000 |
| 27 | 2047 | 12.9 | 273,624,983 | 391,000,000 |
| 28 | 2048 | 13.1 | 282,817,289 | 394,000,000 |
| 29 | 2049 | 13.2 | 291,845,800 | 397,000,000 |
| 30 | 2050 | 13.4 | 300,803,433 | 400,000,000 |

The data was obtained using **Etaware-Inra-2021** and **Etaware-Pred-2021** Influenza forecast models. (Supplementary file “S7”). © P. M. Etaware

The number of U.S. citizens vaccinated against influenza infection annually and the rate at which influenza and pneumonia kill infected individuals annually were used as determinants (predictors) for the development of another model for the estimation of deaths associated with influenza and pneumonia in the United States. The developed model was christened Etaware-Mort-2021 (Fig 9).

**Fig 9.**
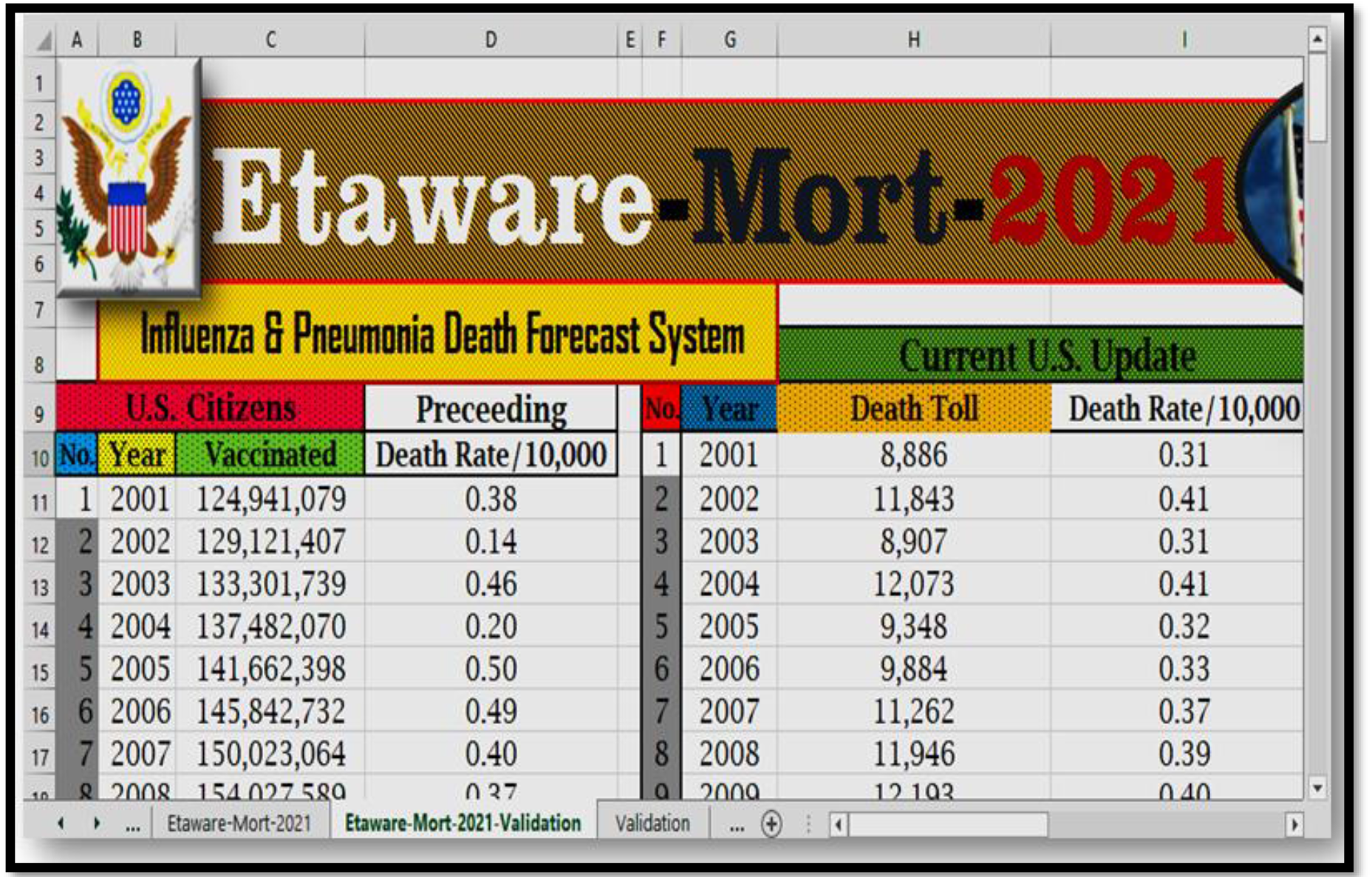
Etaware-Mort-2021 forecast system for flu and pneumonia related deaths in the U.S.

Etaware-Mort-2021 was structured using the regression equation described below (Data available in Supplementary File “S8 and S9”):

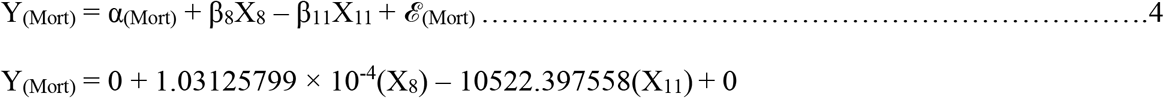

Y_(Mort)_ = 0 + 1.03125799 × 10^−4^(X_8_) – 10522.397558(X_11_) + 0

Y_(Mort)_ = Deaths associated with influenza and pneumonia in the U.S.

X_8_ = Total number of vaccinated individuals in the U.S.

X_11_ = Previous recorded mortality rate per 10,000 individuals in the U.S.

α_(Mort)_ = 0 (Suppressed)

*ℰ*_(Mort)_ = 0 (Suppressed)

The model statistics for Etaware-Mort-2021 was stated thus:

▪ R^2^ = 0.89
▪ MSEP = 6673436.10
▪ Adj. R^2^ = 0.88
▪ Pre R^2^ = 0.51

Model Validation

▪ Method: Bootstrap
▪ Validate N Times: 10
▪ LGO Pre R^2^ = 0.55

The relation trend between the selected variables and deaths associated with influenza and pneumonia in the United States was described in Fig 10.

**Fig 10.**
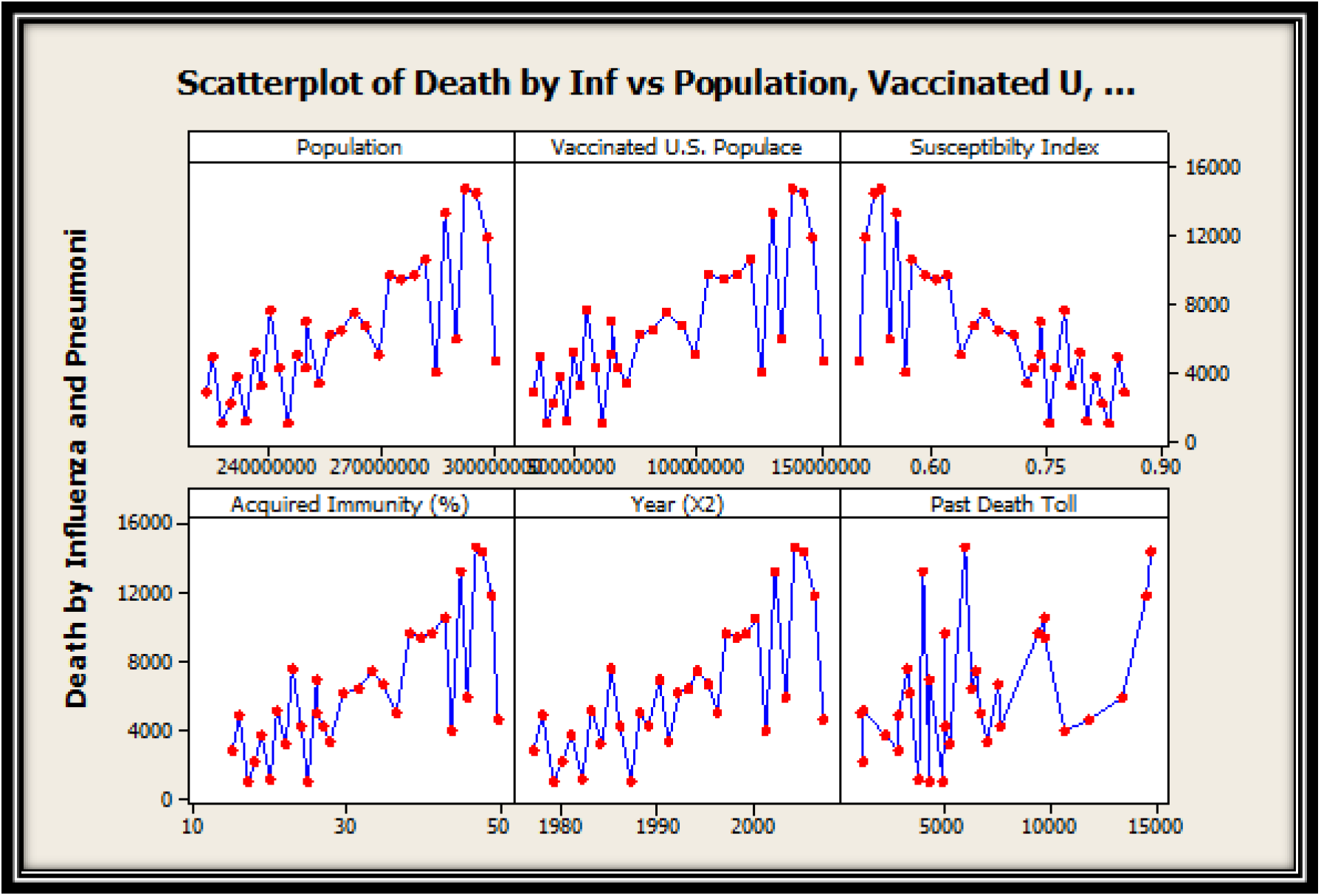
Predictors versus influenza and pneumonia associated deaths in the U.S.

It was estimated that the death toll from influenza and pneumonia in the United States will increase arithmetically from 12,909 casualties in 2021 to 23,133 deaths in 2050 as shown in Fig 11. Also, it is expected that the rate of death from the disease will increase from 40 persons in 2021 to 58 persons in 2050 per unit population size of 1,000,000 individuals encountered or censored in the United States (Fig 12). Fortunately, there is a decline in the death risk associated with contracting the influenza virus (Fig 13) i.e. 26 deaths out of 100,000 infected individuals (2021) compared to 8 deaths per 100,000 infected individuals in the United States (2050).

**Fig 11.**
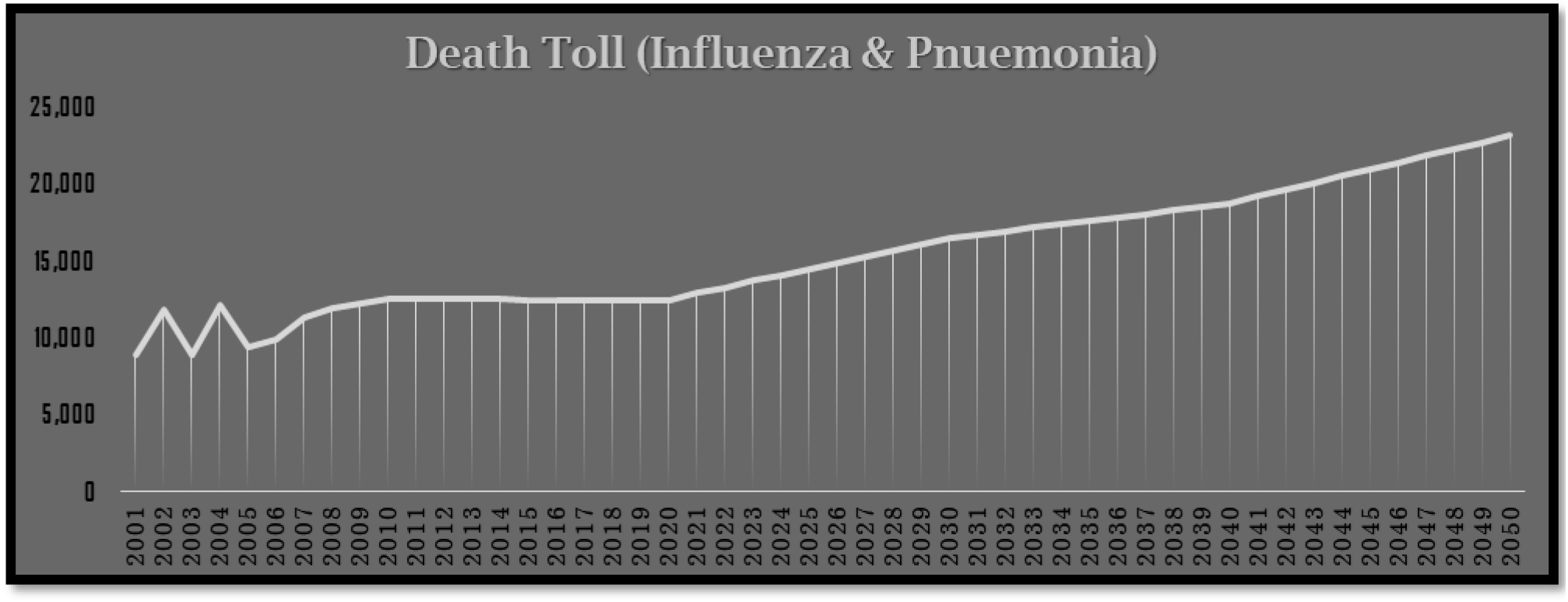
Estimated deaths associated with influenza and pneumonia in the U.S. (2021 -2050)

**Fig 12.**
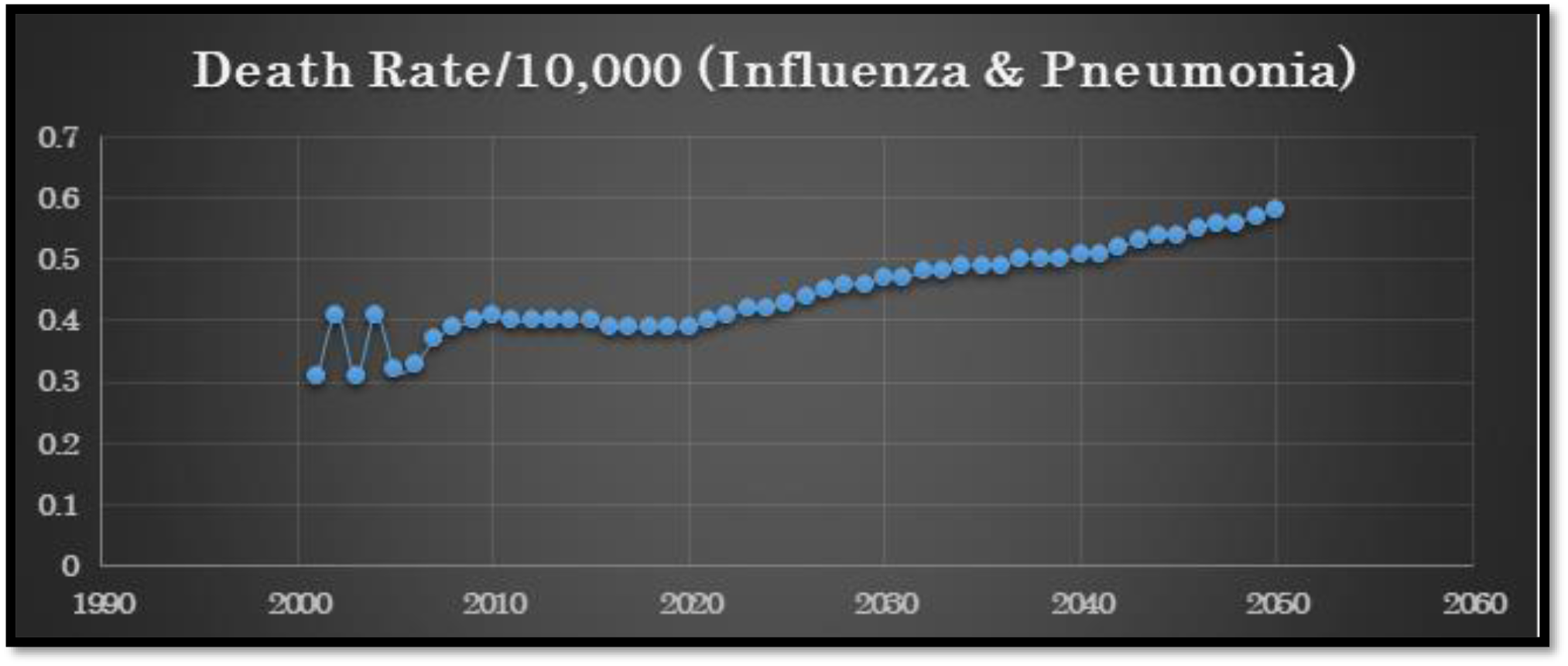
Estimated death rate for influenza and pneumonia (per 10,000 persons) in the U.S.

**Fig 13.**
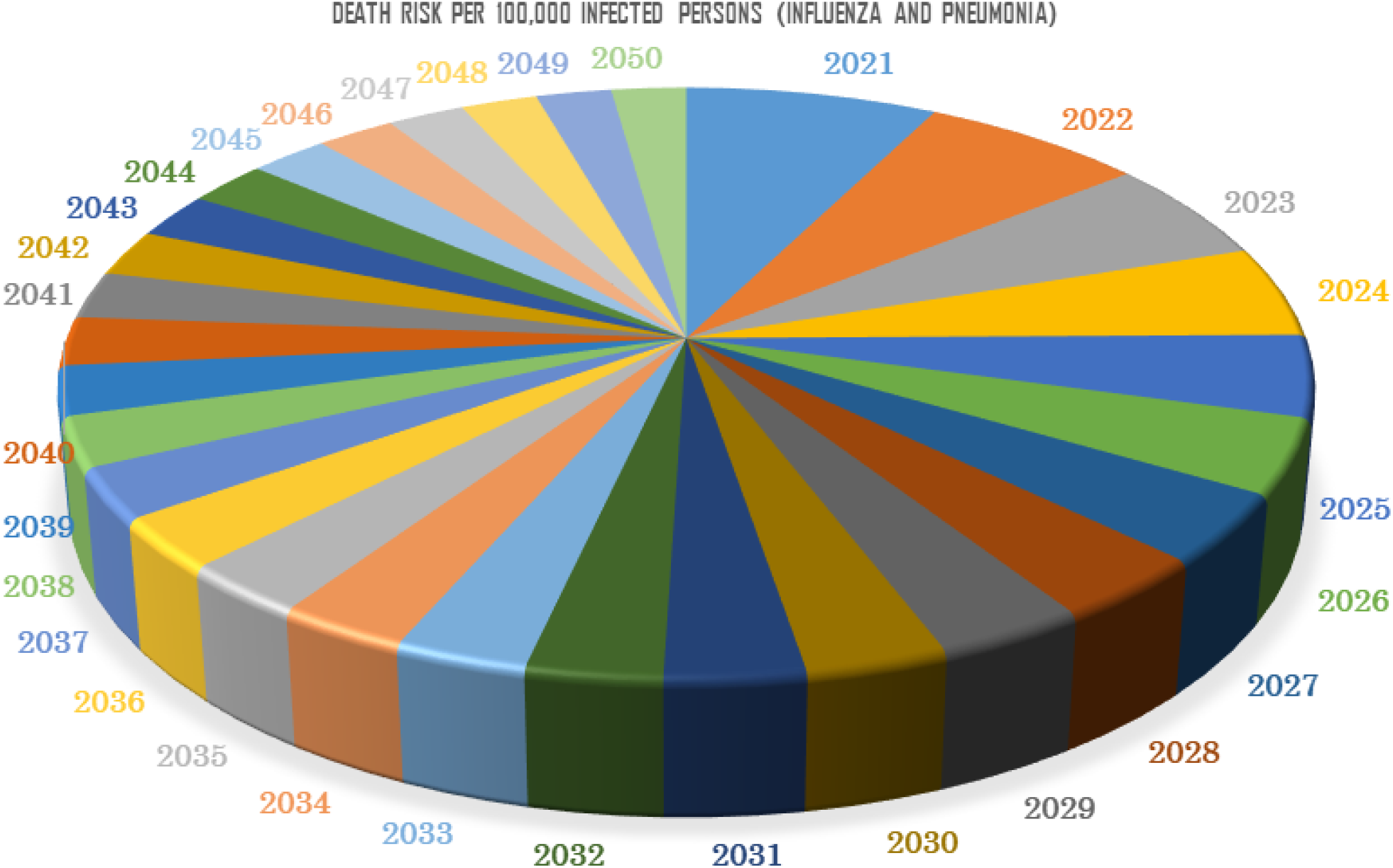
Influenza and pneumonia mortality risk (per 100,000 persons) in the U.S.

## 4.0 Discussion

The estimated amount of U.S. citizens to be vaccinated against influenza infection over the years will tend to increase appreciably till 2050, in direct correlation with the projected population size. This might be due to the fact that numerous awareness programs sponsored by the U.S. government and other health official to sensitize their citizens on the dangers associated with contracting the deadly influenza virus and other respiratory infections are already in place. Even though these figures are encouraging, in actual sense, the ratio of vaccinated individuals to that of the total population is what is most important, if the United States have to achieve herd immunity against the disease. Sadly, there are numerous setbacks to achieving herd immunity in the United States such as the perception or predisposition of U.S. citizens to the reception of all manner of vaccines, or other logistics like shortfall in the development and supply of potent vaccines, the pressure of managing multiple jobs for survival, academic, research or study leaves, excessive travel-schedule for business or other functions, immunocompromised or incapacitated individuals etc. These are some of the reasons that will inhibit the progress of vaccination against influenza and indeed all other forms of transmittable diseases in the United States. Fortunately, it is expected that the immunity level of the U.S. populace to influenza infection will increase while the susceptibility of humans to the influenza virus will drastically reduce. The observation noted in this research was in line with the agitations and aims of WHO i.e. to eradicate influenza infections and other forms of transmissible ailments in the nearest future.

The influenza outbreak estimation in the United States made by Etaware-Pred-2021 showed that as the population of the United States’ citizens increase geometrically through natality and migration, so also, the number of influenza infections in the U.S. will increase, unless there are potent influenza vaccines in circulation, strict compliance of U.S. citizens to influenza vaccination and a high level of personal hygiene among the citizens of the United States before the situation predicted by Etaware-Pred-2021 can be averted. It is also expected that the spread of the disease from human to human will increase over the years regardless of the chemistry of the pathogen (viral chemistry). It was further estimated that the death toll from influenza and pneumonia in the United States will increase arithmetically from 2021 to 2050. The observations raised by this research have been a major crux for concern to the U.S. government, the Centre of Disease Control of the United States, Health and Medical practitioners in the U.S., World Health Organization and the global health community in general.

## 5.0 Conclusion

It is expected that the rate of influenza infections in the United States will increase in the nearest future due to constant mutation and adaptation of the pathogen to the human body system, and a flawless adaptation of influenza virus to extremely low temperatures. unless there are potent influenza vaccines in circulation, strict compliance of U.S. citizens to influenza vaccination and a high level of personal hygiene among the citizens of the United States, influenza infection will become a major threat to the peaceful existence of humanity worldwide. These situations can be averted if the U.S. citizens act accordingly. Finally, all the itemized predictors stated initially had significant effects on influenza outbreak and mortality in the United States.

## Data Availability

the data used for this research are available for review. The data were submitted alongside the manuscript

## Ethical Statement

This is to confirm that:

Dr. P. M. Etaware declare that he has no conflict of interest in the publication of this research article.

Thank you

Peter M. Etaware (Ph.D.)

## Ethical Approval

‘Not applicable’. No human or animal specimens was subjected to laboratory analysis.

## Consent to Participate

The author gave his consent to participate in this research.

## Consent for Publication

The author gave his approval for the publication of this article

## Availability of Data and Material

All data and materials used in this research are available.

## Authors’ Contributions

P. M. E conceptualized, designed the experiment, conducted the research, wrote the draft manuscript, reviewed the manuscript and approved the final version of the manuscript.

**Supplementary file “S1”. Raw data on influenza vaccination history in the United States**

**Supplementary file “S2”. Statistical analysis of data obtained in the U.S. on Influenza vaccination**

**Supplementary file “S3”. Data obtained for influenza outbreak and infection rate in the U.S**.

**Supplementary file “S4”. Analyzed data on influenza infection rate in the U.S**.

**Supplementary file “S5”. Analyzed data on influenza outbreak in the U.S**.

**Supplementary file “S6”. Post-Analysis data**

**Supplementary file “S7”. The predictions made by the developed models (commands, formula and functions erased)**

**Supplementary file “S8”. Raw data on deaths associated with influenza and pneumonia in the United States**

**Supplementary file “S9”. Analyzed data on deaths associated with influenza and pneumonia in the United States**

