## Supplementary File (S2) for "Influenza outbreak, disease transmission rate and mortality risk in the United States (2021 to 2050 Projection)"

Analysis

REGRESSION: MULTIPLE (ONE SUBSET)

2021-03-17 08:41:19

Using: C:\Users\LENOVO\Desktop\Influenza-PLOS ONE\Data Influenza PLOS ONE\Medical Data\Vaccination\Model-Vaccination.dt

X Columns:

1) Year	4) % 65yrs Po	5) Population	6) % 65yrs Po

Y Column: 3) Vaccinated

Keep If:

Calculate Constant: false

Total number of data points = 35

Number of data points used = 35

Regression equation:

col(3)[Vaccinated] =

-11199.859025*col(1)[Year]

+1.24133297571*col(4)[% 65yrs Po]

+0.20342302602*col(5)[Population]

-4470313.8705*col(6)[% 65yrs Po]

R^2	= 0.98711114318	AIC = 1026.33181641	MSEP	= 5.52352118e12

adj R^2 = 0.98544806488	BIC = 1029.33077583	PRESS	= 1.96218539e14

PRE R^2 = 0.94693612479	MAE = 1789784.01831	LOO MAE = 2018674.16209

For each term in the ANOVA table below, if P<=0.05, that term was a

significant source of Y's variation.

Source	SS	df	MS	F	P

------------------------ ------------- -------- --------- --------- ---------

Regression	1.15905016e16	4 2.8976e15 593.5446 .0000 ***

col(1)[Year]	8.11787467e15	1 8.1179e15 1662.8515 .0000 ***

col(4)[% 65yrs Po]	3.27690338e15	1 3.2769e15 671.23525 .0000 ***

col(5)[Population]	1.79256985e14	1 1.7926e14 36.718692 .0000 ***

col(6)[% 65yrs Po]	1.64665668e13	1 1.6467e13 3.3729832 .0759 ns

Error	1.51338901e14	31 4.8819e12

------------------------ ------------- -------- --------- --------- ---------

Total	1.17418405e16	35

Table of Statistics for the Regression Coefficients:

Column	Coef. Std Error t(Coef=0)	P	+/-95% CL

------------------------ --------- --------- --------- --------- ---------

col(1)[Year]	-11199.86 13690.575 -0.818071 .4196 ns	27922.112

col(4)[% 65yrs Po]	1.241333 0.7288433 1.7031547 .0985 ns	1.4864858

col(5)[Population]	0.203423 0.0908443 2.2392494 .0325 *	0.1852782

col(6)[% 65yrs Po]	-4470314	2434058 -1.836568 .0759 ns	4964294.1

Degrees of freedom for two-tailed t tests = 31

If P<=0.05, the coefficient is significantly different from 0.

Residuals:

Row	Y observed	Y expected	Residual

--------- ------------- ------------- -------------

1	3442860.8 3682715.47175 -239854.67175

2	3690953.47 4305701.09324 -614747.62324

3	5301766.36 3187803.29239 2113963.06761

4	5107096.47 3858302.38296 1248794.08704

5	9327070.74 4543780.21929 4783290.52071

6	5698149.49 5242314.11084 455835.379158

7	5501611.5 5950495.57231 -448884.07231

8	4932818.57 6157127.29991 -1224308.7299

9	5293760 6707992.05926 -1414232.0593

10	5677757.61 7238001.15382 -1560243.5438

11	5800749.15 7777453.38921 -1976704.2392

12	6196298.68 8363738.55514 -2167439.8751

13	6606484.12 8956612.41173 -2350128.2917

14	7038078.72 9542648.62314 -2504569.9031

15	7491030.5 10129417.8988 -2638387.3988

16	8533731.08 10812055.1047 -2278324.0247

17	9623351.08 11488392.9533 -1865041.8733

18	11578962.78 11456438.3787 122524.401333

19	13372451.4 12517893.0392 854558.360844

20	15523961.28 13606920.9973 1917040.28269

21	17087971.16 14691846.0456 2396125.11444

22	18309241.5 15823864.7719 2485376.72812

23	19565718.84 16949615.4157	2616103.4243

24	21153423.5 18102101.2064 3051322.29357

25	21653355.15 19306695.8209 2346659.32915

26	21789979.47	20518808.885 1271170.58503

27	21903771.33 21737082.8907 166688.439301

28	22782098.3 22912628.1609 -130529.86086

29	22917039.6 23909323.2296 -992283.62962

30	31174913.667 32992509.4365 -1817595.7695

31	30484988.452 33201172.4116 -2716183.9596

32	32297014.973 33416132.3156 -1119117.3426

33	30363003.056 33624823.5274 -3261820.4714

34	35766203.763 33831774.0444 1934429.71864

35	37637420.588 34079259.0191 3558161.56894

Validation Method: Bootstrap

Validate N Times: 10

Leave-Group-Out PRESS	= 8.12610888e13

Leave-Group-Out PRE R^2 = 0.9364989

Leave-Group-Out MAE	= 2215474.37562

(The validation method randomly assigns rows of data to validation groups,

so the Leave-Group-Out statistics printed above will vary.

You can reduce the variability by increasing 'Validate N Times'.)

Group Leave-Group-Out Validation Equations

----- ----------------------------------------------------------------------

1 -12416.863638*col(1)[Year] +1.27463171797*col(4)[% 65yrs Po] +0.22228079317*col(5)[Population] -4786621.3075*col(6)[% 65yrs Po]

2 -21720.41709*col(1)[Year] +0.6811682085*col(4)[% 65yrs Po] +0.26756078366*col(5)[Population] -2656003.1057*col(6)[% 65yrs Po]

3 -3884.3382121*col(1)[Year] +1.66530312182*col(4)[% 65yrs Po] +0.12789224265*col(5)[Population] -5200926.8978*col(6)[% 65yrs Po]

4 -14490.773284*col(1)[Year] +0.97357777483*col(4)[% 65yrs Po] +0.19902063862*col(5)[Population] -3123454.0586*col(6)[% 65yrs Po]

5 -15980.365792*col(1)[Year] +1.05536735812*col(4)[% 65yrs Po] +0.25834463377*col(5)[Population] -4383411.1032*col(6)[% 65yrs Po]

6 -25394.508187*col(1)[Year] +0.44337710577*col(4)[% 65yrs Po] +0.27844480122*col(5)[Population] -1634591.1307*col(6)[% 65yrs Po]

7 4993.1361531*col(1)[Year] +2.25555662885*col(4)[% 65yrs Po] +0.09742137674*col(5)[Population] -7509259.9357*col(6)[% 65yrs Po]

8 13896.1375779*col(1)[Year] +2.65106078375*col(4)[% 65yrs Po] +0.03921175741*col(5)[Population] -8846921.4634*col(6)[% 65yrs Po]

9 -11396.566734*col(1)[Year] +1.41914333289*col(4)[% 65yrs Po] +0.18677873154*col(5)[Population] -4574128.407*col(6)[% 65yrs Po]

10 -14860.982644*col(1)[Year] +1.12887035574*col(4)[% 65yrs Po] +0.25580023039*col(5)[Population] -4655864.2037*col(6)[% 65yrs Po]

(The validation method randomly assigns rows of data to validation groups,

so the LGO Validation Equations will vary. Since these equations are

generated during the individual validation runs, increasing

'Validate N Times' will not decrease the variability of the coefficients.)

REGRESSION: MULTIPLE (ONE SUBSET)

2021-03-17 08:41:19

Using: C:\Users\LENOVO\Desktop\Influenza-PLOS ONE\Data Influenza PLOS ONE\Medical Data\Vaccination\Model-Vaccination.dt

X Columns:

1) Year	4) % 65yrs Po	5) Population	6) % 65yrs Po

Y Column: 3) Vaccinated

Keep If:

Calculate Constant: false

Total number of data points = 35

Number of data points used = 35

Regression equation:

col(3)[Vaccinated] =

-11199.859025*col(1)[Year]

+1.24133297571*col(4)[% 65yrs Po]

+0.20342302602*col(5)[Population]

-4470313.8705*col(6)[% 65yrs Po]

R^2	= 0.98711114318	AIC = 1026.33181641	MSEP	= 5.52352118e12

adj R^2 = 0.98544806488	BIC = 1029.33077583	PRESS	= 1.96218539e14

PRE R^2 = 0.94693612479	MAE = 1789784.01831	LOO MAE = 2018674.16209

For each term in the ANOVA table below, if P<=0.05, that term was a

significant source of Y's variation.

Source	SS	df	MS	F	P

------------------------ ------------- -------- --------- --------- ---------

Regression	1.15905016e16	4 2.8976e15 593.5446 .0000 ***

col(1)[Year]	8.11787467e15	1 8.1179e15 1662.8515 .0000 ***

col(4)[% 65yrs Po]	3.27690338e15	1 3.2769e15 671.23525 .0000 ***

col(5)[Population]	1.79256985e14	1 1.7926e14 36.718692 .0000 ***

col(6)[% 65yrs Po]	1.64665668e13	1 1.6467e13 3.3729832 .0759 ns

Error	1.51338901e14	31 4.8819e12

------------------------ ------------- -------- --------- --------- ---------

Total	1.17418405e16	35

Table of Statistics for the Regression Coefficients:

Column	Coef. Std Error t(Coef=0)	P	+/-95% CL

------------------------ --------- --------- --------- --------- ---------

col(1)[Year]	-11199.86 13690.575 -0.818071 .4196 ns	27922.112

col(4)[% 65yrs Po]	1.241333 0.7288433 1.7031547 .0985 ns	1.4864858

col(5)[Population]	0.203423 0.0908443 2.2392494 .0325 *	0.1852782

col(6)[% 65yrs Po]	-4470314	2434058 -1.836568 .0759 ns	4964294.1

Degrees of freedom for two-tailed t tests = 31

If P<=0.05, the coefficient is significantly different from 0.

Residuals:

Row	Y observed	Y expected	Residual

--------- ------------- ------------- -------------

1	3442860.8 3682715.47175 -239854.67175

2	3690953.47 4305701.09324 -614747.62324

3	5301766.36 3187803.29239 2113963.06761

4	5107096.47 3858302.38296 1248794.08704

5	9327070.74 4543780.21929 4783290.52071

6	5698149.49 5242314.11084 455835.379158

7	5501611.5 5950495.57231 -448884.07231

8	4932818.57 6157127.29991 -1224308.7299

9	5293760 6707992.05926 -1414232.0593

10	5677757.61 7238001.15382 -1560243.5438

11	5800749.15 7777453.38921 -1976704.2392

12	6196298.68 8363738.55514 -2167439.8751

13	6606484.12 8956612.41173 -2350128.2917

14	7038078.72 9542648.62314 -2504569.9031

15	7491030.5 10129417.8988 -2638387.3988

16	8533731.08 10812055.1047 -2278324.0247

17	9623351.08 11488392.9533 -1865041.8733

18	11578962.78 11456438.3787 122524.401333

19	13372451.4 12517893.0392 854558.360844

20	15523961.28 13606920.9973 1917040.28269

21	17087971.16 14691846.0456 2396125.11444

22	18309241.5 15823864.7719 2485376.72812

23	19565718.84 16949615.4157	2616103.4243

24	21153423.5 18102101.2064 3051322.29357

25	21653355.15 19306695.8209 2346659.32915

26	21789979.47	20518808.885 1271170.58503

27	21903771.33 21737082.8907 166688.439301

28	22782098.3 22912628.1609 -130529.86086

29	22917039.6 23909323.2296 -992283.62962

30	31174913.667 32992509.4365 -1817595.7695

31	30484988.452 33201172.4116 -2716183.9596

32	32297014.973 33416132.3156 -1119117.3426

33	30363003.056 33624823.5274 -3261820.4714

34	35766203.763 33831774.0444 1934429.71864

35	37637420.588 34079259.0191 3558161.56894

Validation Method: Bootstrap

Validate N Times: 10

Leave-Group-Out PRESS	= 8.12610888e13

Leave-Group-Out PRE R^2 = 0.9364989

Leave-Group-Out MAE	= 2215474.37562

(The validation method randomly assigns rows of data to validation groups,

so the Leave-Group-Out statistics printed above will vary.

You can reduce the variability by increasing 'Validate N Times'.)

Group Leave-Group-Out Validation Equations

----- ----------------------------------------------------------------------

1 -12416.863638*col(1)[Year] +1.27463171797*col(4)[% 65yrs Po] +0.22228079317*col(5)[Population] -4786621.3075*col(6)[% 65yrs Po]

2 -21720.41709*col(1)[Year] +0.6811682085*col(4)[% 65yrs Po] +0.26756078366*col(5)[Population] -2656003.1057*col(6)[% 65yrs Po]

3 -3884.3382121*col(1)[Year] +1.66530312182*col(4)[% 65yrs Po] +0.12789224265*col(5)[Population] -5200926.8978*col(6)[% 65yrs Po]

4 -14490.773284*col(1)[Year] +0.97357777483*col(4)[% 65yrs Po] +0.19902063862*col(5)[Population] -3123454.0586*col(6)[% 65yrs Po]

5 -15980.365792*col(1)[Year] +1.05536735812*col(4)[% 65yrs Po] +0.25834463377*col(5)[Population] -4383411.1032*col(6)[% 65yrs Po]

6 -25394.508187*col(1)[Year] +0.44337710577*col(4)[% 65yrs Po] +0.27844480122*col(5)[Population] -1634591.1307*col(6)[% 65yrs Po]

7 4993.1361531*col(1)[Year] +2.25555662885*col(4)[% 65yrs Po] +0.09742137674*col(5)[Population] -7509259.9357*col(6)[% 65yrs Po]

8 13896.1375779*col(1)[Year] +2.65106078375*col(4)[% 65yrs Po] +0.03921175741*col(5)[Population] -8846921.4634*col(6)[% 65yrs Po]

9 -11396.566734*col(1)[Year] +1.41914333289*col(4)[% 65yrs Po] +0.18677873154*col(5)[Population] -4574128.407*col(6)[% 65yrs Po]

10 -14860.982644*col(1)[Year] +1.12887035574*col(4)[% 65yrs Po] +0.25580023039*col(5)[Population] -4655864.2037*col(6)[% 65yrs Po]

(The validation method randomly assigns rows of data to validation groups,

so the LGO Validation Equations will vary. Since these equations are

generated during the individual validation runs, increasing

'Validate N Times' will not decrease the variability of the coefficients.)

REGRESSION: MULTIPLE (ONE SUBSET)

2021-03-17 08:43:00

Using: C:\Users\LENOVO\Desktop\Influenza-PLOS ONE\Data Influenza PLOS ONE\Medical Data\Vaccination\Model-Vaccination.dt

X Columns:

1) Year	4) % 65yrs Po	5) Population	6) % 65yrs Po

Y Column: 3) Vaccinated

Keep If:

Calculate Constant: true

Total number of data points = 35

Number of data points used = 35

Regression equation:

col(3)[Vaccinated] = 2043347652.98

-1102594.7429*col(1)[Year]

+0.91492631527*col(4)[% 65yrs Po]

+0.61948326576*col(5)[Population]

-1758058.2046*col(6)[% 65yrs Po]

R^2	= 0.95997612325	AIC = 1027.54522299	MSEP	= 5.77323643e12

adj R^2 = 0.95463960635	BIC = 1031.1563341	PRESS	= 1.97647046e14

PRE R^2 = 0.94663926781	MAE = 1762786.91359	LOO MAE = 2025693.20959

For each term in the ANOVA table below, if P<=0.05, that term was a

significant source of Y's variation.

Source	SS	df	MS	F	P

------------------------ ------------- -------- --------- --------- ---------

Regression	3.54920822e15	4 8.873e14 179.88814 .0000 ***

col(1)[Year]	3.48964046e15	1 3.4896e15 707.47604 .0000 ***

col(4)[% 65yrs Po]	6.37427797e12	1 6.3743e12 1.2922962 .2646 ns

col(5)[Population]	5.22846535e13	1 5.2285e13 10.599986 .0028 **

col(6)[% 65yrs Po]	908822947888	1 9.0882e11 0.1842512 .6708 ns

Error	1.47975631e14	30 4.9325e12

------------------------ ------------- -------- --------- --------- ---------

Total	3.69718385e15	34

Table of Statistics for the Regression Coefficients:

Column	Coef. Std Error t(Coef=0)	P	+/-95% CL

------------------------ --------- --------- --------- --------- ---------

Intercept	2.04335e9 2.47455e9 0.8257459 .4155 ns	5.0537e9

col(1)[Year]	-1102595 1321779.5 -0.834174 .4108 ns	2699434

col(4)[% 65yrs Po]	0.9149263 0.8324498 1.0990769 .2805 ns	1.7000893

col(5)[Population]	0.6194833 0.5120674 1.2097689 .2358 ns	1.0457812

col(6)[% 65yrs Po]	-1758058 4095699.4 -0.429245 .6708 ns	8364534.2

Degrees of freedom for two-tailed t tests = 30

If P<=0.05, the coefficient is significantly different from 0.

Residuals:

Row	Y observed	Y expected	Residual

--------- ------------- ------------- -------------

1	3442860.8 3334581.80499 108278.995006

2	3690953.47	3608515.8456 82437.6244035

3	5301766.36 4150316.94851 1151449.41149

4	5107096.47 4491700.80395 615395.666047

5	9327070.74	4864811.262	4462259.478

6	5698149.49 5265578.24311 432571.246888

7	5501611.5 5686779.46184 -185167.96184

8	4932818.57 6230232.13623 -1297413.5662

9	5293760 6729991.21504	-1436231.215

10	5677757.61	7261882.5102 -1584124.9002

11	5800749.15 7817166.23581 -2016417.0858

12	6196298.68 8382217.37713 -2185918.6971

13	6606484.12 8965722.57448 -2359238.4545

14	7038078.72 9575706.58496	-2537627.865

15	7491030.5 10216098.7417 -2725068.2417

16	8533731.08 10856815.2286 -2323084.1486

17	9623351.08 11535609.2911 -1912258.2111

18	11578962.78 10680069.7199 898893.060083

19	13372451.4 11958101.7085 1414349.69147

20	15523961.28 13228943.4271	2295017.8529

21	17087971.16 14505631.4606 2582339.69937

22	18309241.5 15761643.9298	2547597.5702

23	19565718.84	17022198.861 2543519.97903

24	21153423.5 18267645.9938 2885777.50625

25	21653355.15	19477065.43	2176289.72

26	21789979.47 20674456.1932 1115523.27678

27	21903771.33 21858803.8989	44967.431112

28	22782098.3 23069392.5764 -287294.27643

29	22917039.6 23908371.4164 -991331.81644

30	31174913.667 33082325.2041 -1907411.5371

31	30484988.452 33233415.2313 -2748426.7793

32	32297014.973 33400332.7375 -1103317.7645

33	30363003.056 33611441.5229 -3248438.4669

34	35766203.763 33860397.1572	1905806.6058

35	37637420.588	34051124.415 3586296.17295

Validation Method: Bootstrap

Validate N Times: 10

Leave-Group-Out PRESS	= 8.75056451e13

Leave-Group-Out PRE R^2 = 0.9509611

Leave-Group-Out MAE	= 2155399.77932

(The validation method randomly assigns rows of data to validation groups,

so the Leave-Group-Out statistics printed above will vary.

You can reduce the variability by increasing 'Validate N Times'.)

Group Leave-Group-Out Validation Equations

----- ----------------------------------------------------------------------

1 7383437458.67 -3930641.4996*col(1)[Year] +1.28830909671*col(4)[% 65yrs Po] +1.60568815027*col(5)[Population] +247396.53832*col(6)[% 65yrs Po]

2 1250878945.06 -684087.82086*col(1)[Year] +0.80957883718*col(4)[% 65yrs Po] +0.5315699147*col(5)[Population] -3002244.7894*col(6)[% 65yrs Po]

3 404319226.171 -204050.14741*col(1)[Year] +2.42325255971*col(4)[% 65yrs Po] +0.11449237966*col(5)[Population] -7371174.8953*col(6)[% 65yrs Po]

4 886006250.224 -477332.2109*col(1)[Year] +1.47921247466*col(4)[% 65yrs Po] +0.3221229931*col(5)[Population] -4171558.0256*col(6)[% 65yrs Po]

5 6194003714.85 -3315105.0031*col(1)[Year] +0.41141813635*col(4)[% 65yrs Po] +1.47990045969*col(5)[Population] +2287845.09914*col(6)[% 65yrs Po]

6 3928507917 -2102107.6538*col(1)[Year] +1.02649723176*col(4)[% 65yrs Po] +0.97642660597*col(5)[Population] -950014.72006*col(6)[% 65yrs Po]

7 3849161460.43 -2096494.2364*col(1)[Year] -0.9340762835*col(4)[% 65yrs Po] +1.16861170252*col(5)[Population] +5717897.3286*col(6)[% 65yrs Po]

8 2813218599.75 -1526569.2547*col(1)[Year] +0.08318918468*col(4)[% 65yrs Po] +0.84890285505*col(5)[Population] +1675187.63686*col(6)[% 65yrs Po]

9 1452441642.78 -798916.36682*col(1)[Year] +0.42721672398*col(4)[% 65yrs Po] +0.5516145675*col(5)[Population] -174255.86662*col(6)[% 65yrs Po]

10 2450912568.45 -1324803.0429*col(1)[Year] +0.58286545038*col(4)[% 65yrs Po] +0.76617341434*col(5)[Population] -1125618.0913*col(6)[% 65yrs Po]

(The validation method randomly assigns rows of data to validation groups,

so the LGO Validation Equations will vary. Since these equations are

generated during the individual validation runs, increasing

'Validate N Times' will not decrease the variability of the coefficients.)
