## Supplementary File (S4) for "Influenza outbreak, disease transmission rate and mortality risk in the United States (2021 to 2050 Projection)"

Analysis

REGRESSION: MULTIPLE (ONE SUBSET)

2021-03-18 23:22:30

Using: C:\Users\LENOVO\Desktop\Influenza-PLOS ONE\Data Influenza PLOS ONE\Medical Data\Model 1.dt

X Columns:

10) Acquired I

Y Column: 8) cIR

Keep If:

Calculate Constant: false

Total number of data points = 9

Number of data points used = 9

Regression equation:

col(8)[cIR] =

0.18906177141*col(10)[Acquired I]

R^2	= 0.90990838611	AIC = 21.9889569656	MSEP	= 11.5206234153

adj R^2 = 0.89864693438	BIC = 24.2077069656	PRESS	= 105.368769969

PRE R^2 = 0.95358678121	MAE = 2.17566615933	LOO MAE = 2.4487558884

For each term in the ANOVA table below, if P<=0.05, that term was a

significant source of Y's variation.

Source	SS	df	MS	F	P

------------------------ ------------- -------- --------- --------- ---------

Regression	837.764161682	1 837.76416 80.798498 .0000 ***

col(10)[Acquired I]	837.764161682	1 837.76416 80.798498 .0000 ***

Error	82.9484885899	8 10.368561

------------------------ ------------- -------- --------- --------- ---------

Total	920.712650272	9

Table of Statistics for the Regression Coefficients:

Column	Coef. Std Error t(Coef=0)	P	+/-95% CL

------------------------ --------- --------- --------- --------- ---------

col(10)[Acquired I]	0.1890618	0.021033 8.9887985 .0000 *** 0.0485023

Degrees of freedom for two-tailed t tests = 8

If P<=0.05, the coefficient is significantly different from 0.

Residuals:

Row	Y observed	Y expected	Residual

--------- ------------- ------------- -------------

1	2.988432 9.84493705251 -6.8565050525

2	10.886968 9.79469804611 1.09226995389

3	9.572431 9.74481163992 -0.1723806399

4	9.538951 9.69527424177 -0.1563232418

5	7.604563 9.64608225946 -2.0415192595

6	9.156931 9.59723191178 -0.4403009118

7	14.159849 9.54871960655 4.61112939345

8	11.288805 9.50054212971 1.78826287029

9	11.875	9.4526958891	2.4223041109

Validation Method: Bootstrap

Validate N Times: 10

Leave-Group-Out PRESS	= 23.50925546

Leave-Group-Out PRE R^2 = 0.651765

Leave-Group-Out MAE	= 2.20397693646

(The validation method randomly assigns rows of data to validation groups,

so the Leave-Group-Out statistics printed above will vary.

You can reduce the variability by increasing 'Validate N Times'.)

Group Leave-Group-Out Validation Equations

----- ----------------------------------------------------------------------

1 0.19677143947*col(10)[Acquired I]

2 0.19092315237*col(10)[Acquired I]

3 0.17405926725*col(10)[Acquired I]

4 0.20209258425*col(10)[Acquired I]

5 0.19390277609*col(10)[Acquired I]

6 0.16535257142*col(10)[Acquired I]

7 0.21036141828*col(10)[Acquired I]

8 0.17455140998*col(10)[Acquired I]

9 0.19882586545*col(10)[Acquired I]

10 0.17014295504*col(10)[Acquired I]

(The validation method randomly assigns rows of data to validation groups,

so the LGO Validation Equations will vary. Since these equations are

generated during the individual validation runs, increasing

'Validate N Times' will not decrease the variability of the coefficients.)

REGRESSION: MULTIPLE (ONE SUBSET)

2021-03-18 23:10:51

Using: C:\Users\LENOVO\Desktop\Influenza-PLOS ONE\Data Influenza PLOS ONE\Medical Data\Model 1.dt

X Columns:

5) pIVD	7) Vaccinated	10) Acquired I

6) pIR	9) Susceptibi

Y Column: 8) cIR

Keep If:

Calculate Constant: false

Total number of data points = 9

Number of data points used = 9

Regression equation:

col(8)[cIR] =

8.34201264e-6*col(5)[pIVD]

-26.502484246*col(6)[pIR]

+2.03597306e-4*col(7)[Vaccinated]

-22445.237562*col(9)[Susceptibi]

-426.83034907*col(10)[Acquired I]

R^2	= 0.96320646378	AIC = 21.9294128545	MSEP	= 21.9568444165

adj R^2 = 0.91721454351	BIC = 33.3044128545	PRESS	= 299.456592058

PRE R^2 = 0.0059770996	MAE = 1.68385299442	LOO MAE = 4.73143184106

For each term in the ANOVA table below, if P<=0.05, that term was a

significant source of Y's variation.

Source	SS	df	MS	F	P

------------------------ ------------- -------- --------- --------- ---------

Regression	886.83637603	5 177.36728 20.94295 .0057 **

col(5)[pIVD]	803.165229036	1 803.16523 94.835131 .0006 ***

col(6)[pIR]	0.77343968758	1 0.7734397 0.0913252 .7776 ns

col(7)[Vaccinated]	71.9838573998	1 71.983857 8.4996192 .0434 *

col(9)[Susceptibi]	5.77369488332	1 5.7736949 0.6817391 .4554 ns

col(10)[Acquired I]	5.14015502285	1 5.140155 0.6069327 .4795 ns

Error	33.8762742425	4 8.4690686

------------------------ ------------- -------- --------- --------- ---------

Total	920.712650272	9

Table of Statistics for the Regression Coefficients:

Column	Coef. Std Error t(Coef=0)	P	+/-95% CL

------------------------ --------- --------- --------- --------- ---------

col(5)[pIVD]	8.342e-6 1.6807e-5 0.4963287 .6457 ns	4.6665e-5

col(6)[pIR]	-26.50248 52.721736 -0.502686 .6416 ns	146.37901

col(7)[Vaccinated]	2.036e-4 2.6254e-4 0.7754903 .4813 ns	7.2893e-4

col(9)[Susceptibi]	-22445.24 29241.412 -0.767584 .4855 ns	81187.176

col(10)[Acquired I]	-426.8303 547.87945 -0.779059 .4795 ns	1521.1572

Degrees of freedom for two-tailed t tests = 4

If P<=0.05, the coefficient is significantly different from 0.

Residuals:

Row	Y observed	Y expected	Residual

--------- ------------- ------------- -------------

1	2.988432 5.10432467459 -2.1158926746

2	10.886968 9.16833528827 1.71863271173

3	9.572431 6.93842005158 2.63401094842

4	9.538951 9.10018770885 0.43876329115

5	7.604563 10.3032117733 -2.6986487733

6	9.156931 11.4654725855 -2.3085415855

7	14.159849 11.6232216385 2.53662736145

8	11.288805 11.7430603955 -0.4542553955

9	11.875 11.6256957919 0.24930420807

Validation Method: Bootstrap

Validate N Times: 10

Leave-Group-Out PRESS	= 4892.36691306

Leave-Group-Out PRE R^2 = 0.5949411

Leave-Group-Out MAE	= 17.8567547238

(The validation method randomly assigns rows of data to validation groups,

so the Leave-Group-Out statistics printed above will vary.

You can reduce the variability by increasing 'Validate N Times'.)

Group Leave-Group-Out Validation Equations

----- ----------------------------------------------------------------------

1 1.38634964e-6*col(5)[pIVD] -4.5688625565*col(6)[pIR] -6.1202586e-5*col(7)[Vaccinated] +6851.01417228*col(9)[Susceptibi] +127.62544426*col(10)[Acquired I]

2 2.68913816e-5*col(5)[pIVD] -84.100536929*col(6)[pIR] +1.3860537e-4*col(7)[Vaccinated] -15636.974048*col(9)[Susceptibi] -287.21393956*col(10)[Acquired I]

3 -5.3625081e-4*col(5)[pIVD] +1666.22971061*col(6)[pIR] -4.7445728e-5*col(7)[Vaccinated] +16008.4971328*col(9)[Susceptibi] +0*col(10)[Acquired I]

4 -5.4331383e-6*col(5)[pIVD] +16.7832276805*col(6)[pIR] +6.01409598e-5*col(7)[Vaccinated] -6245.9857129*col(9)[Susceptibi] -129.60479209*col(10)[Acquired I]

5 2.59113814e-5*col(5)[pIVD] -82.158241091*col(6)[pIR] +2.09971292e-4*col(7)[Vaccinated] -23391.454065*col(9)[Susceptibi] -437.84532381*col(10)[Acquired I]

6 -5.0892136e-5*col(5)[pIVD] +160.398342332*col(6)[pIR] -5.3368809e-4*col(7)[Vaccinated] +60283.8471745*col(9)[Susceptibi] +1105.71604244*col(10)[Acquired I]

7 -3.0001603e-5*col(5)[pIVD] +94.7107579982*col(6)[pIR] -1.6446028e-4*col(7)[Vaccinated] +18983.8328354*col(9)[Susceptibi] +336.910848292*col(10)[Acquired I]

8 -1.0020738e-4*col(5)[pIVD] +314.020027497*col(6)[pIR] -6.5678393e-4*col(7)[Vaccinated] +74841.5037375*col(9)[Susceptibi] +1354.71804227*col(10)[Acquired I]

9 3.66471559e-5*col(5)[pIVD] -114.89131487*col(6)[pIR] +8.84174796e-4*col(7)[Vaccinated] -98437.980191*col(9)[Susceptibi] -1845.3072179*col(10)[Acquired I]

10 -5.2781588e-5*col(5)[pIVD] +167.25277311*col(6)[pIR] +8.36644212e-5*col(7)[Vaccinated] -7863.5687916*col(9)[Susceptibi] -188.59610503*col(10)[Acquired I]

(The validation method randomly assigns rows of data to validation groups,

so the LGO Validation Equations will vary. Since these equations are

generated during the individual validation runs, increasing

'Validate N Times' will not decrease the variability of the coefficients.)

REGRESSION: MULTIPLE (ONE SUBSET)

2021-03-18 23:05:08

Using: C:\Users\LENOVO\Desktop\Influenza-PLOS ONE\Data Influenza PLOS ONE\Medical Data\Model 1.dt

X Columns:

2) Population	6) pIR	9) Susceptibi

5) pIVD	7) Vaccinated	10) Acquired I

Y Column: 8) cIR

Keep If:

Calculate Constant: false

Total number of data points = 9

Number of data points used = 9

Regression equation:

col(8)[cIR] =

-4.2663828e-5*col(2)[Population]

+8.42022402e-6*col(5)[pIVD]

-26.748008888*col(6)[pIR]

+2.18949673e-5*col(7)[Vaccinated]

+20318.7974194*col(9)[Susceptibi]

+0*col(10)[Acquired I]

R^2	= 0.96319848933	AIC = 23.9313632588	MSEP	= 43.9232064691

adj R^2 =	0.917196601	BIC = 37.8063632588	PRESS	= 299.437530496

PRE R^2 = 0.00585884816	MAE = 1.68465799228	LOO MAE = 4.73402696505

For each term in the ANOVA table below, if P<=0.05, that term was a

significant source of Y's variation.

Source	SS	df	MS	F	P

------------------------ ------------- -------- --------- --------- ---------

Regression	886.829033853	5 177.36581 20.938238 .0057 **

col(2)[Population]	845.397109048	1 845.39711 99.800104 .0006 ***

col(5)[pIVD]	4.81829506386	1 4.8182951 0.5688053 .4927 ns

col(6)[pIR]	26.2371578128	1 26.237158 3.0973267 .1532 ns

col(7)[Vaccinated]	5.07230421513	1 5.0723042 0.5987914 .4822 ns

col(9)[Susceptibi]	5.30416771334	1 5.3041677 0.6261631 .4730 ns

col(10)[Acquired I]	0	0

Error	33.883616419	4 8.4709041

------------------------ ------------- -------- --------- --------- ---------

Total	920.712650272	9

Table of Statistics for the Regression Coefficients:

Column	Coef. Std Error t(Coef=0)	P	+/-95% CL

------------------------ --------- --------- --------- --------- ---------

col(2)[Population]	-4.266e-5 5.4808e-5 -0.778418 .4798 ns	1.5217e-4

col(5)[pIVD]	8.4202e-6 1.6886e-5 0.4986416 .6442 ns	4.6884e-5

col(6)[pIR]	-26.74801 52.969461	-0.50497 .6401 ns	147.0668

col(7)[Vaccinated]	2.1895e-5 2.9353e-5 0.7459104 .4972 ns	8.1498e-5

col(9)[Susceptibi]	20318.797	25677.59 0.7913047 .4730 ns	71292.419

col(10)[Acquired I]	0	0	0

Degrees of freedom for two-tailed t tests = 4

If P<=0.05, the coefficient is significantly different from 0.

Residuals:

Row	Y observed	Y expected	Residual

--------- ------------- ------------- -------------

1	2.988432 5.10629247384 -2.1178604738

2	10.886968 9.16733656898 1.71963143102

3	9.572431 6.93933666688 2.63309433312

4	9.538951 9.09616498054 0.44278601946

5	7.604563 10.3024488879 -2.6978858879

6	9.156931 11.4671325145 -2.3102015145

7	14.159849 11.6247834086 2.53506559138

8	11.288805 11.7438180851 -0.4550130851

9	11.875 11.6246164057 0.25038359428

Validation Method: Bootstrap

Validate N Times: 10

Leave-Group-Out PRESS	= 707.335968085

Leave-Group-Out PRE R^2 = 0.4586722

Leave-Group-Out MAE	= 10.5642833494

(The validation method randomly assigns rows of data to validation groups,

so the Leave-Group-Out statistics printed above will vary.

You can reduce the variability by increasing 'Validate N Times'.)

Group Leave-Group-Out Validation Equations

----- ----------------------------------------------------------------------

1 -3.0604259e-4*col(2)[Population] +1.6922817e-4*col(5)[pIVD] -538.04838398*col(6)[pIR] +1.68860062e-4*col(7)[Vaccinated] +141815.492671*col(9)[Susceptibi] +0*col(10)[Acquired I]

2 1.87076074e-6*col(2)[Population] -1.4975369e-5*col(5)[pIVD] +46.555136466*col(6)[pIR] -3.5794608e-6*col(7)[Vaccinated] +0*col(9)[Susceptibi] +0*col(10)[Acquired I]

3 -5.3959968e-6*col(2)[Population] +6.39292255e-5*col(5)[pIVD] -201.09785954*col(6)[pIR] +1.06256965e-5*col(7)[Vaccinated] +0*col(9)[Susceptibi] +0*col(10)[Acquired I]

4 5.89677228e-6*col(2)[Population] -4.5165791e-5*col(5)[pIVD] +142.964973741*col(6)[pIR] -8.4267786e-6*col(7)[Vaccinated] -1027.675374*col(9)[Susceptibi] +0*col(10)[Acquired I]

5 -1.1884246e-4*col(2)[Population] -3.7413452e-6*col(5)[pIVD] +10.958695357*col(6)[pIR] +5.95259927e-5*col(7)[Vaccinated] +57054.9570813*col(9)[Susceptibi] +0*col(10)[Acquired I]

6 5.47956374e-4*col(2)[Population] -8.6922841e-5*col(5)[pIVD] +273.473338555*col(6)[pIR] -2.8919668e-4*col(7)[Vaccinated] -257992.96632*col(9)[Susceptibi] +0*col(10)[Acquired I]

7 -7.1228919e-5*col(2)[Population] +1.40210967e-6*col(5)[pIVD] -4.9212838885*col(6)[pIR] +3.57622486e-5*col(7)[Vaccinated] +34175.3086219*col(9)[Susceptibi] +0*col(10)[Acquired I]

8 -4.7021941e-5*col(2)[Population] +1.70840575e-5*col(5)[pIVD] -54.399416229*col(6)[pIR] +2.44442556e-5*col(7)[Vaccinated] +22294.5539761*col(9)[Susceptibi] +0*col(10)[Acquired I]

9 1.45246117e-4*col(2)[Population] -1.047559e-4*col(5)[pIVD] +328.268866169*col(6)[pIR] -8.5538114e-5*col(7)[Vaccinated] -65429.235454*col(9)[Susceptibi] +0*col(10)[Acquired I]

10 6.45232553e-5*col(2)[Population] +4.85811284e-6*col(5)[pIVD] -15.038291267*col(6)[pIR] -3.2408878e-5*col(7)[Vaccinated] -30909.791543*col(9)[Susceptibi] +0*col(10)[Acquired I]

(The validation method randomly assigns rows of data to validation groups,

so the LGO Validation Equations will vary. Since these equations are

generated during the individual validation runs, increasing

'Validate N Times' will not decrease the variability of the coefficients.)

REGRESSION: MULTIPLE (ONE SUBSET)

2021-03-18 23:05:57

Using: C:\Users\LENOVO\Desktop\Influenza-PLOS ONE\Data Influenza PLOS ONE\Medical Data\Model 1.dt

X Columns:

5) pIVD	7) Vaccinated	10) Acquired I

6) pIR	9) Susceptibi

Y Column: 8) cIR

Keep If:

Calculate Constant: false

Total number of data points = 9

Number of data points used = 9

Regression equation:

col(8)[cIR] =

8.34201264e-6*col(5)[pIVD]

-26.502484246*col(6)[pIR]

+2.03597306e-4*col(7)[Vaccinated]

-22445.237562*col(9)[Susceptibi]

-426.83034907*col(10)[Acquired I]

R^2	= 0.96320646378	AIC = 21.9294128545	MSEP	= 21.9568444165

adj R^2 = 0.91721454351	BIC = 33.3044128545	PRESS	= 299.456592058

PRE R^2 = 0.0059770996	MAE = 1.68385299442	LOO MAE = 4.73143184106

For each term in the ANOVA table below, if P<=0.05, that term was a

significant source of Y's variation.

Source	SS	df	MS	F	P

------------------------ ------------- -------- --------- --------- ---------

Regression	886.83637603	5 177.36728 20.94295 .0057 **

col(5)[pIVD]	803.165229036	1 803.16523 94.835131 .0006 ***

col(6)[pIR]	0.77343968758	1 0.7734397 0.0913252 .7776 ns

col(7)[Vaccinated]	71.9838573998	1 71.983857 8.4996192 .0434 *

col(9)[Susceptibi]	5.77369488332	1 5.7736949 0.6817391 .4554 ns

col(10)[Acquired I]	5.14015502285	1 5.140155 0.6069327 .4795 ns

Error	33.8762742425	4 8.4690686

------------------------ ------------- -------- --------- --------- ---------

Total	920.712650272	9

Table of Statistics for the Regression Coefficients:

Column	Coef. Std Error t(Coef=0)	P	+/-95% CL

------------------------ --------- --------- --------- --------- ---------

col(5)[pIVD]	8.342e-6 1.6807e-5 0.4963287 .6457 ns	4.6665e-5

col(6)[pIR]	-26.50248 52.721736 -0.502686 .6416 ns	146.37901

col(7)[Vaccinated]	2.036e-4 2.6254e-4 0.7754903 .4813 ns	7.2893e-4

col(9)[Susceptibi]	-22445.24 29241.412 -0.767584 .4855 ns	81187.176

col(10)[Acquired I]	-426.8303 547.87945 -0.779059 .4795 ns	1521.1572

Degrees of freedom for two-tailed t tests = 4

If P<=0.05, the coefficient is significantly different from 0.

Residuals:

Row	Y observed	Y expected	Residual

--------- ------------- ------------- -------------

1	2.988432 5.10432467459 -2.1158926746

2	10.886968 9.16833528827 1.71863271173

3	9.572431 6.93842005158 2.63401094842

4	9.538951 9.10018770885 0.43876329115

5	7.604563 10.3032117733 -2.6986487733

6	9.156931 11.4654725855 -2.3085415855

7	14.159849 11.6232216385 2.53662736145

8	11.288805 11.7430603955 -0.4542553955

9	11.875 11.6256957919 0.24930420807

Validation Method: Bootstrap

Validate N Times: 10

Leave-Group-Out PRESS	= 734.852774873

Leave-Group-Out PRE R^2 = 0.385442

Leave-Group-Out MAE	= 9.82185506262

(The validation method randomly assigns rows of data to validation groups,

so the Leave-Group-Out statistics printed above will vary.

You can reduce the variability by increasing 'Validate N Times'.)

Group Leave-Group-Out Validation Equations

----- ----------------------------------------------------------------------

1 3.98339457e-6*col(5)[pIVD] -12.459264941*col(6)[pIR] -2.0573839e-4*col(7)[Vaccinated] +22639.3287021*col(9)[Susceptibi] +432.134234639*col(10)[Acquired I]

2 -1.1892109e-4*col(5)[pIVD] +376.656531363*col(6)[pIR] -1.4022377e-5*col(7)[Vaccinated] +4575.69621827*col(9)[Susceptibi] +0*col(10)[Acquired I]

3 1.66710463e-5*col(5)[pIVD] -50.909326174*col(6)[pIR] +2.25829917e-7*col(7)[Vaccinated] -81.463051203*col(9)[Susceptibi] +0*col(10)[Acquired I]

4 5.57137183e-5*col(5)[pIVD] -173.98810836*col(6)[pIR] +2.84595122e-4*col(7)[Vaccinated] -32270.98513*col(9)[Susceptibi] -588.38320307*col(10)[Acquired I]

5 2.29505686e-6*col(5)[pIVD] -6.827550643*col(6)[pIR] -0.0011477759*col(7)[Vaccinated] +126971.029078*col(9)[Susceptibi] +2403.55383762*col(10)[Acquired I]

6 -2.1851515e-5*col(5)[pIVD] +69.402906587*col(6)[pIR] -8.8191489e-5*col(7)[Vaccinated] +10307.8786933*col(9)[Susceptibi] +179.426899634*col(10)[Acquired I]

7 -5.1827198e-5*col(5)[pIVD] +164.031848668*col(6)[pIR] +3.30580402e-4*col(7)[Vaccinated] -35068.848034*col(9)[Susceptibi] -706.62087069*col(10)[Acquired I]

8 6.10885919e-5*col(5)[pIVD] -191.07643785*col(6)[pIR] +3.93772737e-6*col(7)[Vaccinated] -1293.4006058*col(9)[Susceptibi] +0*col(10)[Acquired I]

9 1.51533295e-6*col(5)[pIVD] -5.7304871064*col(6)[pIR] -1.4527895e-6*col(7)[Vaccinated] +516.615108469*col(9)[Susceptibi] +0*col(10)[Acquired I]

10 -2.1416267e-5*col(5)[pIVD] +67.9653965481*col(6)[pIR] -9.7799643e-5*col(7)[Vaccinated] +11372.4414514*col(9)[Susceptibi] +199.539310376*col(10)[Acquired I]

(The validation method randomly assigns rows of data to validation groups,

so the LGO Validation Equations will vary. Since these equations are

generated during the individual validation runs, increasing

'Validate N Times' will not decrease the variability of the coefficients.)

REGRESSION: MULTIPLE (ONE SUBSET)

2021-03-18 23:06:48

Using: C:\Users\LENOVO\Desktop\Influenza-PLOS ONE\Data Influenza PLOS ONE\Medical Data\Model 1.dt

X Columns:

5) pIVD	6) pIR	9) Susceptibi	10) Acquired I

Y Column: 8) cIR

Keep If:

Calculate Constant: false

Total number of data points = 9

Number of data points used = 9

Regression equation:

col(8)[cIR] =

-8.958175e-7*col(5)[pIVD]

+2.48442513657*col(6)[pIR]

+230.236038177*col(9)[Susceptibi]

-1.9596757773*col(10)[Acquired I]

R^2	= 0.95767469156	AIC = 21.1899801421	MSEP	= 15.1547849083

adj R^2 = 0.92381444481	BIC = 28.3099801421	PRESS	= 209.463866121

PRE R^2 = 0.00256956081	MAE = 1.89091400239	LOO MAE = 3.94289723872

For each term in the ANOVA table below, if P<=0.05, that term was a

significant source of Y's variation.

Source	SS	df	MS	F	P

------------------------ ------------- -------- --------- --------- ---------

Regression	881.743203365	4 220.4358 28.283157 .0013 **

col(5)[pIVD]	803.165229036	1 803.16523 103.05063 .0002 ***

col(6)[pIR]	0.77343968758	1 0.7734397 0.0992367 .7655 ns

col(9)[Susceptibi]	72.720481217	1 72.720481 9.3304482 .0283 *

col(10)[Acquired I]	5.08405342436	1 5.0840534 0.6523128 .4560 ns

Error	38.9694469071	5 7.7938894

------------------------ ------------- -------- --------- --------- ---------

Total	920.712650272	9

Table of Statistics for the Regression Coefficients:

Column	Coef. Std Error t(Coef=0)	P	+/-95% CL

------------------------ --------- --------- --------- --------- ---------

col(5)[pIVD]	-8.958e-7 1.1375e-5 -0.078756 .9403 ns	2.9239e-5

col(6)[pIR]	2.4844251 35.667859 0.0696545 .9472 ns	91.68715

col(9)[Susceptibi]	230.23604 257.87115 0.8928337 .4129 ns	662.87889

col(10)[Acquired I]	-1.959676 2.4263659 -0.807659 .4560 ns	6.2371721

Degrees of freedom for two-tailed t tests = 5

If P<=0.05, the coefficient is significantly different from 0.

Residuals:

Row	Y observed	Y expected	Residual

--------- ------------- ------------- -------------

1	2.988432	6.3081344891 -3.3197024891

2	10.886968 8.52665455749 2.36031344251

3	9.572431	7.1479624488	2.4244685512

4	9.538951 8.58205048313 0.95690051687

5	7.604563	9.6078341414 -2.0032711414

6	9.156931 11.2781738172 -2.1212428172

7	14.159849 11.7494570628 2.41039193723

8	11.288805 10.9317660961 0.35703890393

9	11.875	12.939896222	-1.064896222

Validation Method: Bootstrap

Validate N Times: 10

Leave-Group-Out PRESS	= 134.315744151

Leave-Group-Out PRE R^2 = 0.4759332

Leave-Group-Out MAE	= 4.63228727791

(The validation method randomly assigns rows of data to validation groups,

so the Leave-Group-Out statistics printed above will vary.

You can reduce the variability by increasing 'Validate N Times'.)

Group Leave-Group-Out Validation Equations

----- ----------------------------------------------------------------------

1 -1.3028701e-5*col(5)[pIVD] +41.7850018109*col(6)[pIR] +357.943858711*col(9)[Susceptibi] -3.4159960429*col(10)[Acquired I]

2 1.05931985e-6*col(5)[pIVD] -3.7255748492*col(6)[pIR] +165.894238566*col(9)[Susceptibi] -1.3180250981*col(10)[Acquired I]

3 -4.6444795e-5*col(5)[pIVD] +147.068578501*col(6)[pIR] +1211.51331984*col(9)[Susceptibi] -11.627626067*col(10)[Acquired I]

4 -1.974132e-5*col(5)[pIVD] +62.5476458105*col(6)[pIR] +573.892121041*col(9)[Susceptibi] -5.4013155079*col(10)[Acquired I]

5 3.84927925e-6*col(5)[pIVD] -12.240838735*col(6)[pIR] +21.020068595*col(9)[Susceptibi] +0.02727863619*col(10)[Acquired I]

6 -9.2366883e-6*col(5)[pIVD] +28.3466658714*col(6)[pIR] +514.645819549*col(9)[Susceptibi] -4.5882207108*col(10)[Acquired I]

7 -2.4910373e-5*col(5)[pIVD] +78.4703311305*col(6)[pIR] +727.504501449*col(9)[Susceptibi] -6.7947515253*col(10)[Acquired I]

8 5.10723301e-6*col(5)[pIVD] -16.791341333*col(6)[pIR] +221.679134575*col(9)[Susceptibi] -1.8055711074*col(10)[Acquired I]

9 -1.7706259e-5*col(5)[pIVD] +56.3028730002*col(6)[pIR] +483.94016076*col(9)[Susceptibi] -4.5749266892*col(10)[Acquired I]

10 -2.1928955e-5*col(5)[pIVD] +69.2207352073*col(6)[pIR] +677.95233932*col(9)[Susceptibi] -6.3424293333*col(10)[Acquired I]

(The validation method randomly assigns rows of data to validation groups,

so the LGO Validation Equations will vary. Since these equations are

generated during the individual validation runs, increasing

'Validate N Times' will not decrease the variability of the coefficients.)

REGRESSION: MULTIPLE (ONE SUBSET)

2021-03-18 23:07:37

Using: C:\Users\LENOVO\Desktop\Influenza-PLOS ONE\Data Influenza PLOS ONE\Medical Data\Model 1.dt

X Columns:

5) pIVD	6) pIR	7) Vaccinated	10) Acquired I

Y Column: 8) cIR

Keep If:

Calculate Constant: false

Total number of data points = 9

Number of data points used = 9

Regression equation:

col(8)[cIR] =

-8.7860444e-7*col(5)[pIVD]

+2.42941371991*col(6)[pIR]

+2.08426139e-6*col(7)[Vaccinated]

-6.3269916282*col(10)[Acquired I]

R^2	= 0.95778691329	AIC = 21.1660857641	MSEP	= 15.114603365

adj R^2 = 0.92401644393	BIC = 28.2860857641	PRESS	= 207.814459489

PRE R^2 = 0.00263833464	MAE = 1.88865272914	LOO MAE = 3.93267947631

For each term in the ANOVA table below, if P<=0.05, that term was a

significant source of Y's variation.

Source	SS	df	MS	F	P

------------------------ ------------- -------- --------- --------- ---------

Regression	881.846527334	4 220.46163 28.36167 .0012 **

col(5)[pIVD]	803.165229036	1 803.16523 103.32459 .0002 ***

col(6)[pIR]	0.77343968758	1 0.7734397 0.0995005 .7652 ns

col(7)[Vaccinated]	71.9838573998	1 71.983857 9.2604886 .0286 *

col(10)[Acquired I]	5.92400121025	1 5.9240012 0.7621034 .4226 ns

Error	38.8661229385	5 7.7732246

------------------------ ------------- -------- --------- --------- ---------

Total	920.712650272	9

Table of Statistics for the Regression Coefficients:

Column	Coef. Std Error t(Coef=0)	P	+/-95% CL

------------------------ --------- --------- --------- --------- ---------

col(5)[pIVD]	-8.786e-7 1.1262e-5 -0.078014 .9408 ns	2.895e-5

col(6)[pIR]	2.4294137 35.316179 0.0687904 .9478 ns	90.783128

col(7)[Vaccinated]	2.0843e-6 2.3122e-6	0.901423 .4087 ns	5.9437e-6

col(10)[Acquired I]	-6.326992 7.2475346 -0.872985 .4226 ns	18.630381

Degrees of freedom for two-tailed t tests = 5

If P<=0.05, the coefficient is significantly different from 0.

Residuals:

Row	Y observed	Y expected	Residual

--------- ------------- ------------- -------------

1	2.988432 6.28795098891 -3.2995189889

2	10.886968 8.52608328531 2.36088471469

3	9.572431 7.14702546084 2.42540553916

4	9.538951 8.58986518326 0.94908581674

5	7.604563 9.61894728715 -2.0143842872

6	9.156931 11.2901290687 -2.1331980687

7	14.159849 11.7561243505 2.40372464947

8	11.288805	10.929079254 0.35972574597

9	11.875 12.9269467515 -1.0519467515

Validation Method: Bootstrap

Validate N Times: 10

Leave-Group-Out PRESS	= 157.847322754

Leave-Group-Out PRE R^2 = 0.4920218

Leave-Group-Out MAE	= 5.36199392781

(The validation method randomly assigns rows of data to validation groups,

so the Leave-Group-Out statistics printed above will vary.

You can reduce the variability by increasing 'Validate N Times'.)

Group Leave-Group-Out Validation Equations

----- ----------------------------------------------------------------------

1 8.90905344e-6*col(5)[pIVD] -28.547736153*col(6)[pIR] +3.58005025e-7*col(7)[Vaccinated] -0.8616160438*col(10)[Acquired I]

2 6.1447371e-6*col(5)[pIVD] -19.372540486*col(6)[pIR] -4.1735975e-7*col(7)[Vaccinated] +1.52095093305*col(10)[Acquired I]

3 -2.339069e-6*col(5)[pIVD] +7.18182387377*col(6)[pIR] +1.28552427e-6*col(7)[Vaccinated] -3.8008003625*col(10)[Acquired I]

4 5.26279738e-5*col(5)[pIVD] -165.24789471*col(6)[pIR] -4.9166163e-6*col(7)[Vaccinated] +15.6378812788*col(10)[Acquired I]

5 2.39985661e-5*col(5)[pIVD] -74.292040575*col(6)[pIR] -7.8719301e-6*col(7)[Vaccinated] +24.7715999692*col(10)[Acquired I]

6 1.47617455e-5*col(5)[pIVD] -47.445925936*col(6)[pIR] +1.27610187e-6*col(7)[Vaccinated] -3.6899831242*col(10)[Acquired I]

7 -2.1501717e-5*col(5)[pIVD] +67.9326298414*col(6)[pIR] +5.96252569e-6*col(7)[Vaccinated] -18.666243521*col(10)[Acquired I]

8 -2.1032831e-5*col(5)[pIVD] +66.2456843078*col(6)[pIR] +5.9887649e-6*col(7)[Vaccinated] -18.712569036*col(10)[Acquired I]

9 -2.8533576e-5*col(5)[pIVD] +90.3960292522*col(6)[pIR] +6.69222539e-6*col(7)[Vaccinated] -21.027345959*col(10)[Acquired I]

10 -1.4032713e-5*col(5)[pIVD] +44.8368156516*col(6)[pIR] +3.65978603e-6*col(7)[Vaccinated] -11.503511276*col(10)[Acquired I]

(The validation method randomly assigns rows of data to validation groups,

so the LGO Validation Equations will vary. Since these equations are

generated during the individual validation runs, increasing

'Validate N Times' will not decrease the variability of the coefficients.)

REGRESSION: MULTIPLE (ONE SUBSET)

2021-03-18 23:08:44

Using: C:\Users\LENOVO\Desktop\Influenza-PLOS ONE\Data Influenza PLOS ONE\Medical Data\Model 1.dt

X Columns:

6) pIR	7) Vaccinated	9) Susceptibi	10) Acquired I

Y Column: 8) cIR

Keep If:

Calculate Constant: false

Total number of data points = 9

Number of data points used = 9

Regression equation:

col(8)[cIR] =

-0.3360922124*col(6)[pIR]

+1.11242984e-4*col(7)[Vaccinated]

-12072.331631*col(9)[Susceptibi]

-234.92717312*col(10)[Acquired I]

R^2	= 0.96094051355	AIC = 20.4672852028	MSEP	= 13.9854412794

adj R^2 = 0.92969292439	BIC = 27.5872852028	PRESS	= 153.615433985

PRE R^2 = 0.00179208061	MAE = 1.74291397173	LOO MAE = 3.40061424513

For each term in the ANOVA table below, if P<=0.05, that term was a

significant source of Y's variation.

Source	SS	df	MS	F	P

------------------------ ------------- -------- --------- --------- ---------

Regression	884.750086983	4 221.18752 30.752469 .0010 **

col(6)[pIR]	802.689197508	1 802.6892 111.60067 .0001 ***

col(7)[Vaccinated]	44.5326817417	1 44.532682 6.1915333 .0553 ns

col(9)[Susceptibi]	34.4261180093	1 34.426118 4.7863827 .0803 ns

col(10)[Acquired I]	3.1020897233	1 3.1020897 0.4312943 .5404 ns

Error	35.9625632898	5 7.1925127

------------------------ ------------- -------- --------- --------- ---------

Total	920.712650272	9

Table of Statistics for the Regression Coefficients:

Column	Coef. Std Error t(Coef=0)	P	+/-95% CL

------------------------ --------- --------- --------- --------- ---------

col(6)[pIR]	-0.336092 0.4071269 -0.825522 .4467 ns	1.0465529

col(7)[Vaccinated]	1.1124e-4 1.7068e-4 0.6517504 .5433 ns	4.3876e-4

col(9)[Susceptibi]	-12072.33 18847.619 -0.640523 .5500 ns	48449.348

col(10)[Acquired I]	-234.9272 357.72261	-0.65673 .5404 ns	919.55525

Degrees of freedom for two-tailed t tests = 5

If P<=0.05, the coefficient is significantly different from 0.

Residuals:

Row	Y observed	Y expected	Residual

--------- ------------- ------------- -------------

1	2.988432 5.41878109963 -2.4303490996

2	10.886968 8.44282296994 2.44414503006

3	9.572431 7.32076936578 2.25166163422

4	9.538951	9.090634103	0.448316897

5	7.604563 10.2181468259 -2.6135838259

6	9.156931 11.7772172487 -2.6202862487

7	14.159849	11.96177956 2.19806944004

8	11.288805 10.7878848334 0.50092016662

9	11.875 12.0538934035 -0.1788934035

Validation Method: Bootstrap

Validate N Times: 10

Leave-Group-Out PRESS	= 39.4769729337

Leave-Group-Out PRE R^2 = 0.6181832

Leave-Group-Out MAE	= 3.22775313068

(The validation method randomly assigns rows of data to validation groups,

so the Leave-Group-Out statistics printed above will vary.

You can reduce the variability by increasing 'Validate N Times'.)

Group Leave-Group-Out Validation Equations

----- ----------------------------------------------------------------------

1 -0.4854967615*col(6)[pIR] +2.68218653e-4*col(7)[Vaccinated] -29371.765674*col(9)[Susceptibi] -564.21548524*col(10)[Acquired I]

2 0.39392782696*col(6)[pIR] +1.80477757e-4*col(7)[Vaccinated] -19773.307877*col(9)[Susceptibi] -379.63979802*col(10)[Acquired I]

3 -0.2024727103*col(6)[pIR] +2.68353301e-5*col(7)[Vaccinated] -2676.8585597*col(9)[Susceptibi] -58.769446418*col(10)[Acquired I]

4 -0.6546391441*col(6)[pIR] +3.43919617e-4*col(7)[Vaccinated] -37686.278554*col(9)[Susceptibi] -723.26125376*col(10)[Acquired I]

5 0.20202748444*col(6)[pIR] +2.27042661e-4*col(7)[Vaccinated] -24882.115424*col(9)[Susceptibi] -477.5075386*col(10)[Acquired I]

6 -0.237462034*col(6)[pIR] +8.28089425e-5*col(7)[Vaccinated] -9001.0100703*col(9)[Susceptibi] -174.69354906*col(10)[Acquired I]

7 0.0432999835*col(6)[pIR] -6.6666229e-7*col(7)[Vaccinated] +236.762216626*col(9)[Susceptibi] +0*col(10)[Acquired I]

8 0.3073043094*col(6)[pIR] +1.99439946e-4*col(7)[Vaccinated] -21881.170443*col(9)[Susceptibi] -419.23874651*col(10)[Acquired I]

9 -0.8049159124*col(6)[pIR] +2.70308539e-4*col(7)[Vaccinated] -29553.082083*col(9)[Susceptibi] -569.01433824*col(10)[Acquired I]

10 -0.7705386265*col(6)[pIR] -1.7907037e-5*col(7)[Vaccinated] +2311.94182741*col(9)[Susceptibi] +34.6204617683*col(10)[Acquired I]

(The validation method randomly assigns rows of data to validation groups,

so the LGO Validation Equations will vary. Since these equations are

generated during the individual validation runs, increasing

'Validate N Times' will not decrease the variability of the coefficients.)

REGRESSION: MULTIPLE (ONE SUBSET)

2021-03-18 23:09:55

Using: C:\Users\LENOVO\Desktop\Influenza-PLOS ONE\Data Influenza PLOS ONE\Medical Data\Model 1.dt

X Columns:

5) pIVD	9) Susceptibi	5) pIVD

6) pIR	10) Acquired I

Y Column: 8) cIR

Keep If:

Calculate Constant: false

Total number of data points = 9

Number of data points used = 9

Regression equation:

col(8)[cIR] =

-8.958175e-7*col(5)[pIVD]

+2.48442513657*col(6)[pIR]

+230.236038177*col(9)[Susceptibi]

-1.9596757773*col(10)[Acquired I]

+0*col(5)[pIVD]

R^2	= 0.95767469156	AIC = 23.1899801421	MSEP	= 25.2579748472

adj R^2 = 0.92381444481	BIC = 31.9099801421	PRESS	= 209.463866121

PRE R^2 = 0.00256956081	MAE = 1.89091400239	LOO MAE = 3.94289723872

For each term in the ANOVA table below, if P<=0.05, that term was a

significant source of Y's variation.

Source	SS	df	MS	F	P

------------------------ ------------- -------- --------- --------- ---------

Regression	881.743203365	4 220.4358 28.283157 .0013 **

col(5)[pIVD]	803.165229036	1 803.16523 103.05063 .0002 ***

col(6)[pIR]	0.77343968758	1 0.7734397 0.0992367 .7655 ns

col(9)[Susceptibi]	72.720481217	1 72.720481 9.3304482 .0283 *

col(10)[Acquired I]	5.08405342436	1 5.0840534 0.6523128 .4560 ns

col(5)[pIVD]	0	0

Error	38.9694469071	5 7.7938894

------------------------ ------------- -------- --------- --------- ---------

Total	920.712650272	9

Table of Statistics for the Regression Coefficients:

Column	Coef. Std Error t(Coef=0)	P	+/-95% CL

------------------------ --------- --------- --------- --------- ---------

col(5)[pIVD]	-8.958e-7 1.1375e-5 -0.078756 .9403 ns	2.9239e-5

col(6)[pIR]	2.4844251 35.667859 0.0696545 .9472 ns	91.68715

col(9)[Susceptibi]	230.23604 257.87115 0.8928337 .4129 ns	662.87889

col(10)[Acquired I]	-1.959676 2.4263659 -0.807659 .4560 ns	6.2371721

col(5)[pIVD]	0	0	0

Degrees of freedom for two-tailed t tests = 5

If P<=0.05, the coefficient is significantly different from 0.

Residuals:

Row	Y observed	Y expected	Residual

--------- ------------- ------------- -------------

1	2.988432	6.3081344891 -3.3197024891

2	10.886968 8.52665455749 2.36031344251

3	9.572431	7.1479624488	2.4244685512

4	9.538951 8.58205048313 0.95690051687

5	7.604563	9.6078341414 -2.0032711414

6	9.156931 11.2781738172 -2.1212428172

7	14.159849 11.7494570628 2.41039193723

8	11.288805 10.9317660961 0.35703890393

9	11.875	12.939896222	-1.064896222

Validation Method: Bootstrap

Validate N Times: 10

Leave-Group-Out PRESS	= 1127.06139041

Leave-Group-Out PRE R^2 = 0.3882605

Leave-Group-Out MAE	= 9.56865307417

(The validation method randomly assigns rows of data to validation groups,

so the Leave-Group-Out statistics printed above will vary.

You can reduce the variability by increasing 'Validate N Times'.)

Group Leave-Group-Out Validation Equations

----- ----------------------------------------------------------------------

1 1.6055046e-5*col(5)[pIVD] -50.856678216*col(6)[pIR] -142.06017221*col(9)[Susceptibi] +1.58323185267*col(10)[Acquired I] +0*col(5)[pIVD]

2 6.64880597e-8*col(5)[pIVD] -0.2526731702*col(6)[pIR] +238.499837129*col(9)[Susceptibi] -2.1308522608*col(10)[Acquired I] +0*col(5)[pIVD]

3 1.77398104e-5*col(5)[pIVD] -55.506662581*col(6)[pIR] -342.11362642*col(9)[Susceptibi] +3.40980347651*col(10)[Acquired I] +0*col(5)[pIVD]

4 1.47489801e-5*col(5)[pIVD] -46.844411492*col(6)[pIR] -100.37447649*col(9)[Susceptibi] +1.19841957003*col(10)[Acquired I] +0*col(5)[pIVD]

5 -2.1851944e-5*col(5)[pIVD] +69.1553933548*col(6)[pIR] +618.057337476*col(9)[Susceptibi] -5.8117315844*col(10)[Acquired I] +0*col(5)[pIVD]

6 -2.5375638e-4*col(5)[pIVD] +789.879162396*col(6)[pIR] +5096.55976711*col(9)[Susceptibi] -47.211873178*col(10)[Acquired I] +0*col(5)[pIVD]

7 2.17529464e-6*col(5)[pIVD] -7.1042229052*col(6)[pIR] +189.083451328*col(9)[Susceptibi] -1.6011410441*col(10)[Acquired I] +0*col(5)[pIVD]

8 -2.7073393e-5*col(5)[pIVD] +84.2790712879*col(6)[pIR] +370.323010788*col(9)[Susceptibi] -3.2356971291*col(10)[Acquired I] +0*col(5)[pIVD]

9 -6.8368928e-5*col(5)[pIVD] +216.533540537*col(6)[pIR] +1772.98079686*col(9)[Susceptibi] -17.117674323*col(10)[Acquired I] +0*col(5)[pIVD]

10 1.00898452e-5*col(5)[pIVD] -31.736041677*col(6)[pIR] -138.92021716*col(9)[Susceptibi] +1.52099657195*col(10)[Acquired I] +0*col(5)[pIVD]

(The validation method randomly assigns rows of data to validation groups,

so the LGO Validation Equations will vary. Since these equations are

generated during the individual validation runs, increasing

'Validate N Times' will not decrease the variability of the coefficients.)

REGRESSION: MULTIPLE (ONE SUBSET)

2021-03-18 10:29:30

Using: C:\Users\LENOVO\Desktop\Influenza-PLOS ONE\Data Influenza PLOS ONE\Medical Data\Model 1.dt

X Columns:

7) Vaccinated	3) cYear	9) Susceptibi	10) Acq Immuni

Y Column: 8) Infection

Keep If:

Calculate Constant: false

Total number of data points = 9

Number of data points used = 9

Regression equation:

col(8)[Infection ] =

7.2080364e-5*col(7)[Vaccinated]

-8.3405725107*col(3)[cYear]

+10654.3439337*col(9)[Susceptibi]

+0*col(10)[Acq Immuni]

R^2	= 0.9555848445	AIC = 21.623739966	MSEP	= 15.9030649311

adj R^2 = 0.93337726674	BIC = 27.123739966	PRESS	= 125.963562856

PRE R^2 = 0.01219970762	MAE = 1.71379034024	LOO MAE = 2.77996355107

For each term in the ANOVA table below, if P<=0.05, that term was a

significant source of Y's variation.

Source	SS	df	MS	F	P

------------------------ ------------- -------- --------- --------- ---------

Regression	879.819054735	3 293.27302 43.029675 .0002 ***

col(7)[Vaccinated]	840.970595424	1 840.9706 123.38909 .0000 ***

col(3)[cYear]	35.9924912145	1 35.992491 5.280899 .0613 ns

col(9)[Susceptibi]	2.855968097	1 2.8559681 0.419034 .5414 ns

col(10)[Acq Immuni]	0	0

Error	40.893595537	6 6.8155993

------------------------ ------------- -------- --------- --------- ---------

Total	920.712650272	9

Table of Statistics for the Regression Coefficients:

Column	Coef. Std Error t(Coef=0)	P	+/-95% CL

------------------------ --------- --------- --------- --------- ---------

col(7)[Vaccinated]	7.208e-5 1.1481e-4 0.6278061 .5533 ns	2.8094e-4

col(3)[cYear]	-8.340573 13.168477 -0.633374 .5499 ns	32.222103

col(9)[Susceptibi]	10654.344 16458.947 0.6473284 .5414 ns	40273.594

col(10)[Acq Immuni]	0	0	0

Degrees of freedom for two-tailed t tests = 6

If P<=0.05, the coefficient is significantly different from 0.

Residuals:

Row	Y observed	Y expected	Residual

--------- ------------- ------------- -------------

1	2.988432	5.7351445635 -2.7467125635

2	10.886968 7.17332055375 3.71364744625

3	9.572431 8.41993459428 1.15249640572

4	9.538951 9.46397193934 0.07497906066

5	7.604563	10.316303174	-2.711740174

6	9.156931 10.9767841374 -1.8198531374

7	14.159849 11.4455589904	2.7142900096

8	11.288805 11.7225556526 -0.4337506526

9	11.875 11.8183563876 0.05664361244

Validation Method: Bootstrap

Validate N Times: 10

Leave-Group-Out PRESS	= 62.929226226

Leave-Group-Out PRE R^2 = 0.7747452

Leave-Group-Out MAE	= 3.55535913009

(The validation method randomly assigns rows of data to validation groups,

so the Leave-Group-Out statistics printed above will vary.

You can reduce the variability by increasing 'Validate N Times'.)

Group Leave-Group-Out Validation Equations

----- ----------------------------------------------------------------------

1 5.16495711e-5*col(7)[Vaccinated] -5.9555338501*col(3)[cYear] +7557.22002145*col(9)[Susceptibi] +0*col(10)[Acq Immuni]

2 -2.6573488e-4*col(7)[Vaccinated] +30.4290269807*col(3)[cYear] -37866.802513*col(9)[Susceptibi] +0*col(10)[Acq Immuni]

3 -7.8147002e-5*col(7)[Vaccinated] +8.94280500007*col(3)[cYear] -11097.131112*col(9)[Susceptibi] +0*col(10)[Acq Immuni]

4 -5.9511574e-6*col(7)[Vaccinated] +0.63259442566*col(3)[cYear] -623.4911354*col(9)[Susceptibi] +0*col(10)[Acq Immuni]

5 -1.3636801e-4*col(7)[Vaccinated] +15.6452593154*col(3)[cYear] -19545.652484*col(9)[Susceptibi] +0*col(10)[Acq Immuni]

6 -2.3081883e-4*col(7)[Vaccinated] +26.4868018883*col(3)[cYear] -33121.069305*col(9)[Susceptibi] +0*col(10)[Acq Immuni]

7 1.42133847e-4*col(7)[Vaccinated] -16.399353454*col(3)[cYear] +20795.150927*col(9)[Susceptibi] +0*col(10)[Acq Immuni]

8 1.1644749e-4*col(7)[Vaccinated] -13.477936702*col(3)[cYear] +17213.8885519*col(9)[Susceptibi] +0*col(10)[Acq Immuni]

9 3.88161773e-5*col(7)[Vaccinated] -4.4968995836*col(3)[cYear] +5768.84948835*col(9)[Susceptibi] +0*col(10)[Acq Immuni]

10 9.99420591e-5*col(7)[Vaccinated] -11.538764626*col(3)[cYear] +14661.0528705*col(9)[Susceptibi] +0*col(10)[Acq Immuni]

(The validation method randomly assigns rows of data to validation groups,

so the LGO Validation Equations will vary. Since these equations are

generated during the individual validation runs, increasing

'Validate N Times' will not decrease the variability of the coefficients.)

REGRESSION: MULTIPLE (ONE SUBSET)

2021-03-18 10:30:49

Using: C:\Users\LENOVO\Desktop\Influenza-PLOS ONE\Data Influenza PLOS ONE\Medical Data\Model 1.dt

X Columns:

7) Vaccinated	9) Susceptibi	10) Acq Immuni

Y Column: 8) Infection

Keep If:

Calculate Constant: false

Total number of data points = 9

Number of data points used = 9

Regression equation:

col(8)[Infection ] =

1.05059482e-4*col(7)[Vaccinated]

-11443.386817*col(9)[Susceptibi]

-221.51065225*col(10)[Acq Immuni]

R^2	= 0.95561680962	AIC = 19.6172604287	MSEP	= 10.5944131082

adj R^2 = 0.93342521442	BIC = 24.1172604287	PRESS	= 126.165393979

PRE R^2 = 0.01282920887	MAE = 1.71382745947	LOO MAE = 2.78278224784

For each term in the ANOVA table below, if P<=0.05, that term was a

significant source of Y's variation.

Source	SS	df	MS	F	P

------------------------ ------------- -------- --------- --------- ---------

Regression	879.848485426	3 293.28283 43.062105 .0002 ***

col(7)[Vaccinated]	840.970595424	1 840.9706 123.47796 .0000 ***

col(9)[Susceptibi]	36.1142947676	1 36.114295 5.3025865 .0609 ns

col(10)[Acq Immuni]	2.76359523499	1 2.7635952 0.4057729 .5476 ns

Error	40.864164846	6 6.8106941

------------------------ ------------- -------- --------- --------- ---------

Total	920.712650272	9

Table of Statistics for the Regression Coefficients:

Column	Coef. Std Error t(Coef=0)	P	+/-95% CL

------------------------ --------- --------- --------- --------- ---------

col(7)[Vaccinated]	1.0506e-4 1.6593e-4 0.6331508 .5500 ns	4.0602e-4

col(9)[Susceptibi]	-11443.39 18325.539	-0.62445 .5553 ns	44840.979

col(10)[Acq Immuni]	-221.5107 347.73875 -0.637003 .5476 ns	850.88607

Degrees of freedom for two-tailed t tests = 6

If P<=0.05, the coefficient is significantly different from 0.

Residuals:

Row	Y observed	Y expected	Residual

--------- ------------- ------------- -------------

1	2.988432 5.72806719734 -2.7396351973

2	10.886968 7.17671989199 3.71024810801

3	9.572431 8.41855136134 1.15387963866

4	9.538951 9.46868839721 0.07026260279

5	7.604563 10.3202114936 -2.7156484936

6	9.156931 10.9773407446 -1.8204097446

7	14.159849 11.4444949716 2.71535402838

8	11.288805 11.7253347962 -0.4365297962

9	11.875 11.8125204744 0.06247952559

Validation Method: Bootstrap

Validate N Times: 10

Leave-Group-Out PRESS	= 80.3519556862

Leave-Group-Out PRE R^2 = 0.4584562

Leave-Group-Out MAE	= 4.29861739824

(The validation method randomly assigns rows of data to validation groups,

so the Leave-Group-Out statistics printed above will vary.

You can reduce the variability by increasing 'Validate N Times'.)

Group Leave-Group-Out Validation Equations

----- ----------------------------------------------------------------------

1 1.72134556e-6*col(7)[Vaccinated] +75.383407782*col(9)[Susceptibi] -5.9892290433*col(10)[Acq Immuni]

2 6.57823143e-5*col(7)[Vaccinated] -7057.5676538*col(9)[Susceptibi] -139.68794255*col(10)[Acq Immuni]

3 1.02674674e-4*col(7)[Vaccinated] -11243.258109*col(9)[Susceptibi] -215.92069731*col(10)[Acq Immuni]

4 -4.4293391e-4*col(7)[Vaccinated] +49068.831811*col(9)[Susceptibi] +927.022699452*col(10)[Acq Immuni]

5 -5.6665057e-5*col(7)[Vaccinated] +6412.20582216*col(9)[Susceptibi] +117.470459818*col(10)[Acq Immuni]

6 -6.4107369e-4*col(7)[Vaccinated] +71004.456925*col(9)[Susceptibi] +1341.77406405*col(10)[Acq Immuni]

7 2.13780066e-4*col(7)[Vaccinated] -23410.432583*col(9)[Susceptibi] -449.74842773*col(10)[Acq Immuni]

8 4.02655335e-4*col(7)[Vaccinated] -44102.720995*col(9)[Susceptibi] -847.18080159*col(10)[Acq Immuni]

9 1.56076041e-4*col(7)[Vaccinated] -17075.254736*col(9)[Susceptibi] -328.44408142*col(10)[Acq Immuni]

10 4.74303907e-4*col(7)[Vaccinated] -52211.837549*col(9)[Susceptibi] -995.40356879*col(10)[Acq Immuni]

(The validation method randomly assigns rows of data to validation groups,

so the LGO Validation Equations will vary. Since these equations are

generated during the individual validation runs, increasing

'Validate N Times' will not decrease the variability of the coefficients.)

REGRESSION: MULTIPLE (ONE SUBSET)

2021-03-18 10:31:37

Using: C:\Users\LENOVO\Desktop\Influenza-PLOS ONE\Data Influenza PLOS ONE\Medical Data\Model 1.dt

X Columns:

7) Vaccinated	10) Acq Immuni

Y Column: 8) Infection

Keep If:

Calculate Constant: false

Total number of data points = 9

Number of data points used = 9

Regression equation:

col(8)[Infection ] =

1.44438397e-6*col(7)[Vaccinated]

-4.3683375543*col(10)[Acq Immuni]

R^2	= 0.95273236034	AIC = 18.1839463966	MSEP	= 8.05924329269

adj R^2 = 0.93922732044	BIC = 21.1635382333	PRESS	= 83.3889577568

PRE R^2 = 0.08983397638	MAE = 1.88245703619	LOO MAE = 2.51479938813

For each term in the ANOVA table below, if P<=0.05, that term was a

significant source of Y's variation.

Source	SS	df	MS	F	P

------------------------ ------------- -------- --------- --------- ---------

Regression	877.192736492	2 438.59637 70.546431 .0000 ***

col(7)[Vaccinated]	840.970595424	1 840.9706 135.26668 .0000 ***

col(10)[Acq Immuni]	36.222141068	1 36.222141 5.8261831 .0465 *

Error	43.5199137805	7 6.2171305

------------------------ ------------- -------- --------- --------- ---------

Total	920.712650272	9

Table of Statistics for the Regression Coefficients:

Column	Coef. Std Error t(Coef=0)	P	+/-95% CL

------------------------ --------- --------- --------- --------- ---------

col(7)[Vaccinated]	1.4444e-6 5.7355e-7 2.5183179 .0399 *	1.3562e-6

col(10)[Acq Immuni]	-4.368338 1.8097731 -2.413749 .0465 *	4.2794333

Degrees of freedom for two-tailed t tests = 7

If P<=0.05, the coefficient is significantly different from 0.

Residuals:

Row	Y observed	Y expected	Residual

--------- ------------- ------------- -------------

1	2.988432 6.59163094663 -3.6031989466

2	10.886968 7.38110987108 3.50585812892

3	9.572431 8.16244617915 1.40998482085

4	9.538951 8.93571564733 0.60323535267

5	7.604563 9.70100560717 -2.0964426072

6	9.156931 10.4584005367 -1.3014695367

7	14.159849	11.207986323 2.95186267698

8	11.288805 11.9498357835 -0.6610307835

9	11.875 12.6840304723 -0.8090304723

Validation Method: Bootstrap

Validate N Times: 10

Leave-Group-Out PRESS	= 34.911365911

Leave-Group-Out PRE R^2 = 0.6763069

Leave-Group-Out MAE	= 2.68131152655

(The validation method randomly assigns rows of data to validation groups,

so the Leave-Group-Out statistics printed above will vary.

You can reduce the variability by increasing 'Validate N Times'.)

Group Leave-Group-Out Validation Equations

----- ----------------------------------------------------------------------

1 8.87967288e-7*col(7)[Vaccinated] -2.6085031806*col(10)[Acq Immuni]

2 1.63677453e-6*col(7)[Vaccinated] -4.9941043331*col(10)[Acq Immuni]

3 5.0254337e-7*col(7)[Vaccinated] -1.3834407408*col(10)[Acq Immuni]

4 6.20620442e-7*col(7)[Vaccinated] -1.7402649249*col(10)[Acq Immuni]

5 6.44051374e-7*col(7)[Vaccinated] -1.8357556762*col(10)[Acq Immuni]

6 2.4154063e-6*col(7)[Vaccinated] -7.4415598547*col(10)[Acq Immuni]

7 1.22547764e-6*col(7)[Vaccinated] -3.6815134188*col(10)[Acq Immuni]

8 1.96509402e-6*col(7)[Vaccinated] -6.0390374172*col(10)[Acq Immuni]

9 1.16688979e-6*col(7)[Vaccinated] -3.4724853909*col(10)[Acq Immuni]

10 2.26461597e-6*col(7)[Vaccinated] -6.964746043*col(10)[Acq Immuni]

(The validation method randomly assigns rows of data to validation groups,

so the LGO Validation Equations will vary. Since these equations are

generated during the individual validation runs, increasing

'Validate N Times' will not decrease the variability of the coefficients.)
