## Supplementary File (S5) for "Influenza outbreak, disease transmission rate and mortality risk in the United States (2021 to 2050 Projection)"

Analysis

REGRESSION: MULTIPLE (ONE SUBSET)

2021-03-18 10:36:59

Using: C:\Users\LENOVO\Desktop\Influenza-PLOS ONE\Data Influenza PLOS ONE\Medical Data\Model 1.dt

X Columns:

2) Population	6) P Infectio	8) Infection	10) Acq Immuni

5) P IVD Outb	7) Vaccinated	9) Susceptibi

Y Column: 1) IVD Outbre

Keep If:

Calculate Constant: false

Total number of data points = 9

Number of data points used = 9

Regression equation:

col(1)[IVD Outbre] =

3.52063180815*col(2)[Population]

-0.4805729234*col(5)[P IVD Outb]

+1511798.16419*col(6)[P Infectio]

-1.9531558115*col(7)[Vaccinated]

+3143546.64542*col(8)[Infection ]

-1626218899.4*col(9)[Susceptibi]

+0*col(10)[Acq Immuni]

R^2	= 0.99999747905	AIC = 209.052843681	MSEP	= 90434009808.2

adj R^2 = 0.99999243716	BIC = 231.052843681	PRESS	= 747002587243

PRE R^2 = 0.99919533603	MAE = 41093.4527254	LOO MAE = 222333.258701

For each term in the ANOVA table below, if P<=0.05, that term was a

significant source of Y's variation.

Source	SS	df	MS	F	P

------------------------ ------------- -------- --------- --------- ---------

Regression	9.22446675e15	6 1.5374e15 198337.59 .0000 ***

col(2)[Population]	8.45240879e15	1 8.4524e15 1090424.2 .0000 ***

col(5)[P IVD Outb]	5.60507161e13	1 5.6051e13 7230.9635 .0000 ***

col(6)[P Infectio]	2.78847279e14	1 2.7885e14 35973.394 .0000 ***

col(7)[Vaccinated]	5.2545784e13	1 5.2546e13 6778.8009 .0000 ***

col(8)[Infection ]	3.84584798e14	1 3.8458e14 49614.328 .0000 ***

col(9)[Susceptibi]	29377734705.9	1 2.9378e10 3.7899485 .1467 ns

col(10)[Acq Immuni]	0	0

Error	23254459665	3 7.75149e9

------------------------ ------------- -------- --------- --------- ---------

Total	9.22449e15	9

Table of Statistics for the Regression Coefficients:

Column	Coef. Std Error t(Coef=0)	P	+/-95% CL

------------------------ --------- --------- --------- --------- ---------

col(2)[Population]	3.5206318 1.7791128 1.9788693 .1422 ns	5.661931

col(5)[P IVD Outb]	-0.480573 0.5264509 -0.912854 .4286 ns	1.6754019

col(6)[P Infectio]	1511798.2 1652618.9 0.9147894 .4278 ns	5259370.8

col(7)[Vaccinated]	-1.953156 0.9476879 -2.060969 .1314 ns	3.0159658

col(8)[Infection ]	3143546.6 15125.078 207.83672 .0000 *** 48134.748

col(9)[Susceptibi]	-1.6262e9 8.35338e8 -1.946779 .1467 ns	2.65842e9

col(10)[Acq Immuni]	0	0	0

Degrees of freedom for two-tailed t tests = 3

If P<=0.05, the coefficient is significantly different from 0.

Residuals:

Row	Y observed	Y expected	Residual

--------- ------------- ------------- -------------

1	9300000 9244562.34402 55437.6559753

2	34000000 34033647.3783	-33647.37831

3	30000000 30055379.1431 -55379.143111

4	30000000 29999211.4619 788.538111567

5	24000000 24011128.7283 -11128.728261

6	29000000 29022748.3309 -22748.330897

7	45000000 44895571.9603 104428.039688

8	36000000 35975733.6967 24266.3033329

9	38000000 38062016.9568 -62016.956842

Validation Method: Bootstrap

Validate N Times: 10

Leave-Group-Out PRESS	= 463820784649

Leave-Group-Out PRE R^2 = 0.9962866

Leave-Group-Out MAE	= 263528.945613

(The validation method randomly assigns rows of data to validation groups,

so the Leave-Group-Out statistics printed above will vary.

You can reduce the variability by increasing 'Validate N Times'.)

Group Leave-Group-Out Validation Equations

----- ----------------------------------------------------------------------

1 1.39640662957*col(2)[Population] +1.09303552781*col(5)[P IVD Outb] -3467392.3497*col(6)[P Infectio] -0.6894362989*col(7)[Vaccinated] +3176318.40615*col(8)[Infection ] -672916092.27*col(9)[Susceptibi] +0*col(10)[Acq Immuni]

2 2.2298991911*col(2)[Population] +0.38650559007*col(5)[P IVD Outb] -1223002.4682*col(6)[P Infectio] -1.1983091089*col(7)[Vaccinated] +3147753.93717*col(8)[Infection ] -1042623793.3*col(9)[Susceptibi] +0*col(10)[Acq Immuni]

3 16.6873937889*col(2)[Population] -5.399228287*col(5)[P IVD Outb] +16872442.261*col(6)[P Infectio] -9.1612859935*col(7)[Vaccinated] +3188866.94527*col(8)[Infection ] -7740047789.5*col(9)[Susceptibi] +0*col(10)[Acq Immuni]

4 0.77493172887*col(2)[Population] -0.2009647577*col(5)[P IVD Outb] +635090.960009*col(6)[P Infectio] -0.5205179682*col(7)[Vaccinated] +3173334.58458*col(8)[Infection ] -328619744.24*col(9)[Susceptibi] +0*col(10)[Acq Immuni]

5 -0.0270245132*col(2)[Population] +1.32223130796*col(5)[P IVD Outb] -4179528.5833*col(6)[P Infectio] +0.05443110334*col(7)[Vaccinated] +3151493.41741*col(8)[Infection ] +0*col(9)[Susceptibi] +0*col(10)[Acq Immuni]

6 2.00134673367*col(2)[Population] +0.51392023845*col(5)[P IVD Outb] -1628423.7169*col(6)[P Infectio] -1.0701037392*col(7)[Vaccinated] +3149258.20345*col(8)[Infection ] -937434790.61*col(9)[Susceptibi] +0*col(10)[Acq Immuni]

7 1.3607256797*col(2)[Population] +1.10680595119*col(5)[P IVD Outb] -3510835.0632*col(6)[P Infectio] -0.6694795794*col(7)[Vaccinated] +3176510.89032*col(8)[Infection ] -656485912.53*col(9)[Susceptibi] +0*col(10)[Acq Immuni]

8 -0.0286850124*col(2)[Population] +1.5881757383*col(5)[P IVD Outb] -5062018.491*col(6)[P Infectio] +0.0597730942*col(7)[Vaccinated] +3165113.20178*col(8)[Infection ] +0*col(9)[Susceptibi] +0*col(10)[Acq Immuni]

9 -0.0033612591*col(2)[Population] +0.05328702242*col(5)[P IVD Outb] -159092.607*col(6)[P Infectio] -0.0978901486*col(7)[Vaccinated] +3172574.88459*col(8)[Infection ] +33879940.7966*col(9)[Susceptibi] +0*col(10)[Acq Immuni]

10 -0.0730516317*col(2)[Population] +1.84648349069*col(5)[P IVD Outb] -5847995.1322*col(6)[P Infectio] +0.14392468059*col(7)[Vaccinated] +3185169.80734*col(8)[Infection ] +0*col(9)[Susceptibi] +0*col(10)[Acq Immuni]

(The validation method randomly assigns rows of data to validation groups,

so the LGO Validation Equations will vary. Since these equations are

generated during the individual validation runs, increasing

'Validate N Times' will not decrease the variability of the coefficients.)

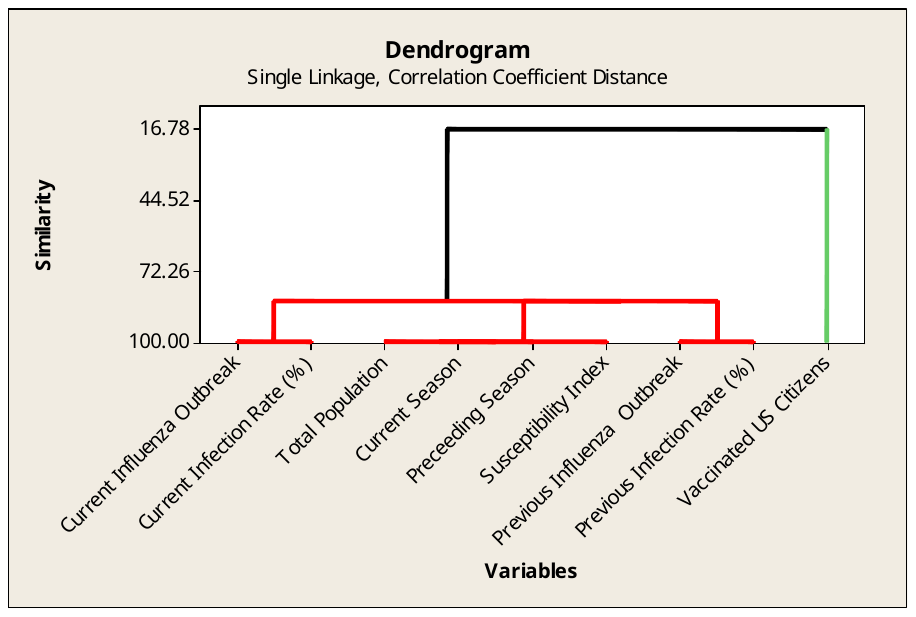

**—————	3/18/2021 11:10:45 AM	————————————————————**

Welcome to Minitab, press F1 for help.

Retrieving worksheet from file:

'C:\Users\LENOVO\Desktop\Influenza-PLOS

ONE\Data Influenza PLOS ONE\Medical Data\Model 1.xls'

Worksheet was saved on Thu Mar 18 2021

**Results for: Model 1**

**Cluster Analysis of Variables: Current Infl, Total Popula, Current Seas, ...**

```
Correlation Coefficient Distance, Single Linkage
Amalgamation Steps


                                                           Number
                                                          of obs.
      Number of  Similarity  Distance  Clusters      New   in new
Step   clusters       level     level   joined   cluster  cluster
   1          8     100.000   0.00000  3      4        3        2
   2          7     100.000   0.00000  2      3        2        3
   3          6      99.998   0.00003  2      9        2        4
   4          5      99.987   0.00027  5      6        5        2
   5          4      99.984   0.00033  1      8        1        2
   6          3      84.318   0.31364  2      5        2        6
   7          2      84.186   0.31628  1      2        1        8
   8          1      16.781   1.66439  1      7        1        9


Final Partition

Cluster 1
  Current Influenza Outbreak  Total Population  Current Season  Preceeding
     Season  Previous Influenza  Outbreak  Previous Infection Rate (%)  Current
     Infection Rate (%)  Susceptibility Index
Cluster 2
  Vaccinated US Citizens
```

**Dendrogram**

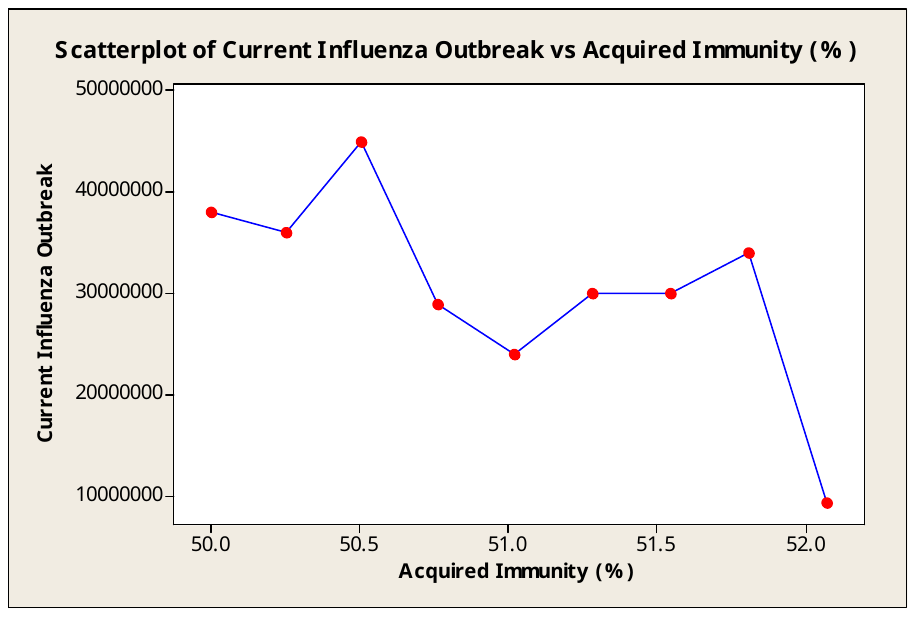

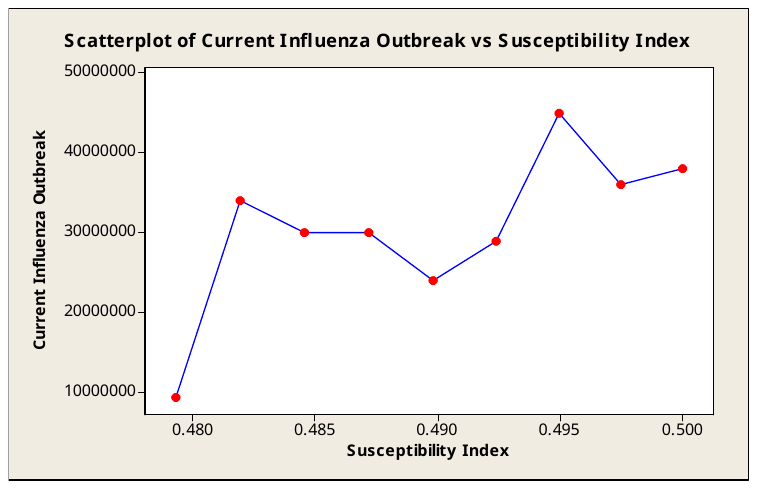

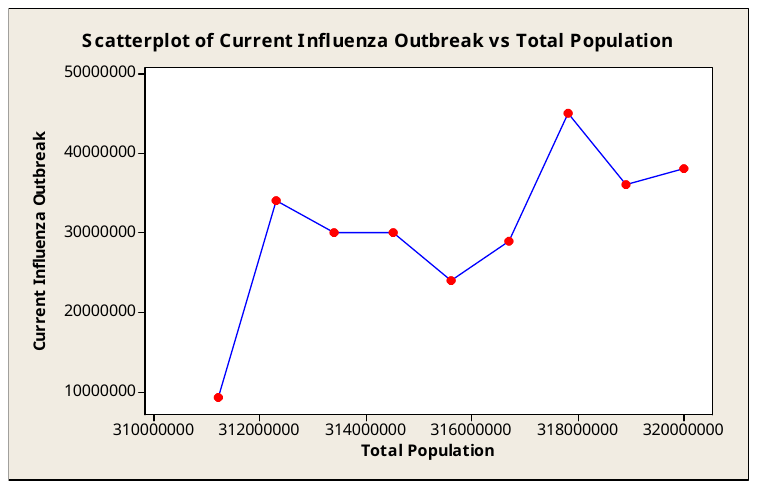

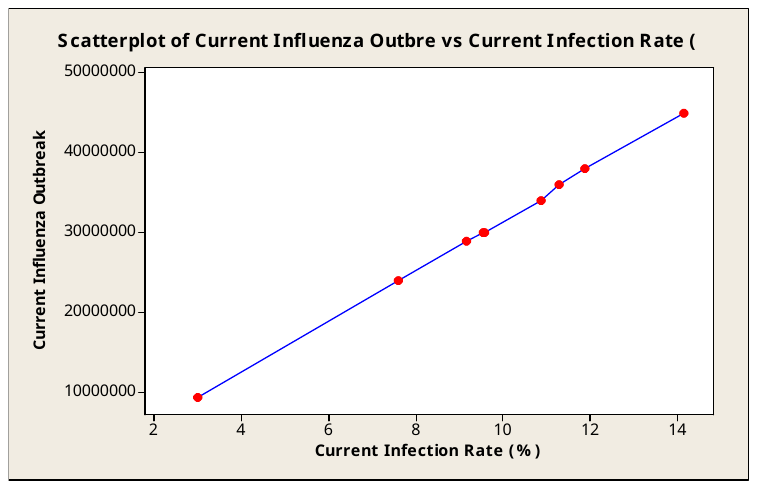

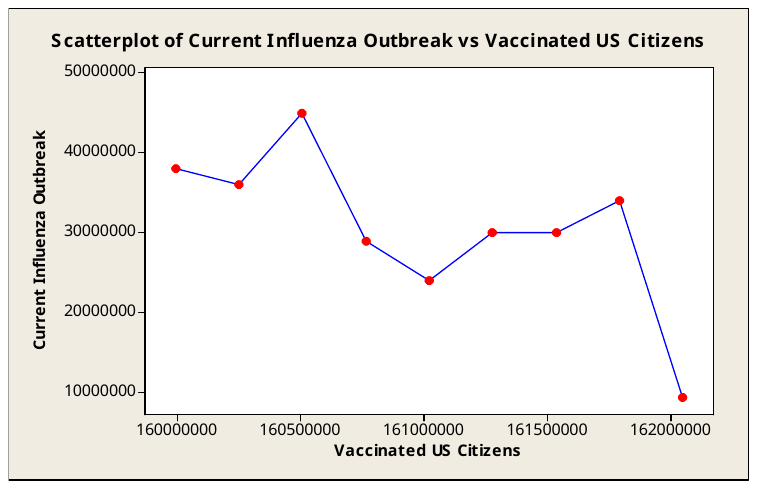

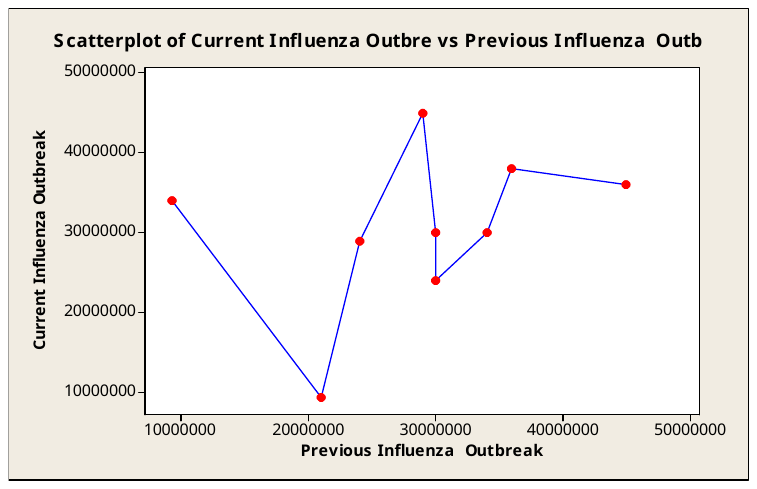

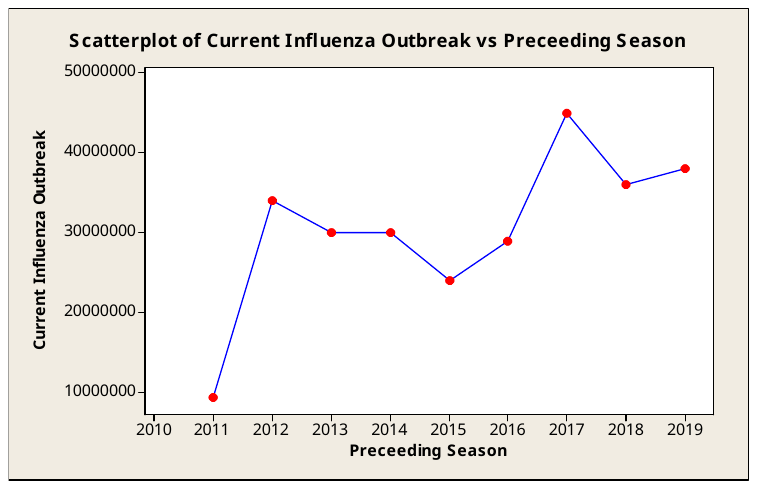

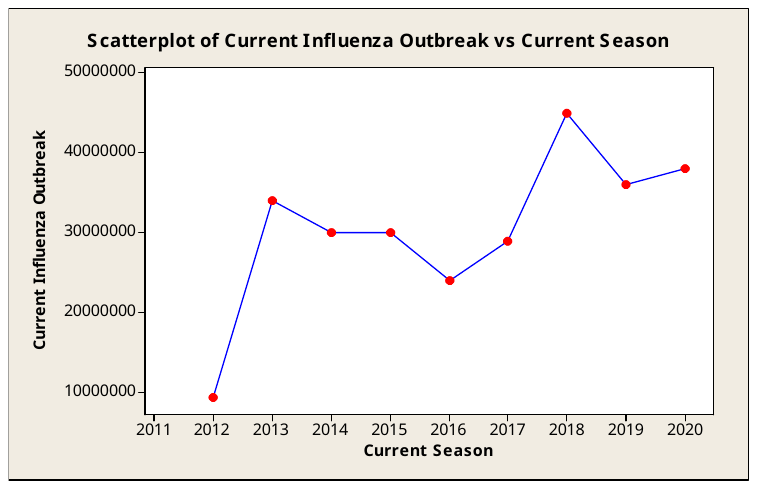

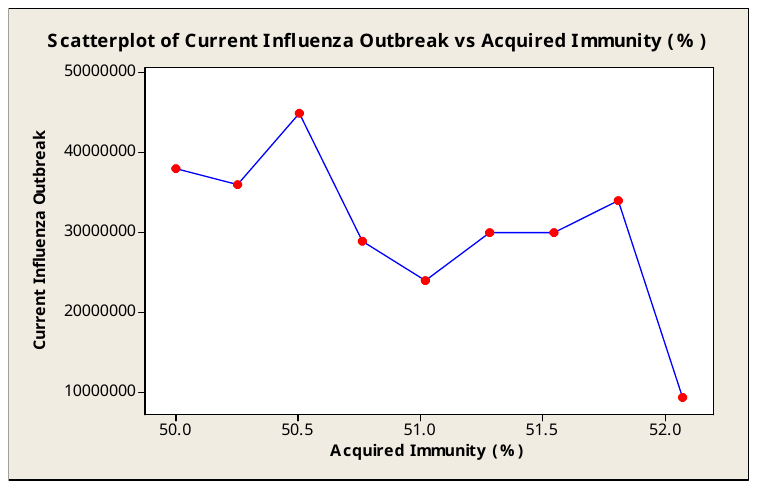

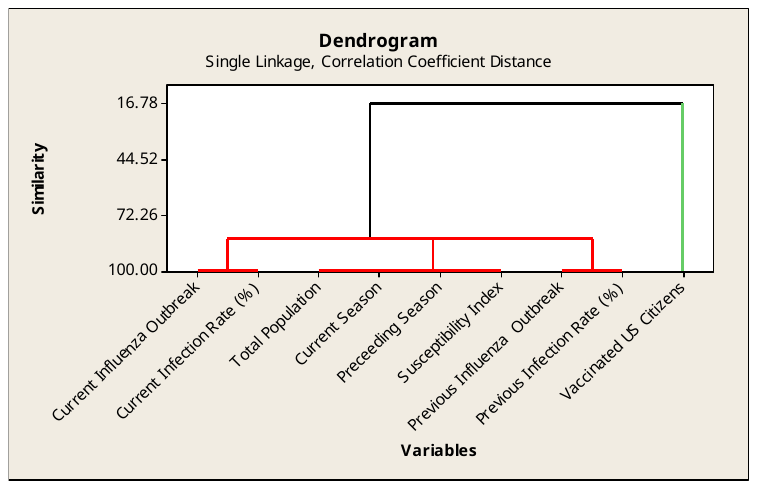

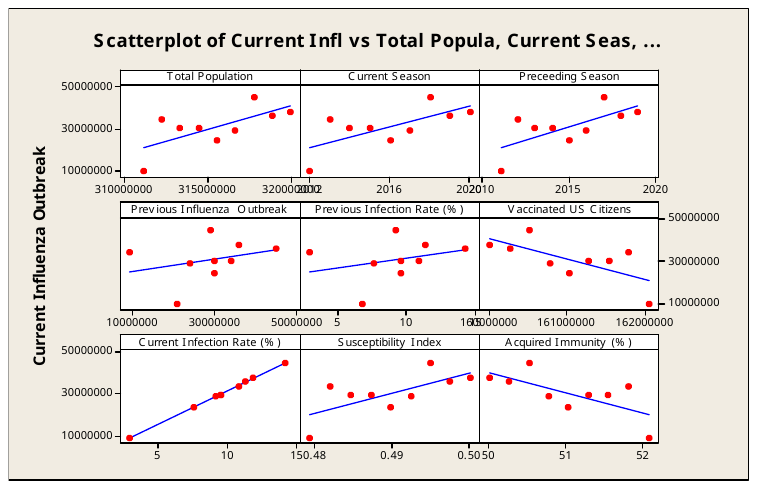

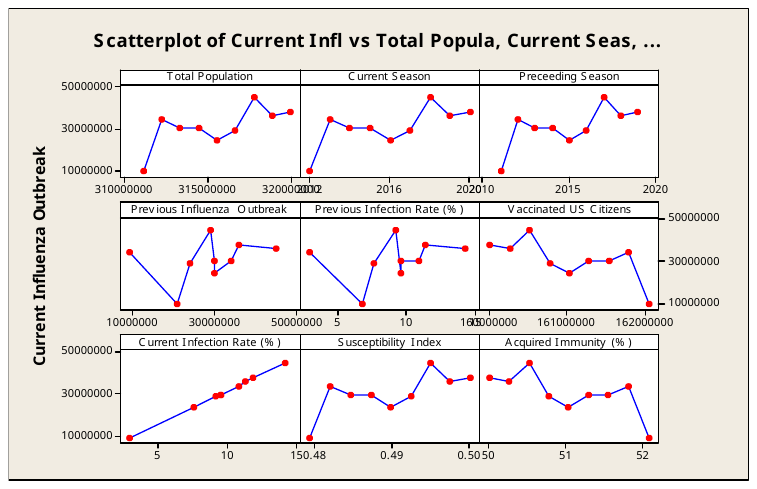

Fitted with group and connect line or trend line

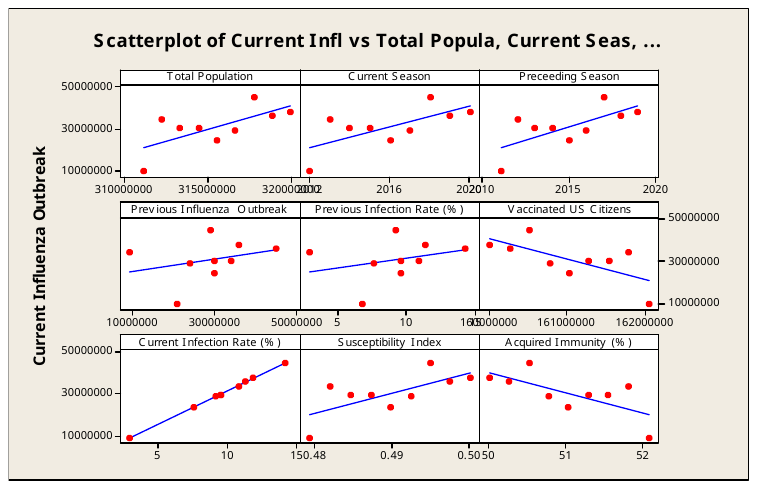

Fitted regression line

REGRESSION: MULTIPLE (ONE SUBSET)

2021-03-18 07:31:57

Using: C:\Users\LENOVO\Desktop\Influenza-PLOS ONE\Data Influenza PLOS ONE\Medical Data\Model 1.dt

X Columns:

2) Population	4) pYear	6) P Infectio

3) cYear	5) P IVD Outb	7) Vaccinated

Y Column: 1) IVD Outbre

Keep If:

Calculate Constant: false

Total number of data points = 9

Number of data points used = 9

Regression equation:

col(1)[IVD Outbre] =

3.74160251697*col(2)[Population]

-565818.53403*col(3)[cYear]

+0*col(4)[pYear]

-2.6075454402*col(5)[P IVD Outb]

+7166215.4418*col(6)[P Infectio]

+0*col(7)[Vaccinated]

R^2	= 0.95830257261	AIC = 294.474880884	MSEP	= 4.98604169e14

adj R^2 = 0.9249446307	BIC = 304.794880884	PRESS	= 2.07079299e15

PRE R^2 = 0.00817221526	MAE = 5937369.19209	LOO MAE =	12379954.23

For each term in the ANOVA table below, if P<=0.05, that term was a

significant source of Y's variation.

Source	SS	df	MS	F	P

------------------------ ------------- -------- --------- --------- ---------

Regression	8.8398525e15	4	2.21e15 28.727869 .0012 **

col(2)[Population]	8.45240879e15	1 8.4524e15	109.875 .0001 ***

col(3)[cYear]	3.42712986e14	1 3.4271e14 4.4550126 .0885 ns

col(4)[pYear]	0	0

col(5)[P IVD Outb]	4.44276605e13	1 4.4428e13 0.5775264 .4816 ns

col(6)[P Infectio]	303059772530	1 3.0306e11 0.0039396 .9524 ns

col(7)[Vaccinated]	0	0

Error	3.84637502e14	5 7.6928e13

------------------------ ------------- -------- --------- --------- ---------

Total	9.22449e15	9

Table of Statistics for the Regression Coefficients:

Column	Coef. Std Error t(Coef=0)	P	+/-95% CL

------------------------ --------- --------- --------- --------- ---------

col(2)[Population]	3.7416025	4.39801	0.850749 .4337 ns	11.305445

col(3)[cYear]	-565818.5 684619.62 -0.826471 .4462 ns	1759870.8

col(4)[pYear]	0	0	0

col(5)[P IVD Outb]	-2.607545 36.411931 -0.071612 .9457 ns	93.599848

col(6)[P Infectio]	7166215.4 1.14174e8 0.0627658 .9524 ns	2.93493e8

col(7)[Vaccinated]	0	0	0

Degrees of freedom for two-tailed t tests = 5

If P<=0.05, the coefficient is significantly different from 0.

Residuals:

Row	Y observed	Y expected	Residual

--------- ------------- ------------- -------------

1	9300000	19716637.105 -10416637.105

2	34000000 26675331.9943 7324668.00572

3	30000000 22421514.5066 7578485.49338

4	30000000 26981385.1539 3018614.84613

5	24000000 30291404.4955 -6291404.4955

6	29000000 35624380.2154 -6624380.2154

7	45000000 37261200.7819 7738799.21808

8	36000000 34942406.1989	1057593.8011

9	38000000 41385739.5485 -3385739.5485

Validation Method: Bootstrap

Validate N Times: 10

Leave-Group-Out PRESS	= 6.93144071e14

Leave-Group-Out PRE R^2 = 0.5409275

Leave-Group-Out MAE	= 11423718.524

(The validation method randomly assigns rows of data to validation groups,

so the Leave-Group-Out statistics printed above will vary.

You can reduce the variability by increasing 'Validate N Times'.)

Group Leave-Group-Out Validation Equations

----- ----------------------------------------------------------------------

1 0.35934577159*col(2)[Population] -35469.88961*col(3)[cYear] +0*col(4)[pYear] +33.6100207855*col(5)[P IVD Outb] -107465484.22*col(6)[P Infectio] +0*col(7)[Vaccinated]

2 5.07592390256*col(2)[Population] -768959.27657*col(3)[cYear] +0*col(4)[pYear] -18.954023005*col(5)[P IVD Outb] +57842147.328*col(6)[P Infectio] +0*col(7)[Vaccinated]

3 9.65421103359*col(2)[Population] -1501658.7318*col(3)[cYear] +0*col(4)[pYear] -56.138806706*col(5)[P IVD Outb] +177674670.027*col(6)[P Infectio] +0*col(7)[Vaccinated]

4 3.08488079877*col(2)[Population] -462315.44214*col(3)[cYear] +0*col(4)[pYear] +2.82045822587*col(5)[P IVD Outb] -9919774.8326*col(6)[P Infectio] +0*col(7)[Vaccinated]

5 10.4853152479*col(2)[Population] -1631769.592*col(3)[cYear] +0*col(4)[pYear] -69.266220305*col(5)[P IVD Outb] +219153726.411*col(6)[P Infectio] +0*col(7)[Vaccinated]

6 -1.1216248962*col(2)[Population] +191539.044033*col(3)[cYear] +0*col(4)[pYear] +22.1281120841*col(5)[P IVD Outb] -69764566.016*col(6)[P Infectio] +0*col(7)[Vaccinated]

7 4.79587713013*col(2)[Population] -718709.30218*col(3)[cYear] +0*col(4)[pYear] +36.2427884464*col(5)[P IVD Outb] -118167060.44*col(6)[P Infectio] +0*col(7)[Vaccinated]

8 3.67700368748*col(2)[Population] -556911.78464*col(3)[cYear] +0*col(4)[pYear] -8.8428182954*col(5)[P IVD Outb] +27095777.7157*col(6)[P Infectio] +0*col(7)[Vaccinated]

9 8.05847952269*col(2)[Population] -1256930.332*col(3)[cYear] +0*col(4)[pYear] -53.343078583*col(5)[P IVD Outb] +169970055.674*col(6)[P Infectio] +0*col(7)[Vaccinated]

10 4.36129763759*col(2)[Population] -657021.19692*col(3)[cYear] +0*col(4)[pYear] +11.1764231938*col(5)[P IVD Outb] -37671331.433*col(6)[P Infectio] +0*col(7)[Vaccinated]

(The validation method randomly assigns rows of data to validation groups,

so the LGO Validation Equations will vary. Since these equations are

generated during the individual validation runs, increasing

'Validate N Times' will not decrease the variability of the coefficients.)

REGRESSION: MULTIPLE (ONE SUBSET)

2021-03-18 07:33:07

Using: C:\Users\LENOVO\Desktop\Influenza-PLOS ONE\Data Influenza PLOS ONE\Medical Data\Model 1.dt

X Columns:

2) Population	4) pYear	6) P Infectio

3) cYear	5) P IVD Outb

Y Column: 1) IVD Outbre

Keep If:

Calculate Constant: false

Total number of data points = 9

Number of data points used = 9

Regression equation:

col(1)[IVD Outbre] =

3.74160251697*col(2)[Population]

-565818.53403*col(3)[cYear]

+0*col(4)[pYear]

-2.6075454402*col(5)[P IVD Outb]

+7166215.4418*col(6)[P Infectio]

R^2	= 0.95830257261	AIC = 292.474880884	MSEP	= 2.49302085e14

adj R^2 = 0.9249446307	BIC = 301.194880884	PRESS	= 2.07079299e15

PRE R^2 = 0.00817221526	MAE = 5937369.19209	LOO MAE =	12379954.23

For each term in the ANOVA table below, if P<=0.05, that term was a

significant source of Y's variation.

Source	SS	df	MS	F	P

------------------------ ------------- -------- --------- --------- ---------

Regression	8.8398525e15	4	2.21e15 28.727869 .0012 **

col(2)[Population]	8.45240879e15	1 8.4524e15	109.875 .0001 ***

col(3)[cYear]	3.42712986e14	1 3.4271e14 4.4550126 .0885 ns

col(4)[pYear]	0	0

col(5)[P IVD Outb]	4.44276605e13	1 4.4428e13 0.5775264 .4816 ns

col(6)[P Infectio]	303059772530	1 3.0306e11 0.0039396 .9524 ns

Error	3.84637502e14	5 7.6928e13

------------------------ ------------- -------- --------- --------- ---------

Total	9.22449e15	9

Table of Statistics for the Regression Coefficients:

Column	Coef. Std Error t(Coef=0)	P	+/-95% CL

------------------------ --------- --------- --------- --------- ---------

col(2)[Population]	3.7416025	4.39801	0.850749 .4337 ns	11.305445

col(3)[cYear]	-565818.5 684619.62 -0.826471 .4462 ns	1759870.8

col(4)[pYear]	0	0	0

col(5)[P IVD Outb]	-2.607545 36.411931 -0.071612 .9457 ns	93.599848

col(6)[P Infectio]	7166215.4 1.14174e8 0.0627658 .9524 ns	2.93493e8

Degrees of freedom for two-tailed t tests = 5

If P<=0.05, the coefficient is significantly different from 0.

Residuals:

Row	Y observed	Y expected	Residual

--------- ------------- ------------- -------------

1	9300000	19716637.105 -10416637.105

2	34000000 26675331.9943 7324668.00572

3	30000000 22421514.5066 7578485.49338

4	30000000 26981385.1539 3018614.84613

5	24000000 30291404.4955 -6291404.4955

6	29000000 35624380.2154 -6624380.2154

7	45000000 37261200.7819 7738799.21808

8	36000000 34942406.1989	1057593.8011

9	38000000 41385739.5485 -3385739.5485

Validation Method: Bootstrap

Validate N Times: 10

Leave-Group-Out PRESS	= 8.56759301e14

Leave-Group-Out PRE R^2 = 0.5754614

Leave-Group-Out MAE	= 14515001.3725

(The validation method randomly assigns rows of data to validation groups,

so the Leave-Group-Out statistics printed above will vary.

You can reduce the variability by increasing 'Validate N Times'.)

Group Leave-Group-Out Validation Equations

----- ----------------------------------------------------------------------

1 7.81681945596*col(2)[Population] -1196212.3038*col(3)[cYear] +0*col(4)[pYear] -25.302643529*col(5)[P IVD Outb] +77281631.3733*col(6)[P Infectio]

2 3.18815903212*col(2)[Population] -474190.74068*col(3)[cYear] +0*col(4)[pYear] +26.483273658*col(5)[P IVD Outb] -85767307.968*col(6)[P Infectio]

3 7.26635890774*col(2)[Population] -1133514.0086*col(3)[cYear] +0*col(4)[pYear] -46.794760204*col(5)[P IVD Outb] +149420461.29*col(6)[P Infectio]

4 9.08190721855*col(2)[Population] -1411471.7487*col(3)[cYear] +0*col(4)[pYear] -52.679202166*col(5)[P IVD Outb] +166610320.037*col(6)[P Infectio]

5 16.0965982844*col(2)[Population] -2508118.3101*col(3)[cYear] +0*col(4)[pYear] -107.68903903*col(5)[P IVD Outb] +339783738.789*col(6)[P Infectio]

6 3.68393455452*col(2)[Population] -549791.88125*col(3)[cYear] +0*col(4)[pYear] +21.2294968337*col(5)[P IVD Outb] -69671815.816*col(6)[P Infectio]

7 -2.5882433107*col(2)[Population] +422623.33871*col(3)[cYear] +0*col(4)[pYear] +51.2292972871*col(5)[P IVD Outb] -162241434.26*col(6)[P Infectio]

8 14.4646960133*col(2)[Population] -2227581.9389*col(3)[cYear] +0*col(4)[pYear] -83.269849668*col(5)[P IVD Outb] +257375224.276*col(6)[P Infectio]

9 1.11958050792*col(2)[Population] -149637.50533*col(3)[cYear] +0*col(4)[pYear] +56.5307710435*col(5)[P IVD Outb] -180920767.79*col(6)[P Infectio]

10 2.95749047384*col(2)[Population] -439857.97035*col(3)[cYear] +0*col(4)[pYear] +18.8117767535*col(5)[P IVD Outb] -61273891.394*col(6)[P Infectio]

(The validation method randomly assigns rows of data to validation groups,

so the LGO Validation Equations will vary. Since these equations are

generated during the individual validation runs, increasing

'Validate N Times' will not decrease the variability of the coefficients.)

REGRESSION: MULTIPLE (ONE SUBSET)

2021-03-18 07:34:08

Using: C:\Users\LENOVO\Desktop\Influenza-PLOS ONE\Data Influenza PLOS ONE\Medical Data\Model 1.dt

X Columns:

7) Vaccinated	4) pYear	6) P Infectio

3) cYear	5) P IVD Outb

Y Column: 1) IVD Outbre

Keep If:

Calculate Constant: false

Total number of data points = 9

Number of data points used = 9

Regression equation:

col(1)[IVD Outbre] =

-10.476657258*col(7)[Vaccinated]

+856710.348693*col(3)[cYear]

+0*col(4)[pYear]

-2.6075946455*col(5)[P IVD Outb]

+7166371.10062*col(6)[P Infectio]

R^2	= 0.95830259227	AIC = 292.474876641	MSEP	= 2.49301967e14

adj R^2 = 0.92494466609	BIC = 301.194876641	PRESS	= 2.07079055e15

PRE R^2 = 0.00817224034	MAE = 5937368.63218	LOO MAE = 12379952.1326

For each term in the ANOVA table below, if P<=0.05, that term was a

significant source of Y's variation.

Source	SS	df	MS	F	P

------------------------ ------------- -------- --------- --------- ---------

Regression	8.83985268e15	4	2.21e15 28.727883 .0012 **

col(7)[Vaccinated]	8.40635364e15	1 8.4064e15 109.27636 .0001 ***

col(3)[cYear]	3.88768377e14	1 3.8877e14 5.0537007 .0745 ns

col(4)[pYear]	0	0

col(5)[P IVD Outb]	4.44275886e13	1 4.4428e13 0.5775257 .4816 ns

col(6)[P Infectio]	303072886636	1 3.0307e11 0.0039397 .9524 ns

Error	3.84637321e14	5 7.6927e13

------------------------ ------------- -------- --------- --------- ---------

Total	9.22449e15	9

Table of Statistics for the Regression Coefficients:

Column	Coef. Std Error t(Coef=0)	P	+/-95% CL

------------------------ --------- --------- --------- --------- ---------

col(7)[Vaccinated]	-10.47666 12.314605 -0.850751 .4337 ns	31.655701

col(3)[cYear]	856710.35 987505.43	0.86755 .4253 ns	2538463.5

col(4)[pYear]	0	0	0

col(5)[P IVD Outb]	-2.607595 36.411925 -0.071614 .9457 ns	93.599832

col(6)[P Infectio]	7166371.1 1.14174e8 0.0627672 .9524 ns	2.93493e8

Degrees of freedom for two-tailed t tests = 5

If P<=0.05, the coefficient is significantly different from 0.

Residuals:

Row	Y observed	Y expected	Residual

--------- ------------- ------------- -------------

1	9300000	19716627.279 -10416627.279

2	34000000 26675330.5846 7324669.41537

3	30000000 22421517.1213 7578482.87871

4	30000000 26981390.8425 3018609.15749

5	24000000 30291405.3681 -6291405.3681

6	29000000 35624386.0872 -6624386.0872

7	45000000 37261202.6624 7738797.33764

8	36000000	34942399.938 1057600.06196

9	38000000 41385740.1041 -3385740.1041

Validation Method: Bootstrap

Validate N Times: 10

Leave-Group-Out PRESS	= 3.28356205e15

Leave-Group-Out PRE R^2 = 0.420705

Leave-Group-Out MAE	= 19302771.3431

(The validation method randomly assigns rows of data to validation groups,

so the Leave-Group-Out statistics printed above will vary.

You can reduce the variability by increasing 'Validate N Times'.)

Group Leave-Group-Out Validation Equations

----- ----------------------------------------------------------------------

1 -17.719552945*col(7)[Vaccinated] +1447921.1847*col(3)[cYear] +0*col(4)[pYear] +18.6731287597*col(5)[P IVD Outb] -62718027.123*col(6)[P Infectio]

2 -6.4173637538*col(7)[Vaccinated] +533676.944721*col(3)[cYear] +0*col(4)[pYear] -1.9697126505*col(5)[P IVD Outb] +5225437.17832*col(6)[P Infectio]

3 -23.237298371*col(7)[Vaccinated] +1864526.06861*col(3)[cYear] +0*col(4)[pYear] -46.708959298*col(5)[P IVD Outb] +148091531.961*col(6)[P Infectio]

4 13.907305601*col(7)[Vaccinated] -1099800.2632*col(3)[cYear] +0*col(4)[pYear] +49.5510173374*col(5)[P IVD Outb] -155004522.07*col(6)[P Infectio]

5 -23.765501842*col(7)[Vaccinated] +1901885.12391*col(3)[cYear] +0*col(4)[pYear] -62.398087141*col(5)[P IVD Outb] +198604002.677*col(6)[P Infectio]

6 -25.5940178*col(7)[Vaccinated] +2051440.85048*col(3)[cYear] +0*col(4)[pYear] -57.283686758*col(5)[P IVD Outb] +181825073.255*col(6)[P Infectio]

7 -22.273469802*col(7)[Vaccinated] +1784367.29267*col(3)[cYear] +0*col(4)[pYear] -53.525919591*col(5)[P IVD Outb] +170263592.409*col(6)[P Infectio]

8 -11.11077743*col(7)[Vaccinated] +911353.951632*col(3)[cYear] +0*col(4)[pYear] +13.807677196*col(5)[P IVD Outb] -45713756.303*col(6)[P Infectio]

9 -116.82575042*col(7)[Vaccinated] +9310369.6888*col(3)[cYear] +0*col(4)[pYear] -317.35566339*col(5)[P IVD Outb] +1005708619.49*col(6)[P Infectio]

10 -20.831278083*col(7)[Vaccinated] +1669591.80656*col(3)[cYear] +0*col(4)[pYear] -48.545742859*col(5)[P IVD Outb] +154575126.915*col(6)[P Infectio]

(The validation method randomly assigns rows of data to validation groups,

so the LGO Validation Equations will vary. Since these equations are

generated during the individual validation runs, increasing

'Validate N Times' will not decrease the variability of the coefficients.)

REGRESSION: MULTIPLE (ONE SUBSET)

2021-03-18 06:09:36

Using: C:\Users\LENOVO\Desktop\Influenza-PLOS ONE\Data Influenza PLOS ONE\Medical Data\Model 1.dt

X Columns:

2) Population	4) P IVD Outb	6) Vaccinated	8) Acq Immuni

3) Year X2	5) P Infectio	7) Susceptibi

Y Column: 1) IVD Outbre

Keep If:

Calculate Constant: false

Total number of data points = 9

Number of data points used = 9

Regression equation:

col(1)[IVD Outbre] =

-154.47927995*col(2)[Population]

+9080348.81456*col(3)[Year X2]

+25.9889199358*col(4)[P IVD Outb]

-82572198.698*col(5)[P Infectio]

+0*col(6)[Vaccinated]

+62247146054.7*col(7)[Susceptibi]

+0*col(8)[Acq Immuni]

R^2	= 0.96369918596	AIC = 295.227487479	MSEP	= 1.30221971e15

adj R^2 = 0.91832316841	BIC = 311.602487479	PRESS	= 2.92397545e15

PRE R^2 = 0.00153216413	MAE = 5285609.80942	LOO MAE = 14788243.2131

REGRESSION: MULTIPLE (ONE SUBSET)

2021-03-18 06:12:47

Using: C:\Users\LENOVO\Desktop\Influenza-PLOS ONE\Data Influenza PLOS ONE\Medical Data\Model 1.dt

X Columns:

2) Population	5) P Infectio	7) Susceptibi

4) P IVD Outb	6) Vaccinated	8) Acq Immuni

Y Column: 1) IVD Outbre

Keep If:

Calculate Constant: false

Total number of data points = 9

Number of data points used = 9

Regression equation:

col(1)[IVD Outbre] =

-130.59510233*col(2)[Population]

+25.9887940442*col(4)[P IVD Outb]

-82571815.448*col(5)[P Infectio]

+66.8746953389*col(6)[Vaccinated]

+62246868567.6*col(7)[Susceptibi]

+0*col(8)[Acq Immuni]

R^2	= 0.96369910178	AIC = 293.22750835	MSEP	= 4.34074242e14

adj R^2 =	0.918322979	BIC = 307.10250835	PRESS	= 2.92396567e15

PRE R^2 = 0.00153200328	MAE = 5285617.2247	LOO MAE = 14788225.3586

For each term in the ANOVA table below, if P<=0.05, that term was a

significant source of Y's variation.

Source	SS	df	MS	F	P

------------------------ ------------- -------- --------- --------- ---------

Regression	8.88963273e15	5 1.7779e15 21.238022 .0056 **

col(2)[Population]	8.45240879e15	1 8.4524e15 100.9673 .0006 ***

col(4)[P IVD Outb]	5.60507161e13	1 5.6051e13 0.6695475 .4592 ns

col(5)[P Infectio]	2.78847279e14	1 2.7885e14 3.3309389 .1420 ns

col(6)[Vaccinated]	5.2545784e13	1 5.2546e13 0.6276798 .4725 ns

col(7)[Susceptibi]	4.97801572e13	1 4.978e13 0.5946433 .4837 ns

col(8)[Acq Immuni]	0	0

Error	3.34857273e14	4 8.3714e13

------------------------ ------------- -------- --------- --------- ---------

Total	9.22449e15	9

Table of Statistics for the Regression Coefficients:

Column	Coef. Std Error t(Coef=0)	P	+/-95% CL

------------------------ --------- --------- --------- --------- ---------

col(2)[Population]	-130.5951 172.29869 -0.757958 .4907 ns	478.37786

col(4)[P IVD Outb]	25.988794 53.084794 0.4895713 .6501 ns	147.38702

col(5)[P Infectio]	-8.2572e7 1.66518e8 -0.495874 .6460 ns	4.62327e8

col(6)[Vaccinated]	66.874695 92.276821 0.7247182 .5087 ns	256.20153

col(7)[Susceptibi]	6.2247e10 8.0722e10 0.7711312 .4837 ns	2.2412e11

col(8)[Acq Immuni]	0	0	0

Degrees of freedom for two-tailed t tests = 4

If P<=0.05, the coefficient is significantly different from 0.

Residuals:

Row	Y observed	Y expected	Residual

--------- ------------- ------------- -------------

1	9300000	15902155.532	-6602155.532

2	34000000	28627905.762 5372094.23805

3	30000000 21778124.2852 8221875.71484

4	30000000 28607292.9558 1392707.04424

5	24000000 32492058.8608 -8492058.8608

6	29000000 36284974.5521 -7284974.5521

7	45000000 36926475.0246	8073524.9754

8	36000000 37406088.5538 -1406088.5538

9	38000000	37274924.449 725075.551029

Validation Method: Bootstrap

Validate N Times: 10

Leave-Group-Out PRESS	= 3.14275739e17

Leave-Group-Out PRE R^2 = 0.5774753

Leave-Group-Out MAE	= 116939038.095

(The validation method randomly assigns rows of data to validation groups,

so the Leave-Group-Out statistics printed above will vary.

You can reduce the variability by increasing 'Validate N Times'.)

Group Leave-Group-Out Validation Equations

----- ----------------------------------------------------------------------

1 1093.09820322*col(2)[Population] -700.65080433*col(4)[P IVD Outb] +2204486613.29*col(5)[P Infectio] -627.43887283*col(6)[Vaccinated] -4.9803902e11*col(7)[Susceptibi] +0*col(8)[Acq Immuni]

2 -115.76783319*col(2)[Population] +84.7973725165*col(4)[P IVD Outb] -268911483.06*col(5)[P Infectio] +62.9468262512*col(6)[Vaccinated] +53988965888.5*col(7)[Susceptibi] +0*col(8)[Acq Immuni]

3 348.001484764*col(2)[Population] +174.194441366*col(4)[P IVD Outb] -551010673.62*col(5)[P Infectio] -158.13697335*col(6)[Vaccinated] -1.7213749e11*col(7)[Susceptibi] +0*col(8)[Acq Immuni]

4 63.4308432184*col(2)[Population] +6.33657758534*col(4)[P IVD Outb] -20764735.82*col(5)[P Infectio] -33.56106843*col(6)[Vaccinated] -29760792152*col(7)[Susceptibi] +0*col(8)[Acq Immuni]

5 273.441372247*col(2)[Population] -4510.723456*col(4)[P IVD Outb] +14036451838.7*col(5)[P Infectio] -526.91937766*col(6)[Vaccinated] +0*col(7)[Susceptibi] +0*col(8)[Acq Immuni]

6 1739.78166764*col(2)[Population] -276.09445133*col(4)[P IVD Outb] +868642735.593*col(5)[P Infectio] -918.32104331*col(6)[Vaccinated] -8.1910116e11*col(7)[Susceptibi] +0*col(8)[Acq Immuni]

7 360.361370538*col(2)[Population] -98.312670665*col(4)[P IVD Outb] +306520115.139*col(5)[P Infectio] -195.09586269*col(6)[Vaccinated] -1.67957e11*col(7)[Susceptibi] +0*col(8)[Acq Immuni]

8 -724.26854932*col(2)[Population] +264.035783075*col(4)[P IVD Outb] -824794233.28*col(5)[P Infectio] +395.5016394*col(6)[Vaccinated] +336652307537*col(7)[Susceptibi] +0*col(8)[Acq Immuni]

9 295.712944588*col(2)[Population] -47.603334337*col(4)[P IVD Outb] +148692509.411*col(5)[P Infectio] -156.09568406*col(6)[Vaccinated] -1.3915572e11*col(7)[Susceptibi] +0*col(8)[Acq Immuni]

10 2191.8977859*col(2)[Population] -1360.3534407*col(4)[P IVD Outb] +4289615026.12*col(5)[P Infectio] -1246.6887953*col(6)[Vaccinated] -1.0026764e12*col(7)[Susceptibi] +0*col(8)[Acq Immuni]

(The validation method randomly assigns rows of data to validation groups,

so the LGO Validation Equations will vary. Since these equations are

generated during the individual validation runs, increasing

'Validate N Times' will not decrease the variability of the coefficients.)

REGRESSION: MULTIPLE (ONE SUBSET)

2021-03-18 06:14:12

Using: C:\Users\LENOVO\Desktop\Influenza-PLOS ONE\Data Influenza PLOS ONE\Medical Data\Model 1.dt

X Columns:

4) P IVD Outb	6) Vaccinated	8) Acq Immuni

5) P Infectio	7) Susceptibi

Y Column: 1) IVD Outbre

Keep If:

Calculate Constant: false

Total number of data points = 9

Number of data points used = 9

Regression equation:

col(1)[IVD Outbre] =

25.7493800777*col(4)[P IVD Outb]

-81820236.517*col(5)[P Infectio]

+623.07019002*col(6)[Vaccinated]

-68654952329*col(7)[Susceptibi]

-1306538627.5*col(8)[Acq Immuni]

R^2	= 0.96370655715	AIC = 291.225659768	MSEP	= 2.16992547e14

adj R^2 = 0.91833975358	BIC = 302.600659768	PRESS	= 2.92427027e15

PRE R^2 = 0.00159135993	MAE = 5283153.02419	LOO MAE = 14780490.5869

For each term in the ANOVA table below, if P<=0.05, that term was a

significant source of Y's variation.

Source	SS	df	MS	F	P

------------------------ ------------- -------- --------- --------- ---------

Regression	8.8897015e15	5 1.7779e15 21.242549 .0056 **

col(4)[P IVD Outb]	8.06030783e15	1 8.0603e15 96.303282 .0006 ***

col(5)[P Infectio]	1.2099713e13	1	1.21e13 0.1445655 .7231 ns

col(6)[Vaccinated]	7.09546213e14	1 7.0955e14 8.4775458 .0436 *

col(7)[Susceptibi]	5.95851155e13	1 5.9585e13 0.7119135 .4463 ns

col(8)[Acq Immuni]	4.81626242e13	1 4.8163e13 0.5754394 .4903 ns

Error	3.34788501e14	4 8.3697e13

------------------------ ------------- -------- --------- --------- ---------

Total	9.22449e15	9

Table of Statistics for the Regression Coefficients:

Column	Coef. Std Error t(Coef=0)	P	+/-95% CL

------------------------ --------- --------- --------- --------- ---------

col(4)[P IVD Outb]	25.74938 52.837091 0.4873353 .6515 ns	146.69928

col(5)[P Infectio]	-8.182e7	1.6574e8 -0.493666 .6474 ns	4.60168e8

col(6)[Vaccinated]	623.07019 825.34039 0.7549251 .4923 ns	2291.5123

col(7)[Susceptibi]	-6.865e10 9.1925e10 -0.746855 .4967 ns	2.5523e11

col(8)[Acq Immuni]	-1.3065e9 1.72235e9 -0.758577 .4903 ns	4.78202e9

Degrees of freedom for two-tailed t tests = 4

If P<=0.05, the coefficient is significantly different from 0.

Residuals:

Row	Y observed	Y expected	Residual

--------- ------------- ------------- -------------

1	9300000 15896132.9477 -6596132.9477

2	34000000 28630962.3922 5369037.60781

3	30000000 21775318.6573 8224681.34269

4	30000000 28619606.2456 1380393.75442

5	24000000 32494393.5529 -8494393.5529

6	29000000 36279893.3223 -7279893.3223

7	45000000 36921694.4964 8078305.50356

8	36000000 37403768.6451 -1403768.6451

9	38000000 37278229.4588 721770.541183

Validation Method: Bootstrap

Validate N Times: 10

Leave-Group-Out PRESS	= 1.52835148e16

Leave-Group-Out PRE R^2 = 0.4299023

Leave-Group-Out MAE	= 33781345.1077

(The validation method randomly assigns rows of data to validation groups,

so the Leave-Group-Out statistics printed above will vary.

You can reduce the variability by increasing 'Validate N Times'.)

Group Leave-Group-Out Validation Equations

----- ----------------------------------------------------------------------

1 54.6214988853*col(4)[P IVD Outb] -173863746.27*col(5)[P Infectio] +899.845837638*col(6)[Vaccinated] -99491906663*col(7)[Susceptibi] -1883832765.4*col(8)[Acq Immuni]

2 17.1213948953*col(4)[P IVD Outb] -55571204.668*col(5)[P Infectio] +1470.56459566*col(6)[Vaccinated] -1.6190824e11*col(7)[Susceptibi] -3085679829.3*col(8)[Acq Immuni]

3 -71.975094403*col(4)[P IVD Outb] +222098702.627*col(5)[P Infectio] -89.134118065*col(6)[Vaccinated] +12264087117.7*col(7)[Susceptibi] +164989120.87*col(8)[Acq Immuni]

4 -615.37222963*col(4)[P IVD Outb] +1952028893.92*col(5)[P Infectio] +1436.9592936*col(6)[Vaccinated] -1.433239e11*col(7)[Susceptibi] -3161804283.3*col(8)[Acq Immuni]

5 24.4017722705*col(4)[P IVD Outb] -78345797.435*col(5)[P Infectio] -1.0824041698*col(6)[Vaccinated] +446455273.041*col(7)[Susceptibi] +0*col(8)[Acq Immuni]

6 305.462568978*col(4)[P IVD Outb] -955358799.69*col(5)[P Infectio] +1644.71309152*col(6)[Vaccinated] -1.8639019e11*col(7)[Susceptibi] -3401622004.7*col(8)[Acq Immuni]

7 7.67835908876*col(4)[P IVD Outb] -28602059.293*col(5)[P Infectio] -4.5831847823*col(6)[Vaccinated] +1637290654*col(7)[Susceptibi] +0*col(8)[Acq Immuni]

8 -184.73362415*col(4)[P IVD Outb] +582856298.186*col(5)[P Infectio] -2197.0207466*col(6)[Vaccinated] +247344736453*col(7)[Susceptibi] +4559481239.49*col(8)[Acq Immuni]

9 -70.371592345*col(4)[P IVD Outb] +223214729.654*col(5)[P Infectio] -420.95243026*col(6)[Vaccinated] +48376797278.7*col(7)[Susceptibi] +864285433.831*col(8)[Acq Immuni]

10 7.23471152816*col(4)[P IVD Outb] -22452844.157*col(5)[P Infectio] -569.09550673*col(6)[Vaccinated] +62699225525.4*col(7)[Susceptibi] +1194681327.36*col(8)[Acq Immuni]

(The validation method randomly assigns rows of data to validation groups,

so the LGO Validation Equations will vary. Since these equations are

generated during the individual validation runs, increasing

'Validate N Times' will not decrease the variability of the coefficients.)

REGRESSION: MULTIPLE (ONE SUBSET)

2021-03-18 06:15:02

Using: C:\Users\LENOVO\Desktop\Influenza-PLOS ONE\Data Influenza PLOS ONE\Medical Data\Model 1.dt

X Columns:

4) P IVD Outb	5) P Infectio	6) Vaccinated	8) Acq Immuni

Y Column: 1) IVD Outbre

Keep If:

Calculate Constant: false

Total number of data points = 9

Number of data points used = 9

Regression equation:

col(1)[IVD Outbre] =

-2.4544214499*col(4)[P IVD Outb]

+6675956.50198*col(5)[P Infectio]

+6.68693970743*col(6)[Vaccinated]

-20313080.595*col(8)[Acq Immuni]

R^2	= 0.9586455105	AIC = 290.40055488	MSEP	= 1.48351029e14

adj R^2 = 0.92556191891	BIC = 297.52055488	PRESS	= 2.01824372e15

PRE R^2 = 0.00860757746	MAE = 5915476.63202	LOO MAE = 12277695.911

For each term in the ANOVA table below, if P<=0.05, that term was a

significant source of Y's variation.

Source	SS	df	MS	F	P

------------------------ ------------- -------- --------- --------- ---------

Regression	8.84301593e15	4 2.2108e15 28.976464 .0012 **

col(4)[P IVD Outb]	8.06030783e15	1 8.0603e15 105.64686 .0001 ***

col(5)[P Infectio]	1.2099713e13	1	1.21e13 0.1585916 .7069 ns

col(6)[Vaccinated]	7.09546213e14	1 7.0955e14 9.3000581 .0284 *

col(8)[Acq Immuni]	6.10621655e13	1 6.1062e13 0.8003449 .4120 ns

Error	3.81474075e14	5 7.6295e13

------------------------ ------------- -------- --------- --------- ---------

Total	9.22449e15	9

Table of Statistics for the Regression Coefficients:

Column	Coef. Std Error t(Coef=0)	P	+/-95% CL

------------------------ --------- --------- --------- --------- ---------

col(4)[P IVD Outb]	-2.454421 35.283108 -0.069564 .9472 ns	90.698115

col(5)[P Infectio]	6675956.5 1.10642e8 0.0603383 .9542 ns	2.84415e8

col(6)[Vaccinated]	6.6869397 7.2438668 0.9231174 .3983 ns	18.620952

col(8)[Acq Immuni]	-2.0313e7	22705821	-0.89462 .4120 ns	58367170

Degrees of freedom for two-tailed t tests = 5

If P<=0.05, the coefficient is significantly different from 0.

Residuals:

Row	Y observed	Y expected	Residual

--------- ------------- ------------- -------------

1	9300000 19516580.6245 -10216580.624

2	34000000 26666457.4265 7333542.57349

3	30000000 22413395.8614 7586604.13859

4	30000000 27058643.9893 2941356.01065

5	24000000 30401381.8151 -6401381.8151

6	29000000 35743556.7941 -6743556.7941

7	45000000 37328214.1125 7671785.88749

8	36000000 34913982.7245 1086017.27552

9	38000000 41258464.5688 -3258464.5688

Validation Method: Bootstrap

Validate N Times: 10

Leave-Group-Out PRESS	= 4.11685572e16

Leave-Group-Out PRE R^2 = 0.3848809

Leave-Group-Out MAE	= 52922423.754

(The validation method randomly assigns rows of data to validation groups,

so the Leave-Group-Out statistics printed above will vary.

You can reduce the variability by increasing 'Validate N Times'.)

Group Leave-Group-Out Validation Equations

----- ----------------------------------------------------------------------

1 10.0584470864*col(4)[P IVD Outb] -32631143.149*col(5)[P Infectio] +2.98575800795*col(6)[Vaccinated] -8619479.9213*col(8)[Acq Immuni]

2 8.44908704356*col(4)[P IVD Outb] -30115856.026*col(5)[P Infectio] +11.6002292839*col(6)[Vaccinated] -35360141.535*col(8)[Acq Immuni]

3 -108.08848314*col(4)[P IVD Outb] +338461650.355*col(5)[P Infectio] +27.3467313376*col(6)[Vaccinated] -85339806.256*col(8)[Acq Immuni]

4 -368.30804864*col(4)[P IVD Outb] +1166502526.63*col(5)[P Infectio] +85.5985454019*col(6)[Vaccinated] -271144739.4*col(8)[Acq Immuni]

5 136.261410199*col(4)[P IVD Outb] -425964708.38*col(5)[P Infectio] -14.83131436*col(6)[Vaccinated] +46948331.0819*col(8)[Acq Immuni]

6 -1489.8259379*col(4)[P IVD Outb] +4742467548.92*col(5)[P Infectio] +182.987403638*col(6)[Vaccinated] -585028539.15*col(8)[Acq Immuni]

7 -73.878969518*col(4)[P IVD Outb] +224232884.235*col(5)[P Infectio] +30.4250612015*col(6)[Vaccinated] -94033625.335*col(8)[Acq Immuni]

8 7.01846436905*col(4)[P IVD Outb] -22384563.155*col(5)[P Infectio] +3.22299346485*col(6)[Vaccinated] -9534139.8692*col(8)[Acq Immuni]

9 20.4731541163*col(4)[P IVD Outb] -67653332.11*col(5)[P Infectio] +8.62416776759*col(6)[Vaccinated] -26060304.809*col(8)[Acq Immuni]

10 22.9944401271*col(4)[P IVD Outb] -72549891.762*col(5)[P Infectio] -1.330061447*col(6)[Vaccinated] +4849364.57514*col(8)[Acq Immuni]

(The validation method randomly assigns rows of data to validation groups,

so the LGO Validation Equations will vary. Since these equations are

generated during the individual validation runs, increasing

'Validate N Times' will not decrease the variability of the coefficients.)

REGRESSION: MULTIPLE (ONE SUBSET)

2021-03-18 06:16:02

Using: C:\Users\LENOVO\Desktop\Influenza-PLOS ONE\Data Influenza PLOS ONE\Medical Data\Model 1.dt

X Columns:

4) P IVD Outb	5) P Infectio	6) Vaccinated

Y Column: 1) IVD Outbre

Keep If:

Calculate Constant: false

Total number of data points = 9

Number of data points used = 9

Regression equation:

col(1)[IVD Outbre] =

26.7466892891*col(4)[P IVD Outb]

-84582483.027*col(5)[P Infectio]

+0.2067155247*col(6)[Vaccinated]

R^2	= 0.95202593961	AIC = 289.736870089	MSEP	= 1.14731618e14

adj R^2 = 0.92803890941	BIC = 294.236870089	PRESS	= 1.08696823e15

PRE R^2 = 0.0812154241	MAE = 6052750.52602	LOO MAE = 9579945.39658

For each term in the ANOVA table below, if P<=0.05, that term was a

significant source of Y's variation.

Source	SS	df	MS	F	P

------------------------ ------------- -------- --------- --------- ---------

Regression	8.78195376e15	3 2.9273e15 39.689196 .0002 ***

col(4)[P IVD Outb]	8.06030783e15	1 8.0603e15 109.28336 .0000 ***

col(5)[P Infectio]	1.2099713e13	1	1.21e13 0.1640505 .6995 ns

col(6)[Vaccinated]	7.09546213e14	1 7.0955e14 9.6201777 .0211 *

Error	4.4253624e14	6 7.3756e13

------------------------ ------------- -------- --------- --------- ---------

Total	9.22449e15	9

Table of Statistics for the Regression Coefficients:

Column	Coef. Std Error t(Coef=0)	P	+/-95% CL

------------------------ --------- --------- --------- --------- ---------

col(4)[P IVD Outb]	26.746689	13.17208 2.0305592 .0886 ns	32.230919

col(5)[P Infectio]	-8.4582e7	42130125 -2.007649 .0915 ns	1.03089e8

col(6)[Vaccinated]	0.2067155 0.0666471 3.1016411 .0211 *	0.1630797

Degrees of freedom for two-tailed t tests = 6

If P<=0.05, the coefficient is significantly different from 0.

Residuals:

Row	Y observed	Y expected	Residual

--------- ------------- ------------- -------------

1	9300000 22555298.6018 -13255298.602

2	34000000	29420304.32 4579695.68004

3	30000000 21932602.4523	8067397.5477

4	30000000 26079508.2196 3920491.78038

5	24000000 28858189.3914 -4858189.3914

6	29000000 31940253.2671 -2940253.2671

7	45000000 34317419.3416 10682580.6584

8	36000000 39052080.7886 -3052080.7886

9	38000000 41118767.0188 -3118767.0188

Validation Method: Bootstrap

Validate N Times: 10

Leave-Group-Out PRESS	= 5.73608889e14

Leave-Group-Out PRE R^2 = 0.8095697

Leave-Group-Out MAE	= 12660294.067

(The validation method randomly assigns rows of data to validation groups,

so the Leave-Group-Out statistics printed above will vary.

You can reduce the variability by increasing 'Validate N Times'.)

Group Leave-Group-Out Validation Equations

----- ----------------------------------------------------------------------

1 10.7704514702*col(4)[P IVD Outb] -34359236.419*col(5)[P Infectio] +0.21328594988*col(6)[Vaccinated]

2 19.5833964475*col(4)[P IVD Outb] -60672305.613*col(5)[P Infectio] +0.12787221548*col(6)[Vaccinated]

3 59.158430925*col(4)[P IVD Outb] -185618343.14*col(5)[P Infectio] +0.18862691615*col(6)[Vaccinated]

4 69.7134644441*col(4)[P IVD Outb] -222434433.5*col(5)[P Infectio] +0.31519435293*col(6)[Vaccinated]

5 15.5421411259*col(4)[P IVD Outb] -49252350.398*col(5)[P Infectio] +0.22195872074*col(6)[Vaccinated]

6 19.2728388856*col(4)[P IVD Outb] -61277488.138*col(5)[P Infectio] +0.238073035*col(6)[Vaccinated]

7 16.0051489183*col(4)[P IVD Outb] -48891606.847*col(5)[P Infectio] +0.09151825667*col(6)[Vaccinated]

8 25.6051066111*col(4)[P IVD Outb] -80784916.411*col(5)[P Infectio] +0.19578687744*col(6)[Vaccinated]

9 13.2286048125*col(4)[P IVD Outb] -39750365.02*col(5)[P Infectio] +0.05110646782*col(6)[Vaccinated]

10 25.2179743725*col(4)[P IVD Outb] -79646962.749*col(5)[P Infectio] +0.21795017035*col(6)[Vaccinated]

(The validation method randomly assigns rows of data to validation groups,

so the LGO Validation Equations will vary. Since these equations are

generated during the individual validation runs, increasing

'Validate N Times' will not decrease the variability of the coefficients.)

REGRESSION: MULTIPLE (ONE SUBSET)

2021-03-18 06:17:08

Using: C:\Users\LENOVO\Desktop\Influenza-PLOS ONE\Data Influenza PLOS ONE\Medical Data\Model 1.dt

X Columns:

4) P IVD Outb	5) P Infectio

Y Column: 1) IVD Outbre

Keep If:

Calculate Constant: false

Total number of data points = 9

Number of data points used = 9

Regression equation:

col(1)[IVD Outbre] =

5.55093555005*col(4)[P IVD Outb]

-14393418.985*col(5)[P Infectio]

R^2	= 0.87510610847	AIC = 296.348106564	MSEP	= 2.13348603e14

adj R^2 = 0.83942213946	BIC =	299.3276984	PRESS	= 1.58300785e15

PRE R^2 = 0.04823720607	MAE = 8379907.14424	LOO MAE = 10290717.6272

For each term in the ANOVA table below, if P<=0.05, that term was a

significant source of Y's variation.

Source	SS	df	MS	F	P

------------------------ ------------- -------- --------- --------- ---------

Regression	8.07240755e15	2 4.0362e15 24.523788 .0007 ***

col(4)[P IVD Outb]	8.06030783e15	1 8.0603e15 48.97406 .0002 ***

col(5)[P Infectio]	1.2099713e13	1	1.21e13 0.0735173 .7941 ns

Error	1.15208245e15	7 1.6458e14

------------------------ ------------- -------- --------- --------- ---------

Total	9.22449e15	9

Table of Statistics for the Regression Coefficients:

Column	Coef. Std Error t(Coef=0)	P	+/-95% CL

------------------------ --------- --------- --------- --------- ---------

col(4)[P IVD Outb]	5.5509356	16.82133 0.3299939 .7511 ns	39.776126

col(5)[P Infectio]	-1.4393e7	53084679 -0.271141 .7941 ns	1.25525e8

Degrees of freedom for two-tailed t tests = 7

If P<=0.05, the coefficient is significantly different from 0.

Residuals:

Row	Y observed	Y expected	Residual

--------- ------------- ------------- -------------

1	9300000 19126200.0213 -9826200.0213

2	34000000 8609946.73071 25390053.2693

3	30000000 32031116.7993 -2031116.7993

4	30000000 28748056.4116 1251943.58835

5	24000000 29229948.0793 -5229948.0793

6	29000000 23766791.7429	5233208.2571

7	45000000 29177586.4499 15822413.5501

8	36000000 45983460.3282 -9983460.3282

9	38000000 37349179.5947 650820.405348

Validation Method: Bootstrap

Validate N Times: 10

Leave-Group-Out PRESS	= 7.20626382e14

Leave-Group-Out PRE R^2 = 0.6414745

Leave-Group-Out MAE	= 12108623.0893

(The validation method randomly assigns rows of data to validation groups,

so the Leave-Group-Out statistics printed above will vary.

You can reduce the variability by increasing 'Validate N Times'.)

Group Leave-Group-Out Validation Equations

----- ----------------------------------------------------------------------

1 14.8808788561*col(4)[P IVD Outb] -43768822.694*col(5)[P Infectio]

2 17.4198885846*col(4)[P IVD Outb] -51674508.395*col(5)[P Infectio]

3 -5.6442442485*col(4)[P IVD Outb] +20736887.3773*col(5)[P Infectio]

4 -0.6819447698*col(4)[P IVD Outb] +5378792.86124*col(5)[P Infectio]

5 5.64615360435*col(4)[P IVD Outb] -14653172.591*col(5)[P Infectio]

6 14.7712085538*col(4)[P IVD Outb] -43576008.558*col(5)[P Infectio]

7 44.2607499837*col(4)[P IVD Outb] -135666035.9*col(5)[P Infectio]

8 9.8076664566*col(4)[P IVD Outb] -27729459.915*col(5)[P Infectio]

9 2.04501919772*col(4)[P IVD Outb] -3039283.0458*col(5)[P Infectio]

10 -7.8546297436*col(4)[P IVD Outb] +28830367.2411*col(5)[P Infectio]

(The validation method randomly assigns rows of data to validation groups,

so the LGO Validation Equations will vary. Since these equations are

generated during the individual validation runs, increasing

'Validate N Times' will not decrease the variability of the coefficients.)

For each term in the ANOVA table below, if P<=0.05, that term was a

significant source of Y's variation.

Source	SS	df	MS	F	P

------------------------ ------------- -------- --------- --------- ---------

Regression	8.8896335e15	5 1.7779e15 21.238073 .0056 **

col(2)[Population]	8.45240879e15	1 8.4524e15 100.96754 .0006 ***

col(3)[Year X2]	3.42712986e14	1 3.4271e14 4.093849 .1131 ns

col(4)[P IVD Outb]	4.44276605e13	1 4.4428e13 0.5307069 .5067 ns

col(5)[P Infectio]	303059772530	1 3.0306e11 0.0036202 .9549 ns

col(6)[Vaccinated]	0	0

col(7)[Susceptibi]	4.97810059e13	1 4.9781e13 0.5946548 .4836 ns

col(8)[Acq Immuni]	0	0

Error	3.34856496e14	4 8.3714e13

------------------------ ------------- -------- --------- --------- ---------

Total	9.22449e15	9

Table of Statistics for the Regression Coefficients:

Column	Coef. Std Error t(Coef=0)	P	+/-95% CL

------------------------ --------- --------- --------- --------- ---------

col(2)[Population]	-154.4793 205.22955 -0.752715 .4935 ns	569.80857

col(3)[Year X2]	9080348.8	12529363 0.7247255 .5087 ns	34787089

col(4)[P IVD Outb]	25.98892 53.084606 0.4895754 .6501 ns	147.3865

col(5)[P Infectio]	-8.2572e7 1.66517e8 -0.495878 .6460 ns	4.62326e8

col(6)[Vaccinated]	0	0	0

col(7)[Susceptibi]	6.2247e10 8.0721e10 0.7711386 .4836 ns	2.2412e11

col(8)[Acq Immuni]	0	0	0

Degrees of freedom for two-tailed t tests = 4

If P<=0.05, the coefficient is significantly different from 0.

Residuals:

Row	Y observed	Y expected	Residual

--------- ------------- ------------- -------------

1	9300000 15902106.0503 -6602106.0503

2	34000000 28627935.0257 5372064.97427

3	30000000 21778132.7287 8221867.27126

4	30000000 28607326.8786 1392673.12143

5	24000000 32492058.9946 -8492058.9946

6	29000000 36284975.9503 -7284975.9503

7	45000000 36926454.3258 8073545.67421

8	36000000 37406103.1472 -1406103.1472

9	38000000 37274906.8987 725093.101334

Validation Method: Bootstrap

Validate N Times: 10

Leave-Group-Out PRESS	= 4.24405791e15

Leave-Group-Out PRE R^2 = 0.5822298

Leave-Group-Out MAE	= 27672546.866

(The validation method randomly assigns rows of data to validation groups,

so the Leave-Group-Out statistics printed above will vary.

You can reduce the variability by increasing 'Validate N Times'.)

Group Leave-Group-Out Validation Equations

----- ----------------------------------------------------------------------

1 644.142236597*col(2)[Population] -42321155.978*col(3)[Year X2] -364.27245456*col(4)[P IVD Outb] +1141292347.8*col(5)[P Infectio] +0*col(6)[Vaccinated] -2.4072749e11*col(7)[Susceptibi] +0*col(8)[Acq Immuni]

2 -658.19576607*col(2)[Population] +40134315.0504*col(3)[Year X2] +115.223560106*col(4)[P IVD Outb] -361346008.21*col(5)[P Infectio] +0*col(6)[Vaccinated] +258969503163*col(7)[Susceptibi] +0*col(8)[Acq Immuni]

3 4.7353014494*col(2)[Population] -726891.64314*col(3)[Year X2] -6.0434048141*col(4)[P IVD Outb] +18477352.4421*col(5)[P Infectio] +0*col(6)[Vaccinated] +0*col(7)[Susceptibi] +0*col(8)[Acq Immuni]

4 -550.77273627*col(2)[Population] +34229631.4347*col(3)[Year X2] +184.52204567*col(4)[P IVD Outb] -577161501.48*col(5)[P Infectio] +0*col(6)[Vaccinated] +214018824991*col(7)[Susceptibi] +0*col(8)[Acq Immuni]

5 1181.74828233*col(2)[Population] -71130526.368*col(3)[Year X2] -145.89936235*col(4)[P IVD Outb] +459165725.945*col(5)[P Infectio] +0*col(6)[Vaccinated] -4.6864709e11*col(7)[Susceptibi] +0*col(8)[Acq Immuni]

6 14.0959421389*col(2)[Population] -3766992.6331*col(3)[Year X2] -217.65181484*col(4)[P IVD Outb] +689546829.087*col(5)[P Infectio] +0*col(6)[Vaccinated] +6381773351.6*col(7)[Susceptibi] +0*col(8)[Acq Immuni]

7 625.008326775*col(2)[Population] -36720400.117*col(3)[Year X2] +53.0764240209*col(4)[P IVD Outb] -164738563.25*col(5)[P Infectio] +0*col(6)[Vaccinated] -2.5156915e11*col(7)[Susceptibi] +0*col(8)[Acq Immuni]

8 107.947772863*col(2)[Population] -7700491.5978*col(3)[Year X2] -93.479159576*col(4)[P IVD Outb] +295173467.703*col(5)[P Infectio] +0*col(6)[Vaccinated] -37822092866*col(7)[Susceptibi] +0*col(8)[Acq Immuni]

9 390.299065004*col(2)[Population] -23611851.415*col(3)[Year X2] -58.092012076*col(4)[P IVD Outb] +181320156.742*col(5)[P Infectio] +0*col(6)[Vaccinated] -1.542282e11*col(7)[Susceptibi] +0*col(8)[Acq Immuni]

10 -821.5275453*col(2)[Population] +51082206.2452*col(3)[Year X2] +259.681496305*col(4)[P IVD Outb] -811239600.37*col(5)[P Infectio] +0*col(6)[Vaccinated] +319086897694*col(7)[Susceptibi] +0*col(8)[Acq Immuni]

(The validation method randomly assigns rows of data to validation groups,

so the LGO Validation Equations will vary. Since these equations are

generated during the individual validation runs, increasing

'Validate N Times' will not decrease the variability of the coefficients.)
