## Supplementary File (S6) for "Influenza outbreak, disease transmission rate and mortality risk in the United States (2021 to 2050 Projection)"

Post-Analysis

```
GET DATA
  /TYPE=XLS
  /FILE='C:\Users\LENOVO\Desktop\Influenza-PLOS ONE\Data Influenza PLOS ONE\Medical Data\Vaccination\Post-Analysis.xls'
  /SHEET=name 'Vaccination'
  /CELLRANGE=full
  /READNAMES=on
  /ASSUMEDSTRWIDTH=32767.

Warning.  Command name: GET DATA
(2101) The column contained no recognized type; defaulting to "Numeric[8,2]"
* Column 4
EXECUTE.
DATASET NAME DataSet1 WINDOW=FRONT.
CROSSTABS
  /TABLES=AIObs BY AIEst
  /FORMAT=AVALUE TABLES
  /STATISTICS=CHISQ CC PHI LAMBDA UC ETA CORR GAMMA D BTAU CTAU KAPPA RISK MCNEMAR
  /CELLS=COUNT
  /COUNT ROUND CELL
  /BARCHART.
```

**Crosstabs**

| **Notes** | | |
| --- | --- | --- |
| Output Created | | 20-MAR-2021 11:44:33 |
| Comments | |  |
| Input | Active Dataset | DataSet1 |
|  | Filter | <none> |
|  | Weight | <none> |
|  | Split File | <none> |
|  | N of Rows in Working Data File | 6 |
| Missing Value Handling | Definition of Missing | User-defined missing values are treated as missing. |
|  | Cases Used | Statistics for each table are based on all the cases with valid data in the specified range(s) for all variables in each table. |
| Syntax | | CROSSTABS  /TABLES=AIObs BY AIEst  /FORMAT=AVALUE TABLES  /STATISTICS=CHISQ CC PHI LAMBDA UC ETA CORR GAMMA D BTAU CTAU KAPPA RISK MCNEMAR  /CELLS=COUNT  /COUNT ROUND CELL  /BARCHART. |
| Resources | Processor Time | 00:00:06.61 |
|  | Elapsed Time | 00:00:07.62 |
|  | Dimensions Requested | 2 |
|  | Cells Available | 174762 |

[DataSet1]

| **Case Processing Summary** | | | | | | |
| --- | --- | --- | --- | --- | --- | --- |
|  | Cases | | | | | |
|  | Valid | | Missing | | Total | |
|  | N | Percent | N | Percent | N | Percent |
| AIObs * AIEst | 6 | 100.0% | 0 | 0.0% | 6 | 100.0% |

| **AIObs * AIEst Crosstabulation** | | | | | | | | |
| --- | --- | --- | --- | --- | --- | --- | --- | --- |
| Count | | | | | | | | |
|  | | AIEst | | | | | | Total |
|  |  | 63.1 | 64.5 | 66.0 | 67.5 | 69.1 | 70.8 |  |
| AIObs | 59.6 | 0 | 0 | 1 | 0 | 0 | 0 | 1 |
|  | 63.4 | 0 | 0 | 0 | 0 | 1 | 0 | 1 |
|  | 65.3 | 0 | 0 | 0 | 1 | 0 | 0 | 1 |
|  | 66.7 | 0 | 0 | 0 | 0 | 0 | 1 | 1 |
|  | 68.1 | 0 | 1 | 0 | 0 | 0 | 0 | 1 |
|  | 69.8 | 1 | 0 | 0 | 0 | 0 | 0 | 1 |
| Total | | 1 | 1 | 1 | 1 | 1 | 1 | 6 |

| **Chi-Square Tests** | | | |
| --- | --- | --- | --- |
|  | Value | df | Asymp. Sig. (2-sided) |
| Pearson Chi-Square | 30.000^a^ | 25 | .224 |
| Likelihood Ratio | 21.501 | 25 | .664 |
| Linear-by-Linear Association | .551 | 1 | .458 |
| McNemar-Bowker Test | . | . | .^b^ |
| N of Valid Cases | 6 |  |  |
| a. 36 cells (100.0%) have expected count less than 5. The minimum expected count is .17. | | | |
| b. Both variables must have identical values of categories. | | | |

| **Directional Measures** | | | | | | |
| --- | --- | --- | --- | --- | --- | --- |
|  | | | Value | Asymp. Std. Error^a^ | Approx. T^b^ | Approx. Sig. |
| Nominal by Nominal | Lambda | Symmetric | 1.000 | .000 | 8.660 | .000 |
|  |  | AIObs Dependent | 1.000 | .000 | 5.477 | .000 |
|  |  | AIEst Dependent | 1.000 | .000 | 5.477 | .000 |
|  | Goodman and Kruskal tau | AIObs Dependent | 1.000 | .000 |  | .462^c^ |
|  |  | AIEst Dependent | 1.000 | .000 |  | .462^c^ |
|  | Uncertainty Coefficient | Symmetric | 1.000 | .000 |  | .664^d^ |
|  |  | AIObs Dependent | 1.000 | .000 |  | .664^d^ |
|  |  | AIEst Dependent | 1.000 | .000 |  | .664^d^ |
| Ordinal by Ordinal | Somers' d | Symmetric | -.333 | .407 | -.818 | .413 |
|  |  | AIObs Dependent | -.333 | .407 | -.818 | .413 |
|  |  | AIEst Dependent | -.333 | .407 | -.818 | .413 |
| Nominal by Interval | Eta | AIObs Dependent | 1.000 |  |  |  |
|  |  | AIEst Dependent | 1.000 |  |  |  |
| a. Not assuming the null hypothesis. | | | | | | |
| b. Using the asymptotic standard error assuming the null hypothesis. | | | | | | |
| c. Based on chi-square approximation | | | | | | |
| d. Likelihood ratio chi-square probability. | | | | | | |

| **Symmetric Measures** | | | | | |
| --- | --- | --- | --- | --- | --- |
|  | | Value | Asymp. Std. Error^a^ | Approx. T^b^ | Approx. Sig. |
| Nominal by Nominal | Phi | 2.236 |  |  | .224 |
|  | Cramer's V | 1.000 |  |  | .224 |
|  | Contingency Coefficient | .913 |  |  | .224 |
| Ordinal by Ordinal | Kendall's tau-b | -.333 | .407 | -.818 | .413 |
|  | Kendall's tau-c | -.333 | .407 | -.818 | .413 |
|  | Gamma | -.333 | .407 | -.818 | .413 |
|  | Spearman Correlation | -.486 | .454 | -1.111 | .329^c^ |
| Interval by Interval | Pearson's R | -.332 | .333 | -.703 | .521^c^ |
| Measure of Agreement | Kappa | .000 | .000 | . |  |
| N of Valid Cases | | 6 |  |  |  |
| a. Not assuming the null hypothesis. | | | | | |
| b. Using the asymptotic standard error assuming the null hypothesis. | | | | | |
| c. Based on normal approximation. | | | | | |

| **Risk Estimate** | |
| --- | --- |
|  | Value |
| Odds Ratio for AIObs (59.6 / 63.4) | ^a^ |
| a. Risk Estimate statistics cannot be computed. They are only computed for a 2*2 table without empty cells. | |

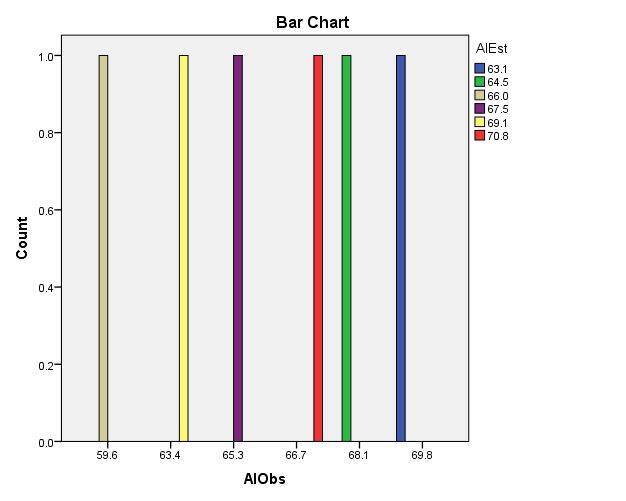

```
T-TEST PAIRS=AIObs VacObs WITH AIEst VacEst (PAIRED)
  /CRITERIA=CI(.9500)
  /MISSING=ANALYSIS.
```

**T-Test**

| **Notes** | | |
| --- | --- | --- |
| Output Created | | 20-MAR-2021 11:52:39 |
| Comments | |  |
| Input | Active Dataset | DataSet1 |
|  | Filter | <none> |
|  | Weight | <none> |
|  | Split File | <none> |
|  | N of Rows in Working Data File | 6 |
| Missing Value Handling | Definition of Missing | User defined missing values are treated as missing. |
|  | Cases Used | Statistics for each analysis are based on the cases with no missing or out-of-range data for any variable in the analysis. |
| Syntax | | T-TEST PAIRS=AIObs VacObs WITH AIEst VacEst (PAIRED)  /CRITERIA=CI(.9500)  /MISSING=ANALYSIS. |
| Resources | Processor Time | 00:00:00.05 |
|  | Elapsed Time | 00:00:00.11 |

[DataSet1]

| **Paired Samples Statistics** | | | | | |
| --- | --- | --- | --- | --- | --- |
|  | | Mean | N | Std. Deviation | Std. Error Mean |
| Pair 1 | AIObs | 65.483 | 6 | 3.6318 | 1.4827 |
|  | AIEst | 66.833 | 6 | 2.8773 | 1.1746 |
| Pair 2 | VacObs | 32953924.17 | 6 | 3041347.910 | 1241625.085 |
|  | VacEst | 33539839.67 | 6 | 372013.950 | 151874.059 |

| **Paired Samples Correlations** | | | | |
| --- | --- | --- | --- | --- |
|  | | N | Correlation | Sig. |
| Pair 1 | AIObs & AIEst | 6 | -.332 | .521 |
| Pair 2 | VacObs & VacEst | 6 | .844 | .035 |

| **Paired Samples Test** | | | | | | | | | |
| --- | --- | --- | --- | --- | --- | --- | --- | --- | --- |
|  | | Paired Differences | | | | | t | df | Sig. (2-tailed) |
|  |  | Mean | Std. Deviation | Std. Error Mean | 95% Confidence Interval of the Difference | |  |  |  |
|  |  |  |  |  | Lower | Upper |  |  |  |
| Pair 1 | AIObs - AIEst | -1.3500 | 5.3294 | 2.1757 | -6.9429 | 4.2429 | -.620 | 5 | .562 |
| Pair 2 | VacObs - VacEst | -585915.500 | 2734797.550 | 1116476.425 | -3455909.517 | 2284078.517 | -.525 | 5 | .622 |

```
GET DATA
  /TYPE=XLS
  /FILE='C:\Users\LENOVO\Desktop\Influenza-PLOS ONE\Data Influenza PLOS ONE\Medical Data\Vaccination\Post-Analysis.xls'
  /SHEET=name 'Infection Rate'
  /CELLRANGE=full
  /READNAMES=on
  /ASSUMEDSTRWIDTH=32767.
EXECUTE.
DATASET NAME DataSet2 WINDOW=FRONT.
DATASET ACTIVATE DataSet2.
DATASET CLOSE DataSet1.
T-TEST PAIRS=IREst WITH IRObs (PAIRED)
  /CRITERIA=CI(.9500)
  /MISSING=ANALYSIS.
```

**T-Test**

| **Notes** | | |
| --- | --- | --- |
| Output Created | | 20-MAR-2021 12:00:38 |
| Comments | |  |
| Input | Active Dataset | DataSet2 |
|  | Filter | <none> |
|  | Weight | <none> |
|  | Split File | <none> |
|  | N of Rows in Working Data File | 9 |
| Missing Value Handling | Definition of Missing | User defined missing values are treated as missing. |
|  | Cases Used | Statistics for each analysis are based on the cases with no missing or out-of-range data for any variable in the analysis. |
| Syntax | | T-TEST PAIRS=IREst WITH IRObs (PAIRED)  /CRITERIA=CI(.9500)  /MISSING=ANALYSIS. |
| Resources | Processor Time | 00:00:00.05 |
|  | Elapsed Time | 00:00:00.22 |

[DataSet2]

| **Paired Samples Statistics** | | | | | |
| --- | --- | --- | --- | --- | --- |
|  | | Mean | N | Std. Deviation | Std. Error Mean |
| Pair 1 | IREst | 9.644 | 9 | .1424 | .0475 |
|  | IRObs | 9.6756 | 9 | 3.12952 | 1.04317 |

| **Paired Samples Correlations** | | | | |
| --- | --- | --- | --- | --- |
|  | | N | Correlation | Sig. |
| Pair 1 | IREst & IRObs | 9 | -.734 | .024 |

| **Paired Samples Test** | | | | | | | | | |
| --- | --- | --- | --- | --- | --- | --- | --- | --- | --- |
|  | | Paired Differences | | | | | t | df | Sig. (2-tailed) |
|  |  | Mean | Std. Deviation | Std. Error Mean | 95% Confidence Interval of the Difference | |  |  |  |
|  |  |  |  |  | Lower | Upper |  |  |  |
| Pair 1 | IREst - IRObs | -.03111 | 3.23550 | 1.07850 | -2.51814 | 2.45591 | -.029 | 8 | .978 |

```
GET DATA
  /TYPE=XLS
  /FILE='C:\Users\LENOVO\Desktop\Influenza-PLOS ONE\Data Influenza PLOS ONE\Medical Data\Vaccination\Post-Analysis.xls'
  /SHEET=name 'Influenza outbreak'
  /CELLRANGE=full
  /READNAMES=on
  /ASSUMEDSTRWIDTH=32767.
EXECUTE.
DATASET NAME DataSet3 WINDOW=FRONT.
T-TEST PAIRS=IVDEst WITH IVDObs (PAIRED)
  /CRITERIA=CI(.9500)
  /MISSING=ANALYSIS.
```

**T-Test**

| **Notes** | | |
| --- | --- | --- |
| Output Created | | 20-MAR-2021 12:05:23 |
| Comments | |  |
| Input | Active Dataset | DataSet3 |
|  | Filter | <none> |
|  | Weight | <none> |
|  | Split File | <none> |
|  | N of Rows in Working Data File | 10 |
| Missing Value Handling | Definition of Missing | User defined missing values are treated as missing. |
|  | Cases Used | Statistics for each analysis are based on the cases with no missing or out-of-range data for any variable in the analysis. |
| Syntax | | T-TEST PAIRS=IVDEst WITH IVDObs (PAIRED)  /CRITERIA=CI(.9500)  /MISSING=ANALYSIS. |
| Resources | Processor Time | 00:00:00.05 |
|  | Elapsed Time | 00:00:00.13 |

[DataSet3]

| **Paired Samples Statistics** | | | | | |
| --- | --- | --- | --- | --- | --- |
|  | | Mean | N | Std. Deviation | Std. Error Mean |
| Pair 1 | IVDEst | 31820140.50 | 10 | 4870073.012 | 1540052.309 |
|  | IVDObs | 29630000.00 | 10 | 9922594.867 | 3137800.008 |

| **Paired Samples Correlations** | | | | |
| --- | --- | --- | --- | --- |
|  | | N | Correlation | Sig. |
| Pair 1 | IVDEst & IVDObs | 10 | -.375 | .285 |

| **Paired Samples Test** | | | | | | | | | |
| --- | --- | --- | --- | --- | --- | --- | --- | --- | --- |
|  | | Paired Differences | | | | | t | df | Sig. (2-tailed) |
|  |  | Mean | Std. Deviation | Std. Error Mean | 95% Confidence Interval of the Difference | |  |  |  |
|  |  |  |  |  | Lower | Upper |  |  |  |
| Pair 1 | IVDEst - IVDObs | 2190140.500 | 12587894.864 | 3980641.872 | -6814697.023 | 11194978.023 | .550 | 9 | .596 |

```
CORRELATIONS
  /VARIABLES=IVDEst IVDObs
  /PRINT=TWOTAIL NOSIG
  /MISSING=PAIRWISE.
```

**Correlations**

| **Notes** | | |
| --- | --- | --- |
| Output Created | | 20-MAR-2021 12:06:48 |
| Comments | |  |
| Input | Active Dataset | DataSet3 |
|  | Filter | <none> |
|  | Weight | <none> |
|  | Split File | <none> |
|  | N of Rows in Working Data File | 10 |
| Missing Value Handling | Definition of Missing | User-defined missing values are treated as missing. |
|  | Cases Used | Statistics for each pair of variables are based on all the cases with valid data for that pair. |
| Syntax | | CORRELATIONS  /VARIABLES=IVDEst IVDObs  /PRINT=TWOTAIL NOSIG  /MISSING=PAIRWISE. |
| Resources | Processor Time | 00:00:00.05 |
|  | Elapsed Time | 00:00:00.05 |

[DataSet3]

| **Correlations** | | | |
| --- | --- | --- | --- |
|  | | IVDEst | IVDObs |
| IVDEst | Pearson Correlation | 1 | -.375 |
|  | Sig. (2-tailed) |  | .285 |
|  | N | 10 | 10 |
| IVDObs | Pearson Correlation | -.375 | 1 |
|  | Sig. (2-tailed) | .285 |  |
|  | N | 10 | 10 |

```
NONPAR CORR
  /VARIABLES=IVDEst IVDObs
  /PRINT=BOTH TWOTAIL NOSIG
  /MISSING=PAIRWISE.
```

**Nonparametric Correlations**

| **Notes** | | |
| --- | --- | --- |
| Output Created | | 20-MAR-2021 12:06:48 |
| Comments | |  |
| Input | Active Dataset | DataSet3 |
|  | Filter | <none> |
|  | Weight | <none> |
|  | Split File | <none> |
|  | N of Rows in Working Data File | 10 |
| Missing Value Handling | Definition of Missing | User-defined missing values are treated as missing. |
|  | Cases Used | Statistics for each pair of variables are based on all the cases with valid data for that pair. |
| Syntax | | NONPAR CORR  /VARIABLES=IVDEst IVDObs  /PRINT=BOTH TWOTAIL NOSIG  /MISSING=PAIRWISE. |
| Resources | Processor Time | 00:00:00.03 |
|  | Elapsed Time | 00:00:00.08 |
|  | Number of Cases Allowed | 174762 cases^a^ |
| a. Based on availability of workspace memory | | |

[DataSet3]

| **Correlations** | | | | |
| --- | --- | --- | --- | --- |
|  | | | IVDEst | IVDObs |
| Kendall's tau_b | IVDEst | Correlation Coefficient | 1.000 | -.315 |
|  |  | Sig. (2-tailed) | . | .209 |
|  |  | N | 10 | 10 |
|  | IVDObs | Correlation Coefficient | -.315 | 1.000 |
|  |  | Sig. (2-tailed) | .209 | . |
|  |  | N | 10 | 10 |
| Spearman's rho | IVDEst | Correlation Coefficient | 1.000 | -.444 |
|  |  | Sig. (2-tailed) | . | .199 |
|  |  | N | 10 | 10 |
|  | IVDObs | Correlation Coefficient | -.444 | 1.000 |
|  |  | Sig. (2-tailed) | .199 | . |
|  |  | N | 10 | 10 |
