## Supplementary File (S9) for "Influenza outbreak, disease transmission rate and mortality risk in the United States (2021 to 2050 Projection)"

Analysis

Mortality-B

REGRESSION: MULTIPLE (ONE SUBSET)

2021-03-21 03:50:50

Using: C:\Users\LENOVO\Desktop\Influenza-PLOS ONE\Data Influenza PLOS ONE\Medical Data\Model 2.dt

X Columns:

4) Vaccinated	7) PMR 10 000

Y Column: 1) A Deaths

Keep If:

Calculate Constant: false

Total number of data points = 31

Number of data points used = 31

Regression equation:

col(1)[A Deaths] =

1.03125799e-4*col(4)[Vaccinated]

-10522.397558*col(7)[PMR 10 000]

R^2	= 0.89143485111	AIC = 486.98353557	MSEP	= 6673436.10413

adj R^2 = 0.88394759946	BIC = 489.249885154	PRESS	= 215684718.138

PRE R^2 = 0.50614235357	MAE = 1859.11476338	LOO MAE = 2017.87392059

For each term in the ANOVA table below, if P<=0.05, that term was a

significant source of Y's variation.

Source	SS	df	MS	F	P

------------------------ ------------- -------- --------- --------- ---------

Regression	1486340905.68	2 7.4317e8 119.06036 .0000 ***

col(4)[Vaccinated]	1461999938.17	1	1.462e9 234.22114 .0000 ***

col(7)[PMR 10 000]	24340967.5062	1 24340968 3.8995687 .0579 ns

Error	181016954.324	29 6241963.9

------------------------ ------------- -------- --------- --------- ---------

Total	1667357860	31

Table of Statistics for the Regression Coefficients:

Column	Coef. Std Error t(Coef=0)	P	+/-95% CL

------------------------ --------- --------- --------- --------- ---------

col(4)[Vaccinated]	1.0313e-4 1.5398e-5 6.6975318 .0000 *** 3.1492e-5

col(7)[PMR 10 000]	-10522.4 5328.5177 -1.974733 .0579 ns	10898.042

Degrees of freedom for two-tailed t tests = 29

If P<=0.05, the coefficient is significantly different from 0.

Validation Method: Bootstrap

Validate N Times: 10

Leave-Group-Out PRESS	= 96106479.9441

Leave-Group-Out PRE R^2 = 0.5534462

Leave-Group-Out MAE	= 2364.78116922

(The validation method randomly assigns rows of data to validation groups,

so the Leave-Group-Out statistics printed above will vary.

You can reduce the variability by increasing 'Validate N Times'.)

Group Leave-Group-Out Validation Equations

----- ----------------------------------------------------------------------

1 1.00683575e-4*col(4)[Vaccinated] -7519.7112075*col(7)[PMR 10 000]

2 9.61074548e-5*col(4)[Vaccinated] -4506.6401044*col(7)[PMR 10 000]

3 9.96805441e-5*col(4)[Vaccinated] -12555.804695*col(7)[PMR 10 000]

4 1.12074257e-4*col(4)[Vaccinated] -12586.769243*col(7)[PMR 10 000]

5 7.48931592e-5*col(4)[Vaccinated] -1521.4210299*col(7)[PMR 10 000]

6 1.08271921e-4*col(4)[Vaccinated] -9711.8175137*col(7)[PMR 10 000]

7 1.04133211e-4*col(4)[Vaccinated] -10438.476962*col(7)[PMR 10 000]

8 8.18370759e-5*col(4)[Vaccinated] -5373.6986601*col(7)[PMR 10 000]

9 9.19283667e-5*col(4)[Vaccinated] -7560.4867438*col(7)[PMR 10 000]

10 1.13051681e-4*col(4)[Vaccinated] -20376.853885*col(7)[PMR 10 000]

(The validation method randomly assigns rows of data to validation groups,

so the LGO Validation Equations will vary. Since these equations are

generated during the individual validation runs, increasing

'Validate N Times' will not decrease the variability of the coefficients.)

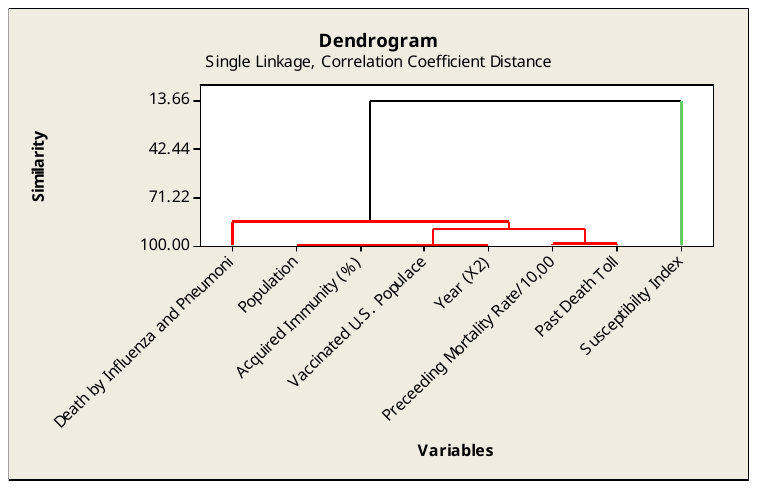

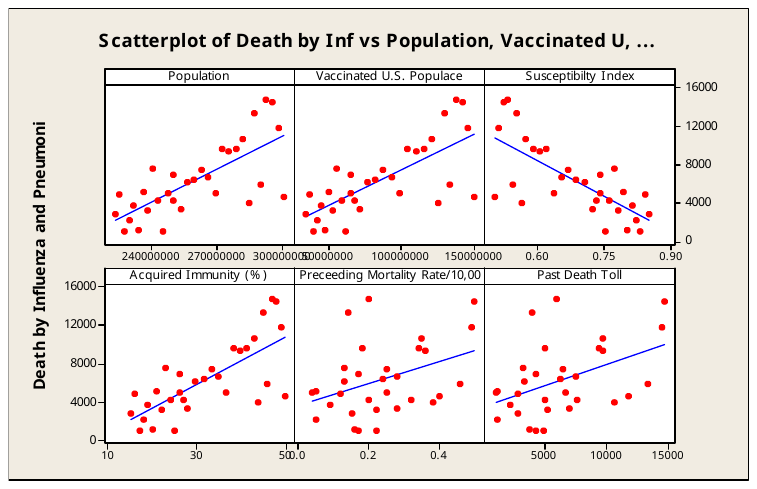

Regression line

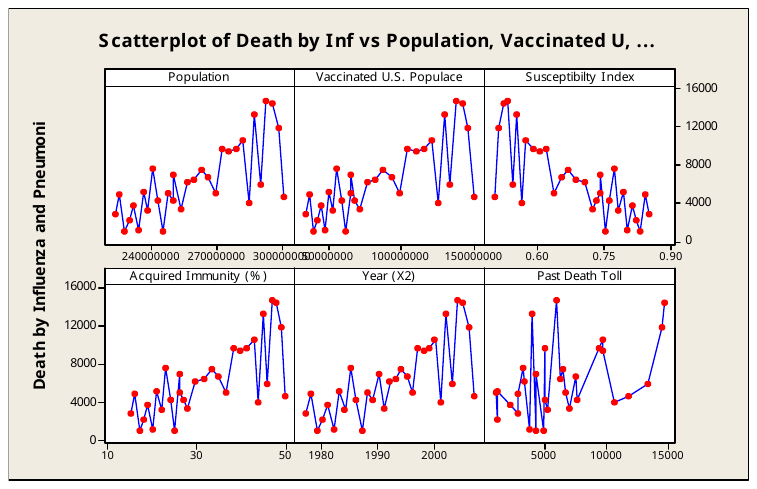

Connect line
